# Auditory white matter tract development in infants exposed to HIV and antiretrovirals

**DOI:** 10.64898/2026.03.09.26347575

**Authors:** Amy S Graham, Barbara Laughton, Francesca Little, Andre van der Kouwe, Mamadou Kaba, Ernesta M Meintjes, Marcin Jankiewicz, Martha J Holmes

**Author notes:** **Correspondence:** Amy Graham.

## Abstract

Although HIV exposure has previously been found to affect brain white matter (WM) tract integrity and language development in infants and children, the impacts of HIV and antiretroviral therapy (ART) exposure on central auditory tracts remain unclear. Moreover, no research to date has investigated the relationship between auditory WM tract development and language outcomes in infants exposed to HIV but uninfected (iHEU). Brain images were acquired at the age of 0-5 weeks for 31 infants whose mothers began ART pre-conception (iHEU-pre), 29 infants whose mothers began ART post-conception (iHEU-post) and 25 infants who were HIV-unexposed (iHU). Full-probabilistic diffusion tensor imaging (DTI) tractography was used to assess WM integrity in tracts connected to central auditory structures. Language assessments were carried out at 9-14 months using the Griffiths Mental Development Scales (GMDS). Linear regression analysis was used to compare DTI tractography results between iHEU and iHU and to assess the relationship between DTI measures and language. Finally, the impacts of HIV and ART exposure on associations between language and DTI measures were visualised using groupwise language-DTI correlation plots. There were no results after multiple comparison correction. Unadjusted results show recurring patterns of reduced fractional anisotropy (FA), driven by iHEU-post, in auditory tracts of iHEU compared to iHU. Both iHEU-pre and iHEU-post contributed to the patterns of uncorrected elevations in mean diffusivity (MD) observed in the entire iHEU group, with the left medial geniculate nucleus being the auditory structure most frequently observed within the affected tracts. Effect sizes of uncorrected differences, which were small-to-moderate in size, were similar to other infant DTI tractography studies. Groupwise assessment of the data revealed moderately strong correlations between GMDS language scores and DTI measures in some affected tracts, only for iHU. Our findings indicate that HIV/ART exposure may have subtle effects on auditory WM tract development in infants. Delays in auditory tract maturation appear to occur irrespective of ART exposure duration and may be HIV exposure-specific effects. Tracts connected to the left auditory thalamus have notably been implicated in our unadjusted results. HIV and ART exposure may interfere with the way in which auditory WM tracts mature, potentially impacting the role of a small number of these tracts in language processing.

## Introduction

Delayed development or disruptions to the auditory system in infancy and early childhood can have long-term implications for language development and communication ^1–4^. The risk of hearing loss and other auditory-related complications is greater in children with perinatal human immunodeficiency virus (HIV) infection ^5–7^. Moreover, exposure to HIV and antiretrovirals during pregnancy, without infection, may also affect hearing and language development in infants and children ^7–11^. As a greater proportion of pregnant women living with HIV are receiving effective antiretroviral therapy (ART), there is an ever-increasing population of infants who are HIV-exposed *in utero* but uninfected (iHEU).

Studies have explored the influence of HIV and ART exposure on the infant central auditory system in terms of the auditory brainstem response ^11–15^. Yet these studies do not provide a consistent narrative, which may in part result from differences in geographical location and the HIV status of the infants themselves not always being clearly defined. While the two studies carried out in Nigeria have reported no HIV exposure effects ^13,15^, poorer outcomes have been reported for iHEU in South Africa ^11,12^. Moreover, Khoza-Shangase and Nesbitt (2023) have found that it is particularly at an inner ear and brainstem level that iHEU display poorer hearing outcomes compared to iHU, whereas in tests of middle ear function they showed that these groups performed similarly. Yet despite research indicating that the central auditory system may be impacted, particularly among iHEU in South Africa, to our knowledge no research has been done to explore the effects of HIV/ART exposure on the auditory system beyond the brainstem.

Diffusion tensor imaging (DTI), based on measures of water diffusion, is useful for investigating the integrity and arrangement of white matter (WM) tracts in the brain ^16,17^. The main measures of DTI include fractional anisotropy (FA), a measure of axonal integrity, and mean diffusivity (MD), an indicator of WM organization. These measures can be better understood based on complementary measures of axial diffusivity (AD) and radial diffusivity (RD), which quantify diffusion in directions parallel or orthogonal to axonal fibers, respectively. Based on these diffusion measures, full-probabilistic tractography establishes the most likely pathways between pre-determined regions of interest ^18–21^.

The narratives drawn from previous DTI studies investigating HIV and ART exposure suggest a risk of localized delays in WM development. However, the literature does not present a consistent picture of the most vulnerable region(s) or the nature of the changes. Although some studies have shown elevated FA in infants and children who are HEU ^22–24^, others have reported reduced FA ^25^ or unchanged FA measures ^26^. Moreover, while DTI studies report altered MD ^23,25^, others have found similar MD between iHEU and iHU ^22,24,26^. The lack of consensus may be related to different methodologies, maternal treatment effects or the different ages across studies.

The Healthy Baby Study was initiated to explore the effects of HIV exposure in a South African neonate cohort, and to determine the impact of differing timing and duration of *in utero* ART exposure. IHEU born to mothers on ART prior to conception (iHEU-pre), and a second group of iHEU whose mothers initiated treatment in the second trimester (iHEU-post), were compared to a control group of infants who were HIV-unexposed (iHU). A recent study in this cohort by Magondo et al. (2024) explored HIV/ART exposure effects on infant WM tract development using tractography, demonstrating that for infants born to mothers on ART throughout pregnancy there is greater MD in select tracts.

Although studies report subtle HIV exposure effects on the developing brain, the associated functional consequences are unclear. Two studies recently reported HIV exposure effects on structural MRI measures and language performance in early childhood ^27,28^. They further demonstrated relationships between these brain imaging measures and language across their cohort. While a larger hippocampus corresponded with better language scores ^27^, children with reduced thickness of a prefrontal cortex region also performed better in language assessments ^28^. This would suggest that there is scope for exploring the relationship between language and other brain imaging measures within an HIV exposure cohort.

Research has shown that WM tract development in early infancy, particularly the arcuate fasciculus and corpus callosum, correlates with language development in later infancy and childhood ^29–32^. In particular, positive associations between FA in WM tracts and language performance have been demonstrated in healthy participants ^29,30,33,34^, premature participants ^32,35^ and participants at greater risk of autism spectrum disorder ^36^. Whereas MD has been shown to be negatively associated with language outcomes in infants and children that are healthy ^33,34^, preterm ^35,37^ or born below the weight of 1kg ^38^.

We present a probabilistic DTI tractography study of infants from the HBS. The goal of our research was to establish whether *in utero* exposure to HIV and ART affects auditory WM tract development in newborns. More specifically, we aimed to explore whether exposure to ART for a greater period of time during pregnancy has a neuroprotective role. We hypothesized that iHEU would have altered FA and MD in auditory tracts when compared to iHU, and that these differences would be influenced by the duration and timing of ART exposure during pregnancy. To better understand the functional links between auditory WM and language, secondary analysis investigated whether measures of WM tract integrity and development could predict language development in later infancy.

## Materials and methods

### Participants

Of the 225 mothers initially enrolled during pregnancy, a total of 195 mother-infant pairs were ultimately enrolled into the Healthy Baby Study (HBS). Of these, 69 mothers did not have HIV, while 126 mothers were living with HIV. Further, 67 mothers with HIV were on ART pre-conception, and the remaining 59 mothers with HIV initiated treatment post-conception. Infants born to these mothers were categorized as iHEU-pre and iHEU-post, respectively. iHEU received nevirapine and zidovudine treatment after birth, tailored according to risk (maternal viral load > 1000 copies/mL indicates high risk) and infant feeding methods. Ethics approval for this research was obtained from the Universities of Cape Town (557/2020) and Stellenbosch. Parents provided written informed consent.

Infants were categorised into risk groups according to maternal viral loads when the infants were at 8 months gestational age, and this guided postnatal treatment provided to infants. High-risk infants, born to mothers with a viral load >1000 copies/mL, received a 6-week course of both nevirapine and zidovudine. Low-risk infants, however, were provided with nevirapine alone.

### Brain imaging

Prior to scanning, infants were put to sleep and placed comfortably in a pillow containing Styrofoam beads. No sedatives were used; infants were scanned during natural sleep. MRI scans were acquired using a 3 T Skyra scanner (Siemens, Erlangen, Germany) at the Cape Universities Body Imaging Centre (CUBIC), Groote Schuur Hospital (University of Cape Town).

A multi-echo magnetization prepared rapid gradient echo (MEMPRAGE) sequence was used to obtain T1-weighted structural scans (inversion time (TI)=1450 ms; repetition time (TR)=2540 ms; echo times (TE)= 1.69/3.55/5.41/7.27 ms; number of slices = 144; field of view = 192 x 192 mm^2^; voxel size = 1x1x1 mm^3^). To account for motion in real time while acquiring these T1-weighted images, volumetric navigators were utilized within the protocol ^39^.

Two sets of diffusion-weighted volumes with opposite phase encoding gradients (anterior (A)-posterior (P) and PA) were acquired using a twice refocused multi-band spin-echo Echo Planar Imaging (EPI) sequence ^40^ which included a navigator ^41^. The sequence parameters were TR=4800 ms; TE=84 ms; 62 slices; voxel size = 2x2x2 mm^3^, GRAPPA=2, SMS=2, 6/8 partial Fourier encoding. Each set comprised 30 different diffusion directions with b=1000 mm/s^2^ and an additional 6 with b = 0 mm/s^2^ b(0).

### Image processing

Pre-processing of the imaging data began with running Infant FreeSurfer (ver.4a14499) on T1-weighted structural images ^42,43^. The structures included from automated segmentation were the left (L) and right (R) cerebellar cortex, L & R hippocampus; L & R ventral diencephalon, L & R amygdala, L & R putamen, L & R accumbens area; L & R pallidum, L & R caudate, L & R lateral ventricle, L & R cerebral cortex, L & R thalamus, 3^rd^ ventricle, 4^th^ ventricle, midbrain, vermis, pons, medulla.

The 3^rd^ ventricle was included due to its close proximity to important structures in the diencephalon, namely the thalamus and hypothalamus ^44^. Meanwhile, as the 4^th^ ventricle lies between the medulla, pons and cerebellum ^44^, studying tracts connected to this region could enhance our understanding of connections within the brainstem.

To explore WM tracts of the auditory system, manual tracing of eight auditory structures was done by a neuroanatomist. The structures of interest were the L & R inferior colliculus, L & R Heschl’s gyrus, L & R cochlear nuclei (CN) and L & R medial geniculate nucleus (MGN) (Figure 1). In total, 28 regions-of-interest (ROIs) from Infant FreeSurfer automated segmentation and the eight manually traced structures were combined and used as seeds for full-probabilistic DTI tractography.

**Figure 1:**
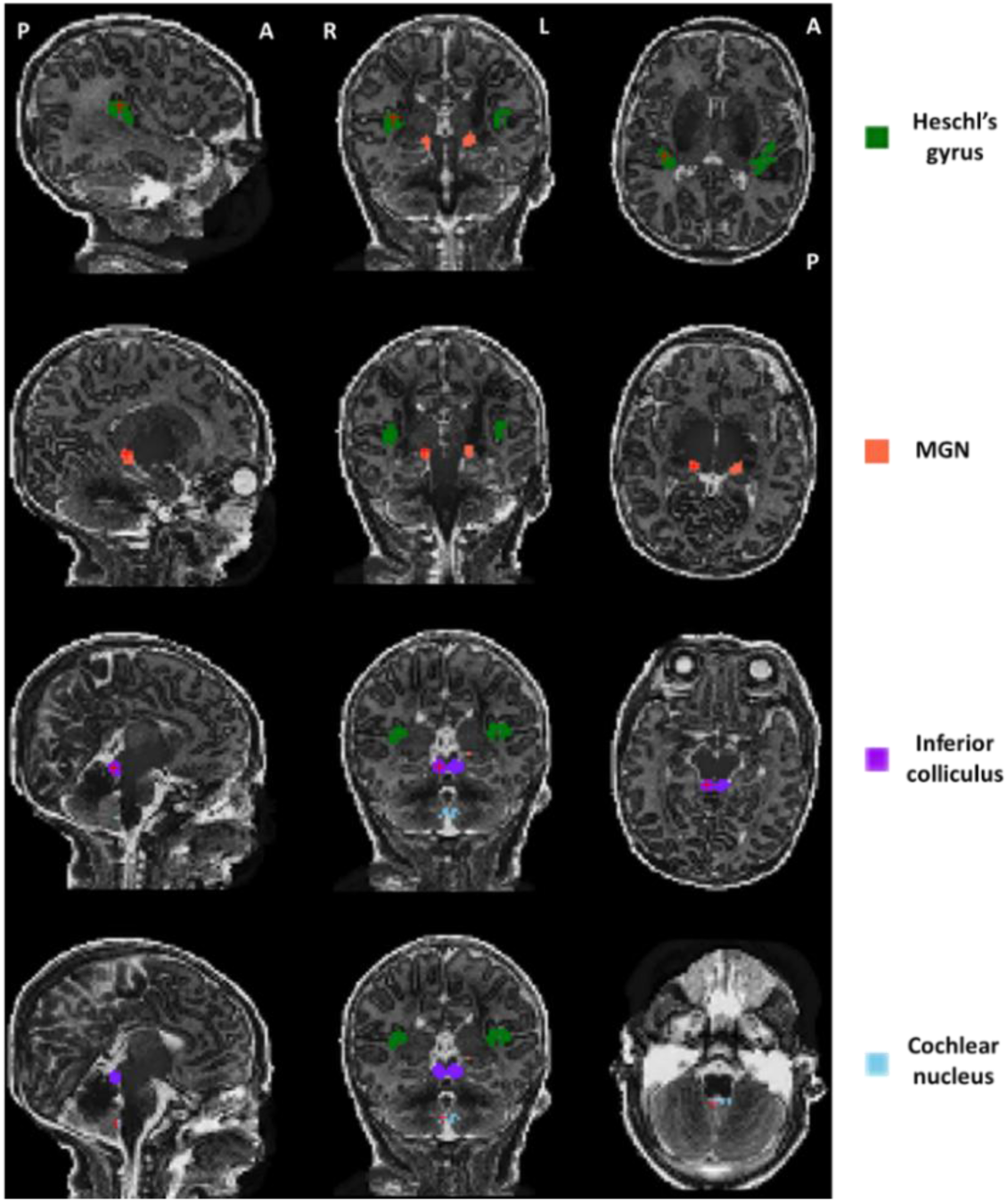
Sagittal, coronal and axial brain images from a randomly selected infant showing the auditory structures of interest in native space (MGN: Medial geniculate nucleus).

A manual check of the diffusion weighted (DW) volumes was done to identify and remove those with distortions or slices with signal dropout. Of the 30 possible diffusion directions, subjects were required to have at least 15 in order to be included (mean for this cohort=25 ± 4). Moreover, they required at least one of the six b0 volumes. TORTOISE (v.3.1.0) tools were implemented for pre-processing. Specifically, DIFF_PREP was used to perform motion correction and correct eddy current distortions, while the Diffeomorphic Registration for Blip-UP blip-Down Diffusion Imaging (DR-BUDDI) function was used to combine AP and PA directions, perform EPI distortion correction and create a final set of diffusion-weighted volumes for each infant ^45^.

Further processing was done using a combination of Functional And Tractographic Connectivity Toolbox (FATCAT) and AFNI (Analysis of Functional NeuroImages) tools. Firstly, diffusion tensors were estimated, following which FA, MD, AD and RD were derived using the three eigenvectors and three eigenvalues of these tensors. Transformation of segmented structural data into DTI space was achieved with 3dAllineate, a tool developed for the alignment of imaging data. 3dROIMaker ^19^ was then used to create a map of labelled ROIs, before inflating the gray matter ROIs to achieve greater overlap with WM regions.

Full-probabilistic tractography was carried out with the 3dTrackID tool ^19,46^ to identify connections between every pair of seeds. Whole brain tractography, which allowed connections between auditory and non-auditory structures to be assessed, used an angle threshold of 60 degrees, a minimum length threshold of 15mm and an FA threshold of 0.1. To isolate intra-auditory tracts specifically, we additionally performed tractography between the auditory structures alone. The angle and length thresholds were optimized for this, with a maximum angle of 55 and minimum length of 10 mm being selected. Ultimately, we took the intersect of tracts that were found in all infants and, therefore, we could be more confident that these were biological connections. Tracts from whole-brain and intra-auditory tractography were combined to provide a final dataset for analysis.

Quality control was done to check that manually traced structures mapped correctly with structural scans and to confirm that structural scans were correctly transformed into DTI space. Whole-brain and intra-auditory tractography outputs were manually checked. We include images of three different tracts which featured among our results, showing these for five infants (Supplementary figures 1-3).

### Language assessments

Psychological testing was carried out by clinicians at the Family Centre for Research with Ubuntu (Stellenbosch University). The Griffiths Mental Development Scales (GMDS) ^47^, which have previously been used within the South African context ^48,49^, and the updated Griffiths III ^50,51^ scales, were used to assess neurodevelopmental outcomes - including language development - at the age of 9-14 months. Only 55 infants included in this study had language assessments carried out. Scores for both the GMDS and Griffiths III scales were available for all of these infants. The language subscale evaluations involved a number of tasks, testing the ability to both comprehend (receptive language) and to express oneself (expressive language), and provide both a raw language score and a Quotient. These scores are standardized according to normal development in British children, whereby they have a mean of 100 and a standard deviation (SD) of 16 ^49^. Scores within 2 SD’s of the mean are considered within expected range, those 1SD to 2SD below the mean are categorised as below average, and those more than 2SD below the mean indicate developmental delay.

### Statistical analysis

Statistical analysis was carried out using R software ^52^. Confounding variables were identified for inclusion in multivariable linear regression models. Relationships between HIV/ART exposure and potential confounders were assessed using the Chi-square, Kruskal-Wallis and Wilcoxon tests from the stats package in R ^52^. Simple linear regression models were used to assess associations of potential confounders with DTI measures and language scores. We considered variables to be confounders if they were associated at p ≤ 0.05 with both the independent and dependent variables being investigated and were not on the path between independent and dependent variable. We included variables that were associated with either FA or MD in more than 10% of the WM connections. Sex, gestational age at birth, birth weight, head circumference at scan, length at scan, maternal weight change during pregnancy (per week), maternal birth age, alcohol exposure and maternal education were each individually assessed to identify potential confounders.

Our final analysis included connections involving at least one auditory seed. Outliers were identified as values greater than 1.5 times the interquartile range, lying above or below the upper and lower quartiles, respectively. Linear regression models were run with and without outliers, and outliers were only excluded if influential.

Linear regression analysis was done to explore the association between HIV/ART exposure and DTI measures in auditory connections. The results of this analysis were visualized in networks plots, created using the iGraph package ^53^. In iHEU specifically, the association between maternal CD4 count and DTI measures was examined using linear regression. These associations were explored only across the auditory connections in which we observed differences between iHEU and iHU, to assess the contribution of maternal immune health.

Secondary analysis examined the relationships between DTI tractography measures in auditory connections and language outcomes at 9-14 months for all auditory tracts and across all infants using simple linear regression analysis. We visualised DTI measure-language outcome relationships separately for iHEU and iHU, and calculated correlations separately for each group, focusing only on tracts in which we had previously found groupwise differences in DTI measures. The same was done for our ART exposure groups. Boxplots of the groupwise language results, and plots showing DTI-language relationships separately for HIV and ART exposure groups, were created using ggplot2 ^54^. The output from all regression models was standardized using the MuMIn package in R ^55^.

We provide measures of effect size. Simple regression models were used to calculate r values, and from these r^2^ values were calculated for each of the predictors included in our regression models. These, together with all observed p values, have been included in the supplementary material. Small, medium and large effect sizes were interpreted as r^2^ values above 0.04, 0.25 and 0.64, and by r values of 0.2, 0.5 and 0.8 ^56^.

As the other HIV exposure DTI studies do not provide measures of effect size, this makes it challenging to compare our findings and to establish what a meaningful effect size is in the context of HIV exposure and WM tracts in infancy. Furthermore, several carry out voxel-wise analysis and/or focus on older children ^23–26^. Yet infant probabilistic tractography studies exploring HIV/ART exposure effects ^57^ and WM injury in premature birth ^58^, have previously provided groupwise means and standard deviations for anatomically similar tracts to those which featured in our results. While the authors did not provide measures of effect size themselves, we could use the information provided by the authors, and the means and standard deviations in our results, to calculate Cohen’s d values ^59^ from which we could then compare our results to these other studies. Supplementary equations 1 & 2 ^60^ were used to calculate pooled standard deviations for groups and then to calculate Cohen’s d. This effect size measure accounts for variation in the data and enables a comparison to be made between other infant studies and our analysis. Cohen’s d values of 0.2, 0.5 and 0.8 are considered small, moderate and large respectively ^59^.

We assessed our results in two ways. Firstly, we used cut-off values to identify statistically relevant results in relation to the null hypothesis. To account for risk of false positives, we corrected for multiple comparisons using the false discovery rate (FDR) approach^61^. As correction for multiple comparisons also increases the risk of false negatives, we report both adjusted and unadjusted results using conventional cut-offs of q ≤ 0.05 and p ≤ 0.05 respectively. For tracts reported at these cutoffs, the 95% CIs were above zero. Secondly, to determine whether our results are meaningful, measures of effect size were also considered. Based on these two different assessments, we could identify the most interesting findings for discussion.

## Results

### Demographics

A total of 186 infants from the HBS had MRI visits, from which T1 images were acquired for 167 infants and DTI images were obtained for 152 infants. In all, DTI scans were successfully processed for 108 infants, however two infants were excluded as their structural scans could not be manually traced due to poor quality. Following quality control steps, DTI data were additionally excluded from analysis for six infants due to suboptimal alignment of DTI and structural images and for 11 infants due to poor tractography output. As four infants were missing important intra-auditory connections, their DTI data were also excluded. The final cohort of 85 infants included 25 iHU and 60 iHEU (Table 1). The study group could be further divided into 31 iHEU-pre and 29 iHEU-post. In our study, infants were scanned at the mean age of 12.39 (±7.49) days.

**Table 1:**
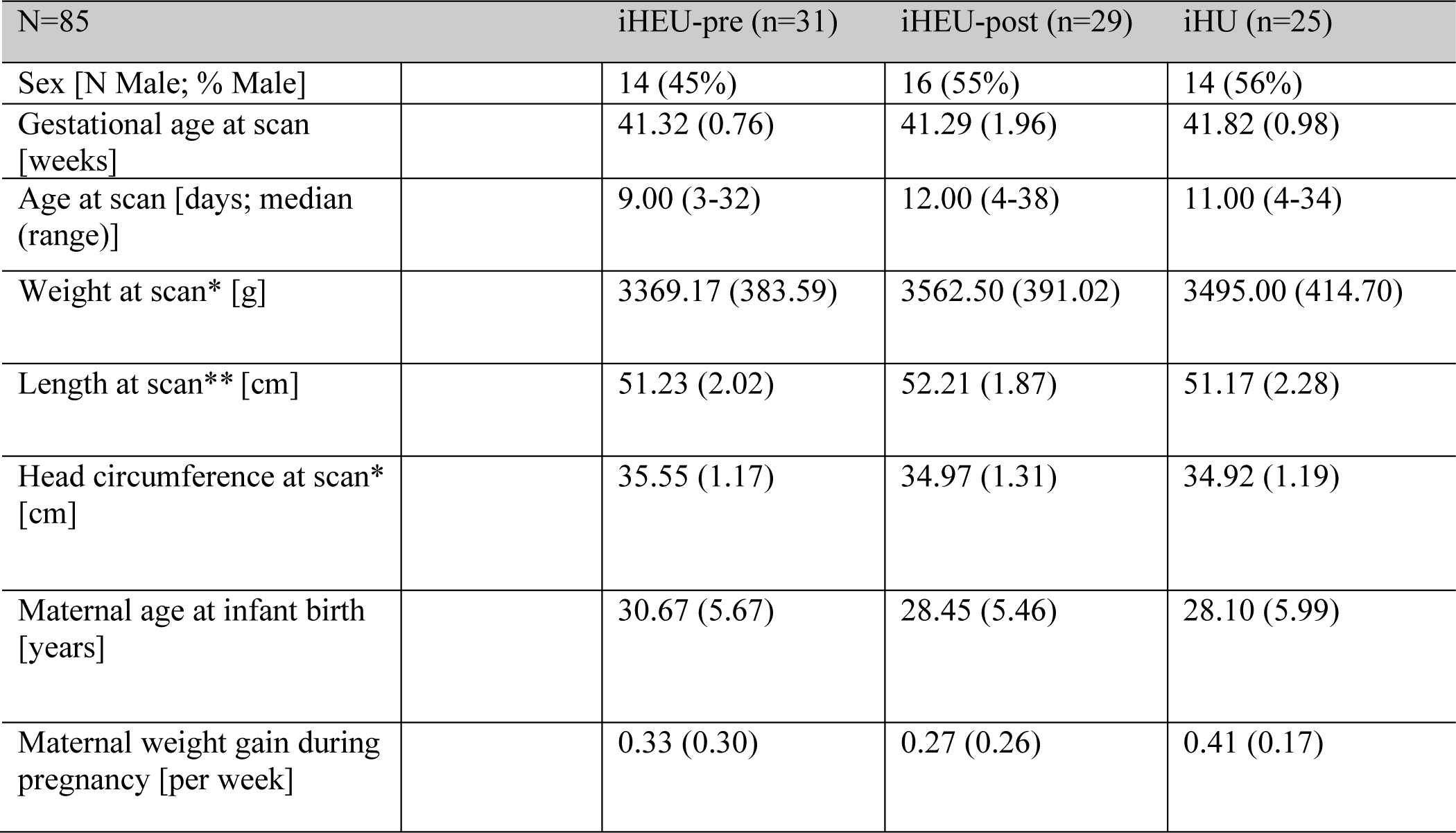
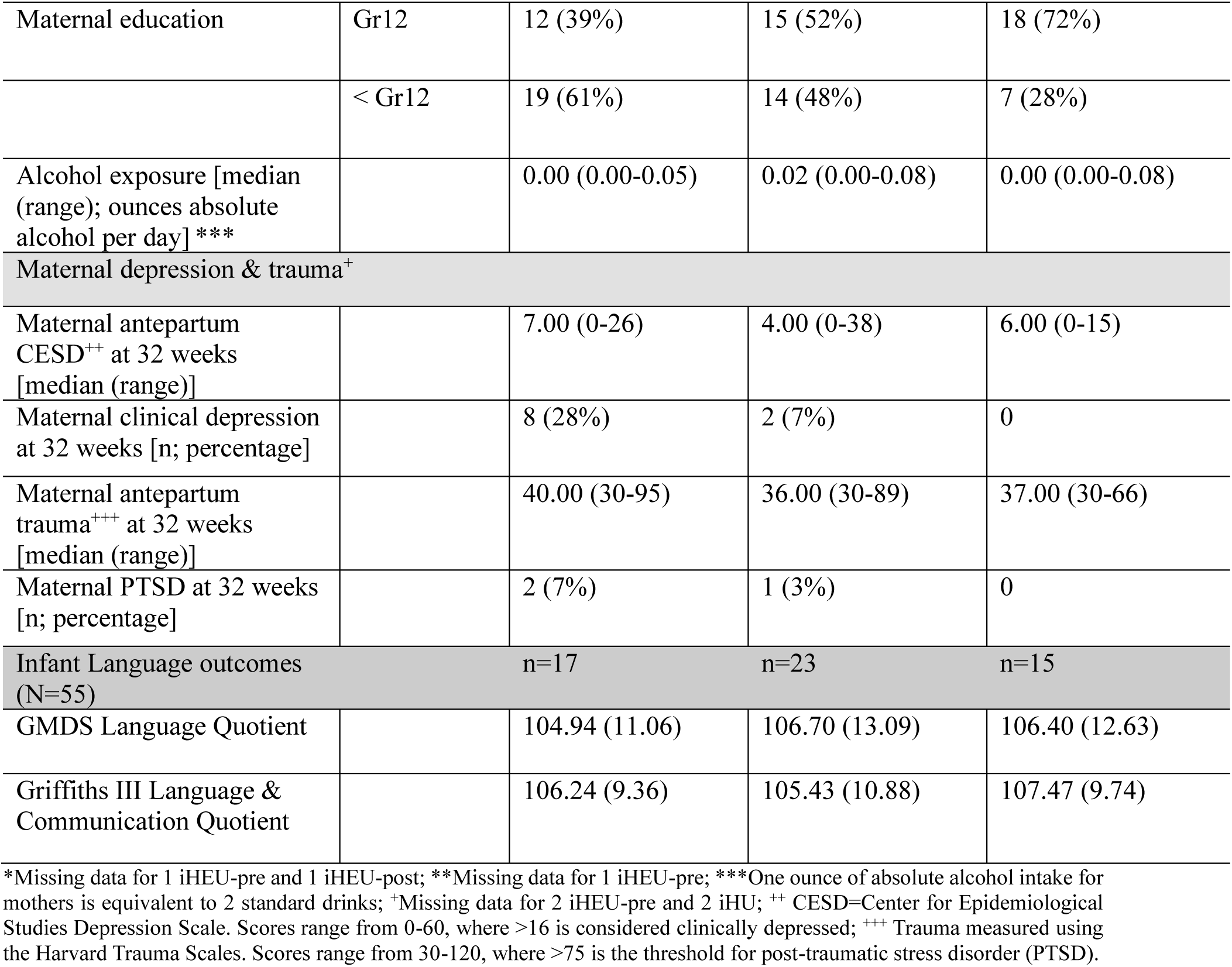
Demographics for infants and mothers summarized as mean and standard deviation according to ART exposure group.

| N=85 | iHEU-pre (n=31) | iHEU-post (n=29) | iHU (n=25) |
| --- | --- | --- | --- |
| Sex [N Male; % Male] | 14 (45%) | 16 (55%) | 14 (56%) |
| Gestational age at scan [weeks] | 41.32 (0.76) | 41.29 (1.96) | 41.82 (0.98) |
| Age at scan [days; median (range)] | 9.00 (3-32) | 12.00 (4-38) | 11.00 (4-34) |
| Weight at scan* [g] | 3369.17 (383.59) | 3562.50 (391.02) | 3495.00 (414.70) |
| Length at scan** [cm] | 51.23 (2.02) | 52.21 (1.87) | 51.17 (2.28) |
| Head circumference at scan* [cm] | 35.55 (1.17) | 34.97 (1.31) | 34.92 (1.19) |
| Maternal age at infant birth [years] | 30.67 (5.67) | 28.45 (5.46) | 28.10 (5.99) |
| Maternal weight gain during pregnancy [per week] | 0.33 (0.30) | 0.27 (0.26) | 0.41 (0.17) |

|  |  |  |  |  |
| --- | --- | --- | --- | --- |
| Maternal education | Gr12 | 12 (39%) | 15 (52%) | 18 (72%) |
|  | < Gr12 | 19 (61%) | 14 (48%) | 7 (28%) |
| Alcohol exposure [median (range); ounces absolute alcohol per day] *** |  | 0.00 (0.00-0.05) | 0.02 (0.00-0.08) | 0.00 (0.00-0.08) |
| Maternal depression & trauma<sup>+</sup> |  |  |  |  |
| Maternal antepartum CESD<sup>++</sup> at 32 weeks [median (range)] |  | 7.00 (0-26) | 4.00 (0-38) | 6.00 (0-15) |
| Maternal clinical depression at 32 weeks [n; percentage] |  | 8 (28%) | 2 (7%) | 0 |
| Maternal antepartum trauma<sup>+++</sup> at 32 weeks [median (range)] |  | 40.00 (30-95) | 36.00 (30-89) | 37.00 (30-66) |
| Maternal PTSD at 32 weeks [n; percentage] |  | 2 (7%) | 1 (3%) | 0 |
| Infant Language outcomes (N=55) |  | n=17 | n=23 | n=15 |
| GMDS Language Quotient |  | 104.94 (11.06) | 106.70 (13.09) | 106.40 (12.63) |
| Griffiths III Language & Communication Quotient |  | 106.24 (9.36) | 105.43 (10.88) | 107.47 (9.74) |

\*Missing data for 1 iHEU-pre and 1 iHEU-post; \*\*Missing data for 1 iHEU-pre; \*\*\*One ounce of absolute alcohol intake for mothers is equivalent to 2 standard drinks; <sup>+</sup>Missing data for 2 iHEU-pre and 2 iHU; <sup>++</sup> CESD=Center for Epidemiological Studies Depression Scale. Scores range from 0-60, where >16 is considered clinically depressed; <sup>+++</sup> Trauma measured using the Harvard Trauma Scales. Scores range from 30-120, where >75 is the threshold for post-traumatic stress disorder (PTSD).

The mean duration of *in utero* ART exposure and maternal CD4 counts are summarized in Table 2. Per definition, iHEU-pre were exposed to ART *in utero* for a longer duration than iHEU-post (p=<0.001). Mothers who began early treatment also had higher CD4 counts than mothers who began treatment during pregnancy (p=0.010). IHEU who were considered low risk were placed on a single round of nevirapine treatment after birth, whereas high-risk infants received additional treatment. One iHEU-pre infant received a second round of nevirapine, while six (1 iHEU-pre and 5 iHEU-post) received additional zidovudine treatment.

**Table 2:**
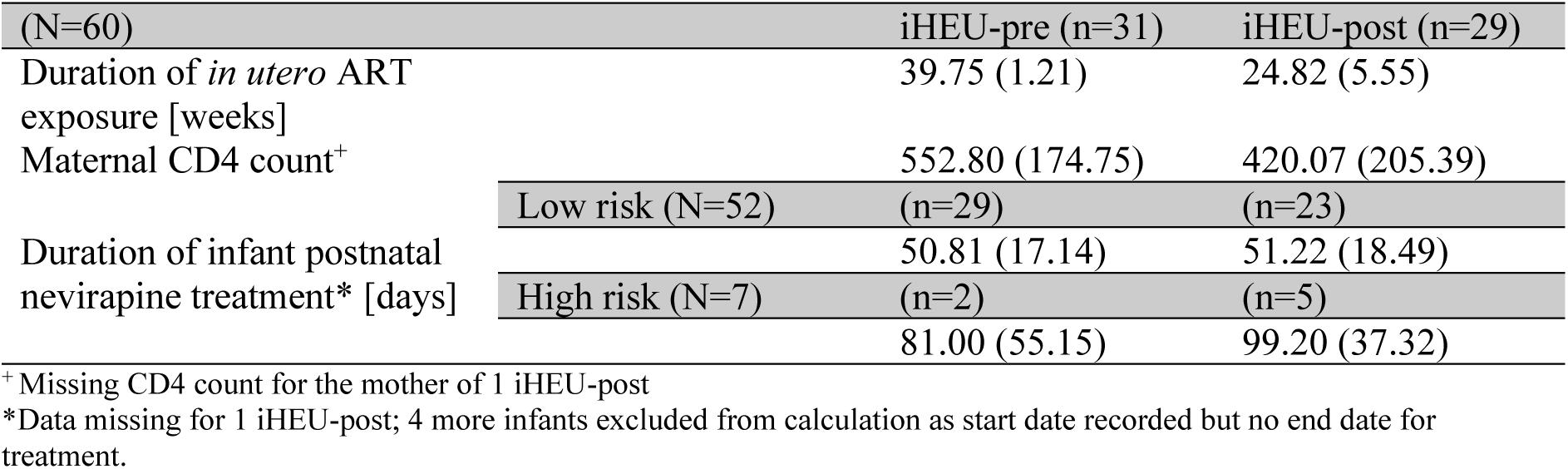
Summary of HIV and ART-related clinical data, providing mean and standard deviation values for both iHEU groups.

All mothers of iHEU included in this analysis received tenofovir disoproxil fumarate (TDF), emtricitabine (FTC) and efavirenz (EFV) treatment during pregnancy. Two mothers (on ART pre conception) had an interruption in this treatment and then restarted the same regimen. Seven mothers (three on ART pre conception; four post conception) were placed on an additional round of dolutegravir (DTG), lamivudine (3TC) and TDF, and one mother (on ART pre conception) took an additional round of DTG, FTC and TDF. One mother (on ART post conception) had an interruption in TDF, FTC, EFV treatment and then switched to DTG, 3TC, TDF, while one mother (on ART post conception) switched treatment twice, firstly to nevirapine (NVP), TDF, FTC and then to DTG, 3TC, TDF.

Our final analysis included eight manually traced auditory structures and 28 structures from automated segmentation. In total there were 390 connections in the brain that were identified in all infants, of which we assessed the 106 connections involving at least one auditory structure – we refer to these as auditory connections. We identified maternal weight gain and education as potential confounders for investigating the associations of HIV/ART exposure with DTI measures. When controlling for these variables in our linear regression models, maternal education was categorised as a binary variable based on whether or not mothers had completed grade 12.

### Impact of HIV/ART exposure on FA in auditory WM pathways

When using linear regression to compare FA between iHEU and iHU, we did not find any connections that showed differences after multiple comparison correction. We report six unadjusted results, in which iHEU were found to have lower FA compared to iHU (Figure 2a, Supplementary figure 4). These results do not include intra-auditory connections, but rather involve connections from auditory structures to non-auditory structures of the brain.

**Figure 2:**
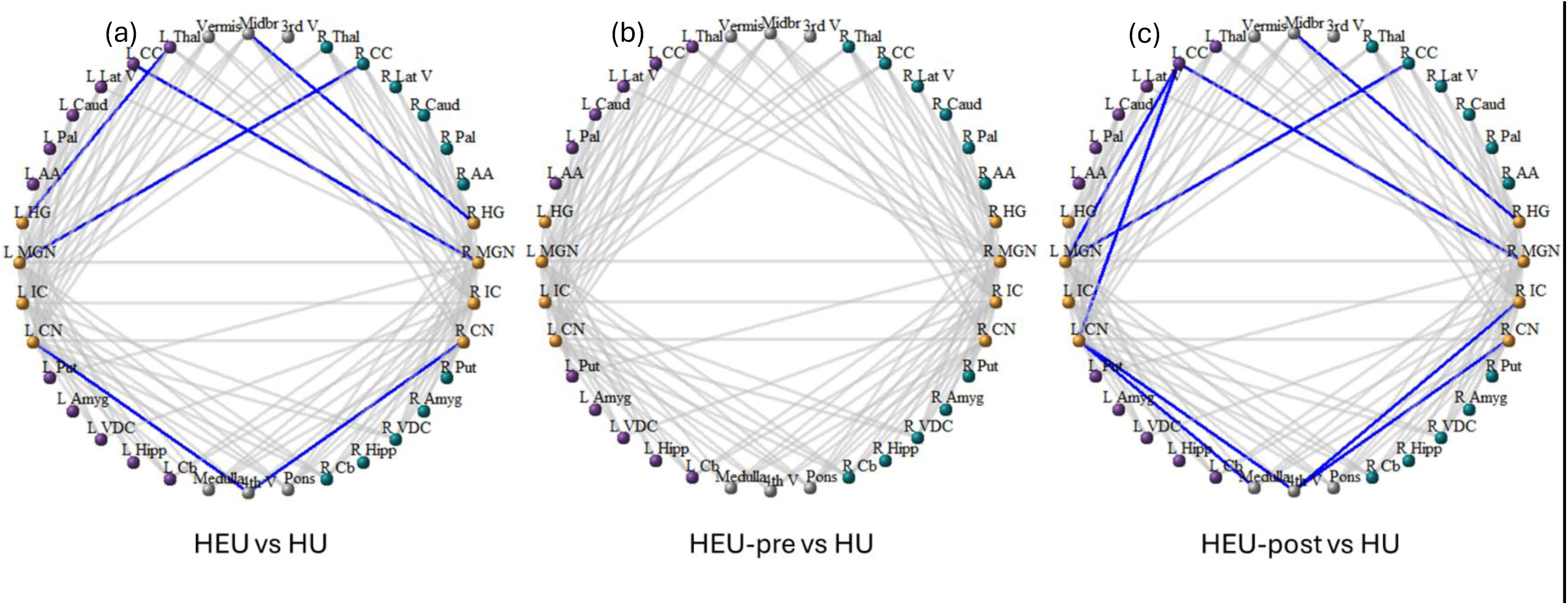
Auditory connections for which differences in FA were found in (a) all HEU (b) HEU-pre and (c) HEU-post infants compared to HU infants. Gray lines indicate auditory connections in which no FA differences were observed; reduced FA with unadjusted p values ≤ 0.05 = blue. *Cb=cerebellar cortex; Hipp=hippocampus; VDC=ventral diencephalon; Amyg=*Amygdala*; Put=putamen; AA= accumbens area; Pal= pallidum; Caud=caudate; Lat V=lateral ventricle; CC=cerebral cortex; Thal=thalamus; CN=cochlear nucleus; IC=inferior colliculus; MGN=medial geniculate nucleus; HG=Heschl’s gyrus; 3^rd^ V = 3^rd^ ventricle; 4^th^ V = 4^th^ ventricle*.

Correlations (r values) between HIV/ART exposure and FA in these 6 tracts ranged from −0.23 to −0.32 (Supplementary table 1). The maximum amount of variation that HIV/ART exposure accounted for in any auditory tract was 10%, as seen by our r^2^ values.

While reductions in FA were accompanied by elevated RD in four connections, no differences in RD were observed after multiple comparison correction (Supplementary figure 5; Supplementary table 1).

### Impact of ART exposure duration on FA in auditory WM pathways

Linear regression analysis comparing ART exposure groups (Figure 2b & c) did not find differences in FA for iHEU-post compared to iHU after FDR correction. Yet we report nine unadjusted results in connections with reduced FA for iHEU-post (Supplementary figure 6). As with analysis comparing all iHEU to iHU, none were for intra-auditory connections. In iHEU-pre, we did not observe any unadjusted differences in FA compared to iHU.

### Impact of HIV/ART exposure on MD in auditory WM pathways

We found no HIV/ART exposure group differences in MD after FDR corrections in iHEU compared to iHU. Prior to multiple comparison correction, greater MD was found for 22 auditory connections (Figure 3a, Supplementary table 1, Supplementary figure 7). Correlations between HIV/ART exposure and MD in these 22 tracts ranged from 0.20 to 0.34. The associated r^2^ values indicate that HIV/ART exposure accounted for at most 11.5% variation in MD for any auditory tract. More than half of the auditory tracts in which higher MD is reported involve the MGN, while 41% were specifically connected to the L MGN. Our unadjusted results show that connections in the left hemisphere are impacted to a greater extent by HIV/ART exposure than connections in the right hemisphere.

**Figure 3:**
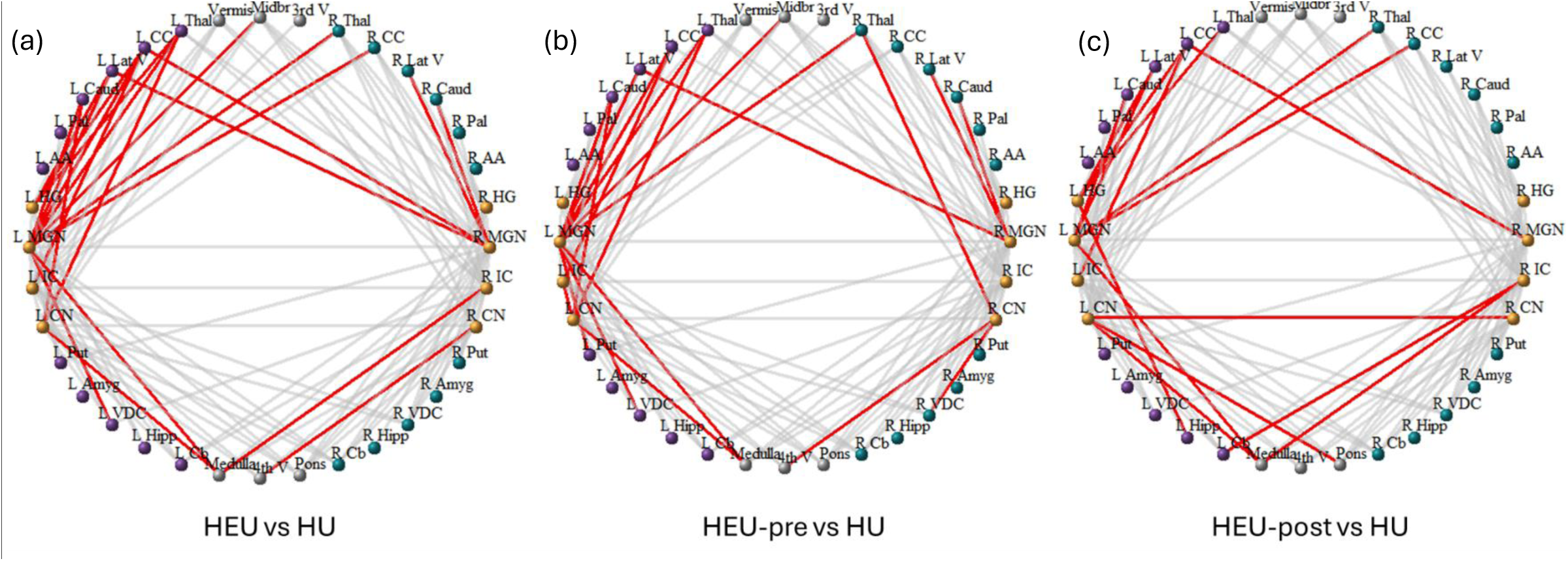
Auditory connections for which we found differences in MD in (a) all HEU (b) HEU-pre and (c) HEU-post infants compared to HU infants. Gray lines indicate auditory connections in which no MD differences were observed; elevated MD with unadjusted p values ≤ 0.05 = red. *(For list of abbreviations see* Figure 2)

In the auditory tracts for which we found elevated MD in iHEU, this was accompanied by elevated unadjusted RD and AD results in many cases (Supplementary figure 5; Supplementary table 1). RD was higher in iHEU for two thirds of connections displaying elevated MD. In total, AD was elevated in 11 of the 22 connections which had elevated MD.

### Impact of ART exposure duration on MD in auditory WM pathways

Further comparisons of iHEU-pre and iHEU-post to iHU yielded no group differences after multiple comparison corrections. Unadjusted results demonstrated that iHEU-pre had greater MD measures compared to iHU for 20 auditory connections, while iHEU-post had higher MD in 16 auditory connections (Figure 3b & c; Supplementary figure 8). Both ART exposure groups contribute to differences in MD observed in iHEU. Of the 22 connections in which our overall analysis found higher MD for iHEU vs iHU, six of these were evident in both iHEU-pre and iHEU-post. Elevated MD in 10 connections was largely driven by the iHEU-pre group, while for six connections the iHEU-post group drove the overall increase in MD observed for all iHEU

While connections involving the L MGN were affected in both iHEU-pre and iHEU-post groups, elevated MD in connections to the R MGN was more often detected for iHEU-pre when compared to iHU. Elevations in RD and/or AD generally corresponded with elevations in MD for both iHEU-pre and iHEU-post.

### Coinciding impacts of HIV/ART exposure on FA and MD in auditory WM pathways

We observed concurrent elevations in MD and reductions in FA for several tracts (Supplementary table 2). In iHEU we observed this pattern in the tracts connecting the L thalamus to L Heschl’s gyrus and the 4^th^ ventricle to the R cochlear nucleus. For both iHEU and iHEU-post, higher MD and lower FA were found for two interhemispheric connections: between the L cerebral cortex and R MGN and between the R cerebral cortex and L MGN. Additionally, the combination was observed in iHEU-post alone for the tract connecting the medulla and L cochlear nucleus.

### Comparison of effect sizes to other infant tractography studies

Visually we observed greater pooled standard deviations for our strongest (unadjusted) findings compared to those for the adjusted results presented by Magondo et al. (2024) (Supplementary figures 9 & 10). The average pooled standard deviation for our strongest FA results was twice as great as that of Magondo et al. (2024) (0.016 vs 0.008). Moreover, the mean pooled standard deviation for our unadjusted MD results of 0.067 was also greater than the value of 0.053 obtained for the results that Magondo et al. (2024) reported.

To compare effect sizes from our data to existing literature, we calculated Cohen’s d for two of our tracts, as well as anatomically comparable tracts in infant studies that used similar methods (Table 3). When considering differences in FA and MD between iHEU and iHU in our cohort for the thalamocortical tract connecting the R CC to the L MGN, which featured in both our FA and MD results, we found that our results were comparable in effect size to those of thalamocortical tract results presented by Magondo et al. (2024) and Liu et al. (2012). Groupwise differences in DTI measures were seen after multiple comparison correction in both of these other studies.

**Table 3:** Effect size calculations for tracts from our study and similar tracts from two other infant tractography studies. Groupwise means and standard deviations for FA and MD in tracts, together with the pooled standard deviation (S_pooled_) and Cohen’s d calculated based on these values using supplementary equations 1 & 2.

| Results from this study |  |  |  |  |
| --- | --- | --- | --- | --- |
| | iHU (n=25) | iHEU (n=60) | $S_{\text{pooled}}$ | Cohen's d |
| FA in R CC – L MGN | 0.257(0.015) | 0.247(0.012) | 0.013 | 0.775 |
| MD in R CC – L MGN | 1.317(0.053) | 1.350(0.054) | 0.054 | -0.615 |
| FA in L Caud – L MGN | 0.226(0.018) | 0.225(0.016) | 0.017 | 0.060 |
| MD in L Caud – L MGN | 1.192(0.053) | 1.222(0.051) | 0.052 | -0.581 |
| Results from Liu et al. (2012) |  |  |  |  |
| | Premature without WM injury (n=41) | Premature with WM injury (n=27) | $S_{\text{pooled}}$ | Cohen's d |
| FA in L sensory STR | 0.24(0.03) | 0.22(0.03) | 0.03 | 0.67 |
| MD in L sensory STR | 1.32(0.08) | 1.37(0.11) | 0.09 | -0.54 |
| Results from Magondo et al. (2024) |  |  |  |  |
| | iHU (n=35) | iHEU-pre (n=36) | $S_{\text{pooled}}$ | Cohen's d |
| FA in L CC – R<br>Thal | 0.235(0.009) | 0.228(0.011) | 0.010 | 0.696 |
| MD in L Caud –<br>L Thal | 1.219(0.043) | 1.247(0.048) | 0.046 | -0.614 |

*L=left; R=right; Caud=caudate; CC=cerebral cortex; Thal=thalamus; STR=superior thalamic radiation. We provide results to 3 decimal places where possible, although in the case of Liu et al. (2012) we were limited as they only provided measures to 2 decimal places. No results were found after multiple comparison correction for this study, while results included from Liu et al. (2012) and Magondo et al. (2024) were reported after multiple comparison correction.*

The tract connecting the L caudate to the L MGN, which only featured in our MD results, was found to have a Cohen’s d value for MD which was moderate in size (Table 3). This was similar in magnitude to the corresponding L caudate – L thalamus tract in the study by Magondo et al. (2024), and that reported by Liu et al. (2012) for the difference in MD in their thalamocortical connection.

### The influence of maternal CD4 on auditory WM pathways

When assessing the relationships between maternal CD4 count and DTI measures across the auditory connections in iHEU, we did not find any associations with either FA or MD for tracts in which we had previously found HIV/ART exposure group differences (Supplementary tables 3 & 4).

### Language development

Linear regression analysis comparing GMDS language quotient scores found no differences when comparing iHEU to iHU (Figure 4a; p=0.904). Similarly, no differences were observed when comparing iHEU-pre (p=0.741) and iHEU-post (p=0.943) to iHU (Supplementary Figure 11a). Furthermore, when looking at the Griffiths III language and communication quotient scores, the results of analysis comparing iHEU to iHU (Figure 4b; p=0.580) and comparing iHEU-pre and iHEU-post to iHU (Supplementary Figure 11b; p=0.733 & p=0.548), similarly found no differences.

**Figure 4:**
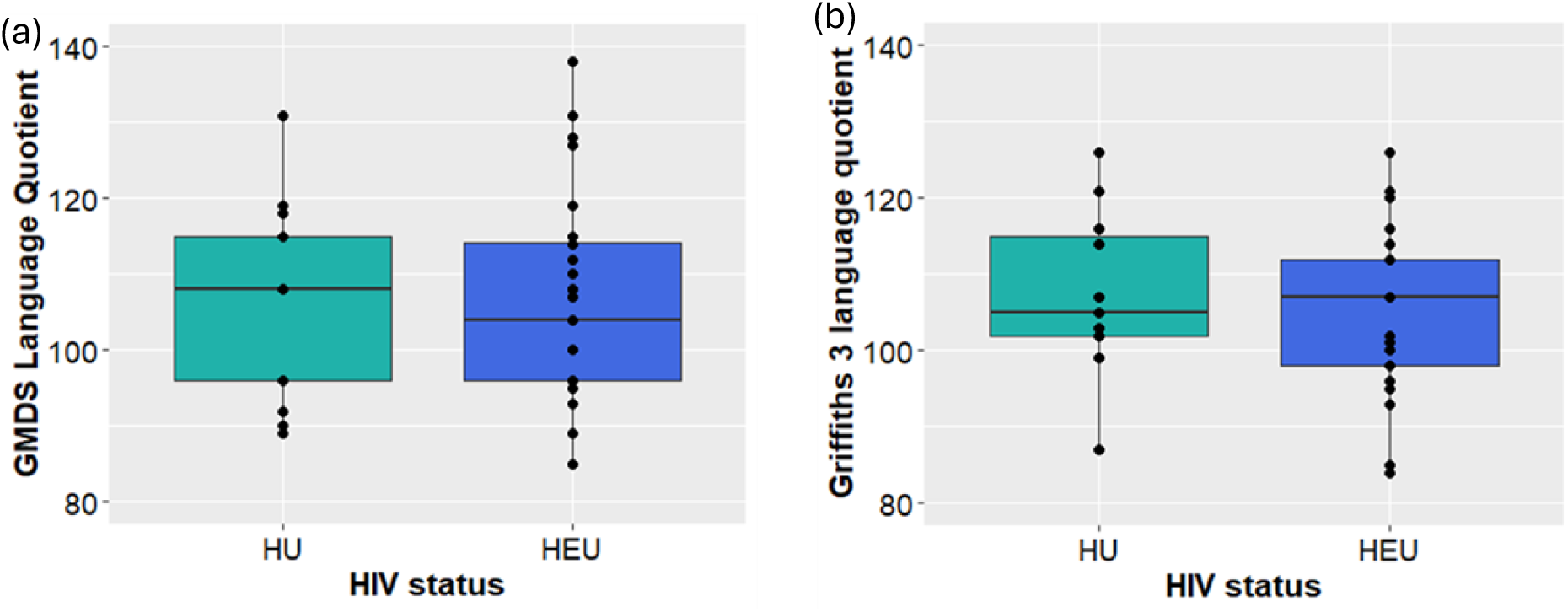
A box and whisker plot summarizing the upper, middle and lower quartiles of (a) GMDS language quotient and (b) Griffiths III language and communication quotient scores according to infants who were HEU and HU; whiskers include data within 1.5 times the interquartile range above and below the upper and lower quartiles.

### Relationship between auditory tract FA and 9-14-month language scores

Analysis exploring the relationships between GMDS language scores at 9-14 months and FA for all neonate auditory connections (Supplementary table 5), found a positive unadjusted association between language performance and FA for a single auditory tract between the R pallidum and R MGN (standardised β = 0.314; p = 0.020). This tract did not correspond with those in which we had observed groupwise FA differences.

We visualised whether HIV/ART exposure influences the relationships between FA and language development, specifically for tracts in which we had previously detected FA differences between iHEU and iHU (Supplementary figure 12). Based on these plots and groupwise correlations, FA was positively correlated with language scores in iHU in two tracts (Figure 5). Between the 4^th^ ventricle & L cochlear nucleus and between the 4^th^ ventricle & R cochlear nucleus – we observed correlation values greater than 0.5 (p < 0.05), indicating moderately strong correlations. Yet iHEU did not display positive FA-language correlations for these tracts.

**Figure 5:**
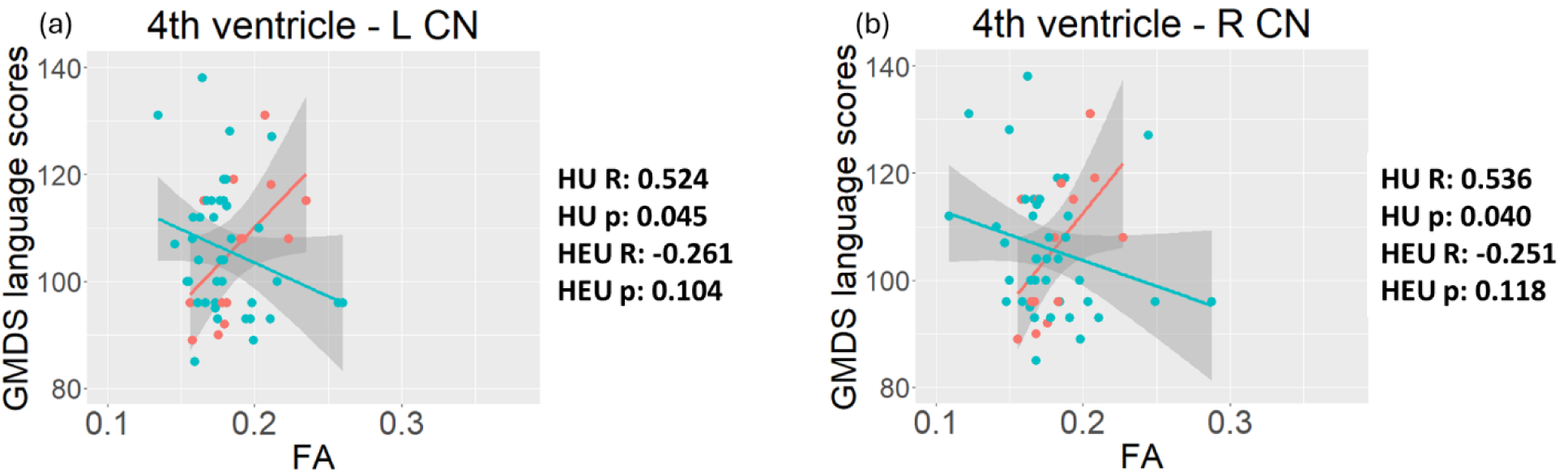
Tracts in which we observe positive correlations between fractional anisotropy (FA) and language outcomes in iHU. Red=iHU; Blue =iHEU.

Additional plots were generated to visualise ART exposure effects on the relationship between FA and language, for tracts in which we had previously observed differences in FA between iHEU-post and iHU (Supplementary figure 13). In four of the nine tracts reported, we observed positive correlations with correlation coefficients greater than 0.5 in iHU. Whereas no clear relationships between FA and language score were observed for these auditory connections among iHEU-post.

### Relationship between auditory tract MD and 9-14-month language scores

There were 4 tracts for which we observed associations between MD and language scores across all infants (Supplementary table 6). In the tracts connecting the R caudate & R Heschl’s gyrus (standardised β = −0.350; p = 0.009), the L cerebellar cortex & R inferior colliculus (standardised β = −0.320; p = 0.017) and the R ventral diencephalon & R cochlear nucleus (standardised β = −0.284; p = 0.036) there were negative associations. Whereas for the connection between the L ventral diencephalon & L inferior colliculus, a positive relationship was observed (standardised β = 0.274; p = 0.045). None of these were tracts in which we had previously found differences in MD between iHEU and iHU.

When examining the MD-language relationship separately according to HIV exposure groups, for tracts in which we had previously found differences in MD between iHEU and iH (Supplementary figure 14), there was generally a negative relationship between MD and language scores for iHU. In particular, for five of these 22 tracts (Figure 6) the correlation was moderately strong in iHU (R < −0.5, p < 0.05). These tracts were between the medulla & L cochlear nucleus, the R thalamus & L MGN, the midbrain & L MGN, the L ventral diencephalon & L MGN and between the L caudate & L cochlear nucleus. Whereas no meaningful correlations between MD and language were observed for iHEU.

**Figure 6:**
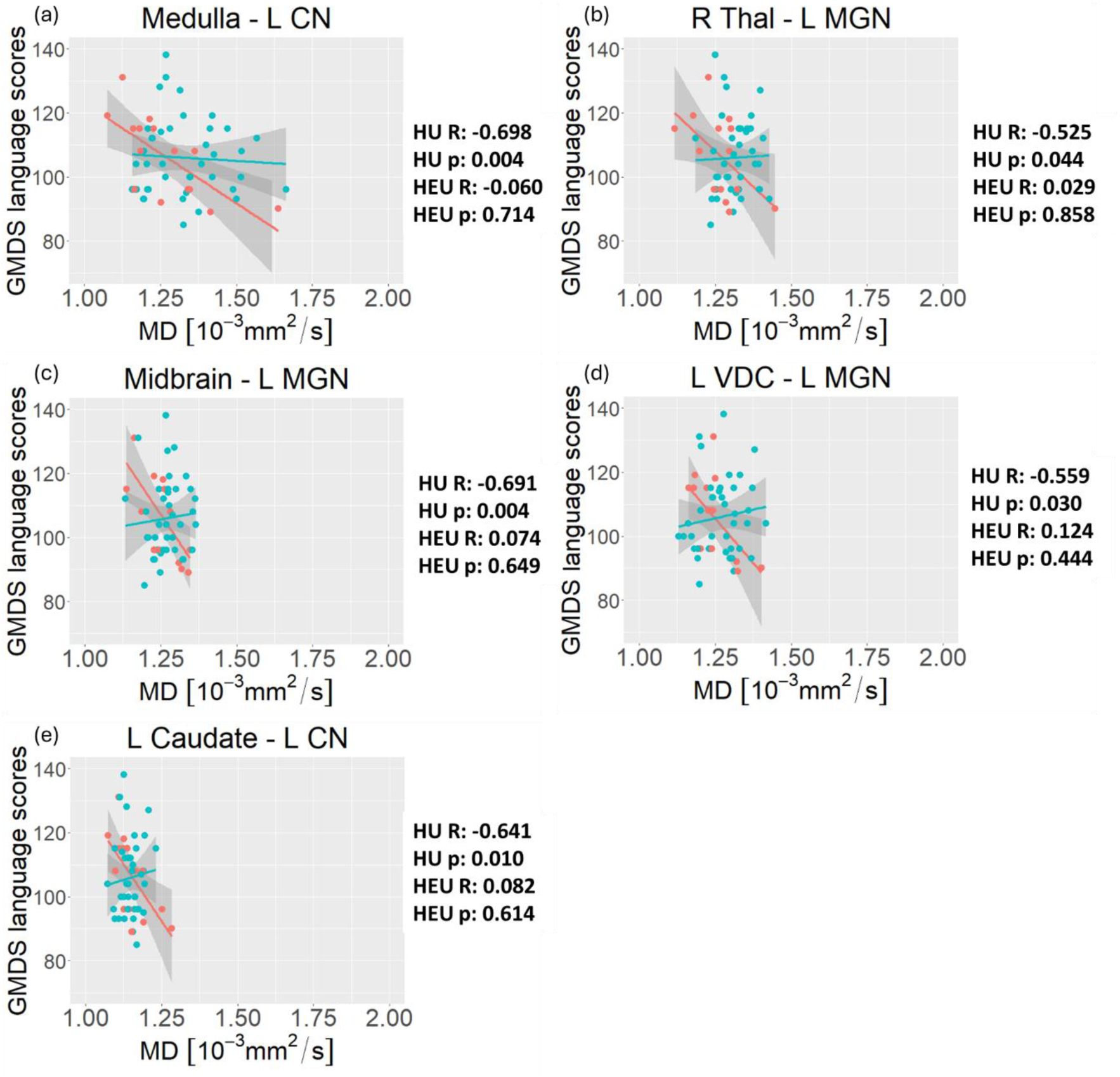
Tracts in which we observe negative correlations between mean diffusivity (MD) and language outcomes in iHU. Red=iHU; Blue =iHEU.

For the 30 tracts in which we had previously observed groupwise MD differences when comparing iHEU-pre and iHEU-post to iHU (Supplementary figure 15), we identified seven tracts in which iHU had negative correlations between MD and language performance at 9-14 months and for which the correlation coefficient was less than −0.5 (p <0.05). In each of these tracts there was no clear relationship between MD and language for either of the iHEU-pre and iHEU-post groups.

## Discussion

We explored the effects of *in utero* HIV and ART exposure, and thus development in the context of a compromised maternal immune system, on neonate auditory WM pathway development. Moreover, we investigated whether such effects were foretelling of language outcomes at 9-14 months. Our hypothesis was that there would be altered FA and MD in auditory WM tracts of iHEU compared to iHU. Yet we reported no HIV/ART exposure effects after multiple comparison correction.

### Methodology and effect sizes

While the effect sizes we observe are similar to those reported by Magondo et al. (2024) and Liu et al. (2012), our results are not as statistically robust. Methodological differences may contribute to the lack of observed group differences after multiple comparison correction compared to whole brain tractography work that has been carried out by Magondo et al. (2024) in this cohort. In contrast to the groupwise differences reported by Magondo et al. (2024), our findings may indicate that HIV/ART exposure has less influence on WM in the central auditory system than on other regions of the neonatal brain.

It has been shown that carrying out tractography to quantify DTI measures in connections to smaller structures can lead to a greater amount of variation in results ^62^. As we incorporated small auditory structures to examine the auditory system, a greater amount of variation in our results may contribute to findings that are not as statistically strong as those of Magondo et al. (2024). Moreover, we included connections to brainstem structures, which can be difficult to image ^63^ and may have led to greater variation. Based on possible methodological sources of variation, patterns in our unadjusted results and effects sizes we suggest that HIV exposure may have a subtle influence on select auditory tracts, particularly those connected to the left MGN.

### Delayed maturation of tracts connected to the auditory thalamus for all iHEU

When focusing on the patterns within our results, a reduction in FA typically suggests poor WM tract integrity or lower axonal density ^64–67^. Among our unadjusted results, we note two of the six connections had reduced FA between the MGN and cerebral cortex. In a previous whole brain tractography study in this cohort, lower FA was similarly reported in four tracts in both iHEU-pre and iHEU-post compared to HUU, with affected tracts connecting the left cerebral cortex to subcortical structures ^64^. Additionally, lower FA has been reported previously in WM tracts of HEU children, in regions including the inferior occipital gyrus, frontal gyrus and paracentral lobule ^25^.

Elevated MD, RD and AD, as seen in several auditory WM tracts when comparing iHEU in this study to iHU, is generally indicative of delayed WM maturation and myelination ^68^. Previous DTI studies that have compared children who are HEU to children who are HU have reported reduced ^23^ or unchanged ^24,26^ MD in WM tracts of iHEU. Although the study by Yadav et al. (2020) found higher MD in the R parietal region of children who are HEU, their remaining results showed reductions in MD in R middle temporal gyrus, R cerebellum crus and L extra nuclear regions.

Furthermore, the L MGN was the structure most frequently observed among the auditory connections for which iHEU as a group had MD differences in our unadjusted results. The MGN is a component of the thalamus referred to as the auditory thalamus ^69^. It connects the inferior colliculus to the primary auditory cortex and provides feedback to upstream auditory regions ^69,70^, also playing an important role in processing speech ^71–73^. Relatively few DTI studies have investigated the MGN and connections to this structure in infants and children. In 4-year-old children with sensorineural hearing loss (SNHL), reduced FA in the MGN has been found compared to control children ^74^.

There is no evidence of poorer integrity or delayed maturation in intra-auditory WM tracts when comparing iHEU to iHU. Rather, in our unadjusted results HIV/ART exposure only appears to impact the integrity of tracts which connect auditory to non-auditory structures of the brain. Overall, our results would indicate that HIV/ART exposure may delay maturation of WM pathways to/from the central auditory system.

While some of the sources of variation in our data cannot be reduced due to methodological limitations, a larger study focused on fewer tracts is needed to confirm if these differences can be independently reproduced. If verified, our findings complement a previous DTI tractography study in this cohort, albeit excluding auditory structures, which reported regional reduced FA and increased MD among iHEU with similar effect sizes ^57^.

### Auditory tract maturation delayed in both pre and post ART exposure groups

Our findings suggest that iHEU-post drive the differences in FA results seen in our overall comparison of iHEU and iHU. This corresponds with what was found in the recent study in the HBS cohort using whole brain tractography, which reported reductions in FA mainly in iHEU-post compared to controls ^57^.

Development of the peripheral auditory system begins during early pregnancy, whereas there is a delay in the establishment of central auditory connections - following which processing of auditory signals in the cortex can begin by approximately 25 weeks gestational age ^75^. As iHEU-post in the HBS were exposed to ART for an average of 24.82 weeks, this would indicate that their mothers generally started treatment prior to the window in which the central auditory system begins to function. However, the impacts of HIV - particularly on the mothers’ immune systems - would be expected to persist beyond the time at which treatment was initiated.

Reduced FA is often accompanied by elevated RD in iHEU-post. Based on the proposed biological models outlined by Dubois et al. (2008), this would imply that the WM developmental stage which is mainly being affected in this ART exposure group, is the initial arrangement of fibers. This phase of WM development is expected to occur *in utero* ^76,77^. Thus, the subtle effects of delayed ART treatment for auditory tract development may begin as early as the fetal development stages.

Furthermore, simultaneous elevations in MD and reductions in FA were observed for three tracts in unadjusted iHEU-post results. This is an interesting finding, as Dubois et al. (2008) defined the myelination of WM fibers as a developmental step in which increases in FA would be expected to occur in conjunction with reductions in MD and RD. Therefore, the combination of results we observe in these tracts for iHEU-post compared to iHU is characteristic of disruptions to the final step in WM tract development, postnatal myelination, described by Dubois et al. (2008).

Unlike the group-wise analysis of FA, for which only iHEU-post had altered connections, the overall differences in MD between iHEU and iHU appear to be driven by both iHEU-pre and iHEU-post for several auditory connections in our unadjusted results. In some cases, MD differences were driven by iHEU-pre alone or iHEU-post alone – thus, altered MD in iHEU compared to iHU is not obviously driven by one group. This differs from Magondo et al. (2024) whose findings suggested an effect due to longer ART exposure, as they only reported MD differences in iHEU-pre and not in iHEU-post.

Elevated MD is generally accompanied by elevated RD and/or AD in both ART exposure groups. This pattern in DTI measure differences is the opposite of what is expected with the rapid accumulation of supporting glial cells, the second WM tract development step as outlined by Dubois et al. (2008). As such, our results suggest that HIV exposure may delay the process of building up the cells required for the final developmental step of auditory tract development, namely myelination. Collectively our findings suggest that HIV exposure, more so than the duration of ART exposure, is responsible for the delays in WM tract maturation.

A closer examination of the connections for which MD differences were observed found that connections to the L MGN were affected in both iHEU-pre and iHEU-post groups. Meanwhile connections to the R MGN were found to have elevated MD mainly in iHEU-pre. Previous whole brain tractography in the HBS cohort, that used only seeds based on the infant FreeSurfer tool, found that iHEU-pre displayed elevated MD largely in tracts connected to the thalamus ^57^. Given that the MGN is part of the auditory thalamic region, our findings complement those presented by Magondo et al. (2024). However, our findings would suggest that connections to the L auditory thalamus are affected by exposure to HIV moreso than exposure to ART for a longer duration.

We report that poorer auditory tract integrity is driven by iHEU-post, while both ART exposure groups appear to have delayed maturation in auditory connections. Various factors could contribute to these differing results. In particular, the immune health of mothers might be expected to play a role in disrupting WM integrity in iHEU-post. As the target cells of HIV, CD4 counts provide a measure of progression of HIV infection ^78^. Thus, our findings of higher CD4 counts for mothers of iHEU-pre, would suggest that mothers who start treatment earlier have a slower progression of disease, corresponding with stronger immune health. However, our results provided no evidence of CD4 count influencing auditory WM tract development in this cohort.

### HIV and ART exposure effects on language development and the relationship between DTI measures & language

#### HIV and ART exposure do not influence language performance at 9-14 months

Overall, we found no differences in 9-14-month GMDS or Griffiths III language scores between iHEU and iHU, or when comparing iHEU-pre and iHEU-post to iHU. This is a positive prognosis for these infants, suggesting that the observed effects of HIV and ART exposure on newborn auditory WM tract integrity and maturation do not influence language development at 9-14 months. However, as previous studies have identified HIV exposure effects on language development in children ^8,9^, it is possible that it is too early to detect the developmental impacts in infants from the HBS, and these may still be observed with time.

#### HIV and ART exposure may affect the auditory tract development-language performance relationship

Visualising the relationships between language scores and DTI measures separately for iHEU and iHU for tracts in which we had observed group differences, revealed correlations in the iHU group. A positive association of moderate strength (r > 0.5) between FA and language, was observed in this group for two of the six tracts in which we had seen groupwise FA differences. These are tracts connected to the L & R CN. Previous research in children that have received cochlear implants to treat severe SNHL has found that increasing FA in the MGN corresponds with better outcomes in measures indicative of auditory processing and language^79^. In a small study of children with hearing loss in one ear, FA in the auditory cortex has been found to negatively correlate with the need for a more personalised teaching approach ^80^. Therefore, our FA-language findings complement other research, but identify the CN specifically.

Moderately strong negative associations between MD and language outcomes were observed for iHU in five (23%) of the tracts in which we had observed groupwise MD differences. Three of these tracts connected to the L MGN, and the other two to the L CN. Ultimately, our results indicate that for a small percentage of tracts greater integrity (with higher FA) and more established organization (with lower MD) ^31,81,82^ of the respective WM tracts, correlate with better performance in language assessments in iHU.

In iHEU, however, we did not find meaningful relationships between language outcomes and DTI measures across any of the affected tracts. Although groupwise differences in FA were only observed previously for the iHEU-post ART exposure group compared to iHU (Figure 2), neither iHEU-post nor iHEU-pre appeared to be specifically responsible for the absence of clear positive correlations between language outcomes and auditory tract integrity (determined by FA). The association between delayed auditory tract development, as seen by higher MD, and poorer language outcomes at 9-14 months in iHU, was also not observed in either iHEU-pre or iHEU-post. Collectively, these findings would imply that ART exposure is not the main factor influencing the relationships between auditory tract integrity or maturation at 0-5 weeks, and language performance at 9-14 months in our cohort.

Our results suggest the process of maturation in auditory tracts may differ for iHEU compared to iHU, despite both groups performing similarly in language assessments. In a recent study by McFayden et al. (2024), FA in the arcuate fasciculus was found to be associated with language in infants at higher risk for (and later diagnosed with) autism spectrum disorder^36^. Meanwhile, in the same study this association was not reported in low-risk infants. McFayden et al. (2024) suggested that they may only observe associations in high-risk infants diagnosed with autism due to how the arcuate fasciculate develops and becomes more rapidly established for use in language activity.

Previous research in infants would suggest that the timing and way in which tracts develop and mature may play an important functional role. A study by Sket et al. (2019) showed that FA in specific segments of the arcuate fasciculus corresponded with language outcomes, whereas FA for the tract as a whole did not. They attributed this finding to the fact that different segments of tracts do not all develop and mature at the same time. Similarly, Farah et al. (2020) showed that FA and MD in specific segments of WM tracts in children correlate with their language performance. This corresponds with studies in infants and children that have similarly shown that even within WM tracts, maturation is not uniform ^83,84^. Therefore, if there is disruption to the timing or sequence with which regions of tracts mature, this could influence how DTI measures in auditory tracts correlate with language performance.

### Strengths, limitations and future work

Previous research in infants and children has reported subtle neurodevelopmental changes in relation to HIV exposure. However, as these studies do not provide measures of effect size, a better understanding of the clinical relevance of these findings is needed. This study is strengthened by the inclusion of both p values and measures of effect size, thereby providing potential insight into the biological meaningfulness of our findings. We also present effect sizes based on data from two published studies, for comparison and to provide context.

Neonates are a challenging age group to study both in terms of acquiring motion-free images and due to the limitations of tools for automated brain image segmentation. However, studying these early days in postnatal development can provide a wealth of information about the impacts of *in utero* exposures with minimal environmental confounding factors.

The period within the first two weeks of life has been identified as a good window for studying the WM tracts of infants - as this is a period in which inter-subject WM metrics are relatively constant, and therefore variability in DTI measures such as FA, are minimal ^85^. Jin et al. (2019) have found that this period is shortly followed by rapid changes in WM.

This study is limited by our sample size, which was smaller than originally anticipated. As such, the statistical power to detect group differences was lower than expected and may have contributed to our lack of results following multiple comparison correction. Although some of our results were equivalent in effect size compared to other studies, our absence of adjusted results may also be due in part to the large number of tracts we explored. While our analysis included 106 tracts, Liu et al. (2012) only presented results for 15 tracts and thus their findings would have been less strongly penalized by multiple comparison correction. Yet as Magondo et al. (2024) studied 173 tracts and still reported group differences after multiple comparison correction, strict correction for multiple comparisons cannot fully account for our lack of adjusted results. However, future studies looking at WM connections to small structures in the infant brain should take this into consideration and could be more selective, focusing on a smaller number of tracts.

Although hearing tests were carried out for a small subset of infants identified as being at greater risk of hearing loss due to cytomegalovirus infection, hearing assessments were not conducted routinely as part of this study. Therefore, we do not have sufficient data in this study with which to assess the relationship between hearing outcomes and DTI measures in auditory WM tracts of iHEU. Other causes of hearing loss in infants, such as cytomegalovirus infection and nutritional deficits, could also be examined in future. As we lacked a group of iHU with cytomegalovirus, we were unable to control for this in our study. Furthermore, the number of iHEU with cytomegalovirus was too small to carry out meaningful statistical analysis.

Maternal HIV viral loads and CD4 counts were not collected as part of the HBS, but rather under standard care provided by the provincial government. The timing at which this data was collected did not always correspond with the times at which infants were scanned. This limits our narrative regarding whether the effects of HIV exposure are due to the virus itself or the effects of HIV on the maternal immune system.

Our analysis only allows us to examine FA and MD for entire tracts. To assess whether the absence of correlations between DTI measures and language in iHEU could be due to HIV/ART exposure-driven changes in the order or way with which auditory tracts develop, future work could do along-tract assessments.

We provide plots showing that the relationships between DTI measures and language appear to differ for HIV and ART exposure groups in a few tracts, implying that there may be interactions occurring here. However, statistical analysis of data from a larger cohort – in which interaction terms are introduced into regression models – would be required to verify this.

## Conclusion

We presented evidence, based on unadjusted results, that exposure to HIV and antiretrovirals may impact the central auditory system. Methodological limitations, such as small seed regions, may be responsible for the lack of statistically robust results. The differences we observe are of a similar effect size to other infant probabilistic tractography studies. Patterns demonstrating elevated MD across both HIV exposure groups indicate that HIV exposure specifically may affect maturity of tracts in the left hemisphere and those connected to the MGN. Moreover, reductions in FA driven by iHEU-post point to a possible harmful effect of late maternal treatment on the integrity of select auditory tracts.

Positive associations between auditory WM tract integrity and language performance, and negative associations between maturation and language, are observed in iHU only for a small proportion of tracts. Meanwhile, there is a lack of associations in iHEU. Together with the groupwise MD differences we observe, this suggests subtle alterations in the way in which auditory tracts mature in iHEU compared to iHU, albeit without detectably affecting language performance. Future studies in a larger cohort will be required to verify these findings and to assess the need for interventions in this population. Moreover, additional research will be required to determine the underlying mechanisms involved in HIV/ART exposure effects on central auditory tract development.

### Data and code availability

Tractography output will be made available upon request and enquiries about additional data can be made. However, raw imaging data may not be made available while it is still being used in ongoing research.

## Acknowledgements

We would like to acknowledge and thank Dr Samantha Fry, for all the clinical assessments she carried out for the HBS infants. Further, we would like to thank Dr Marlie Miles for carrying out the manual tracing of the auditory structures on the brain images of infants.

Sincere thanks to the radiographers at CUBIC, Groote Schuur Hospital, and to the mothers and infants who have participated in the HBS.

