## Supplementary material for "Auditory white matter tract development in infants exposed to HIV and antiretrovirals"

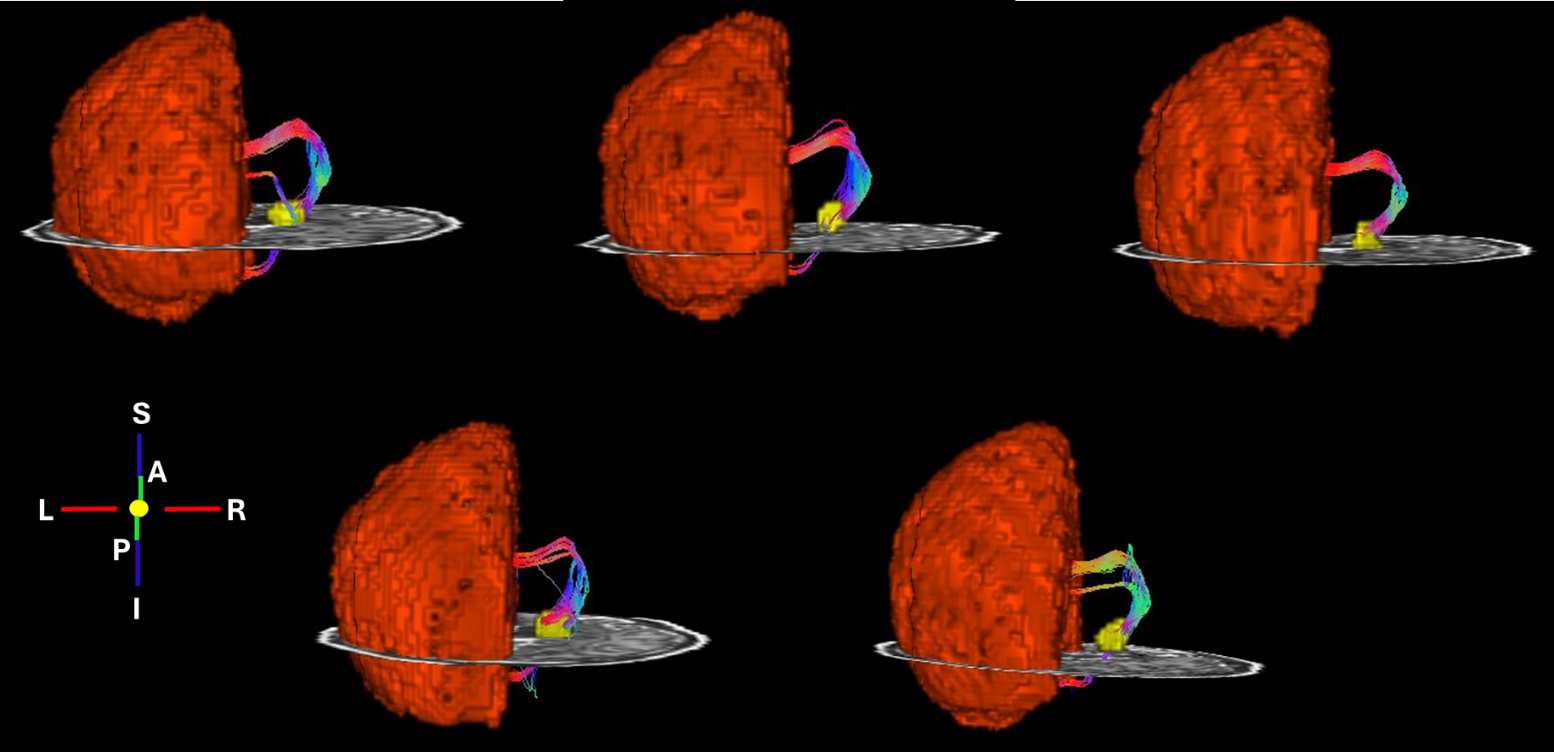

**Supplementary figure 1:** Figures showing the L CC-R MGN tract for five infants (2 males; 3 iHU and 2 iHEU (1 iHEU-pre and 1 iHEU-post)).

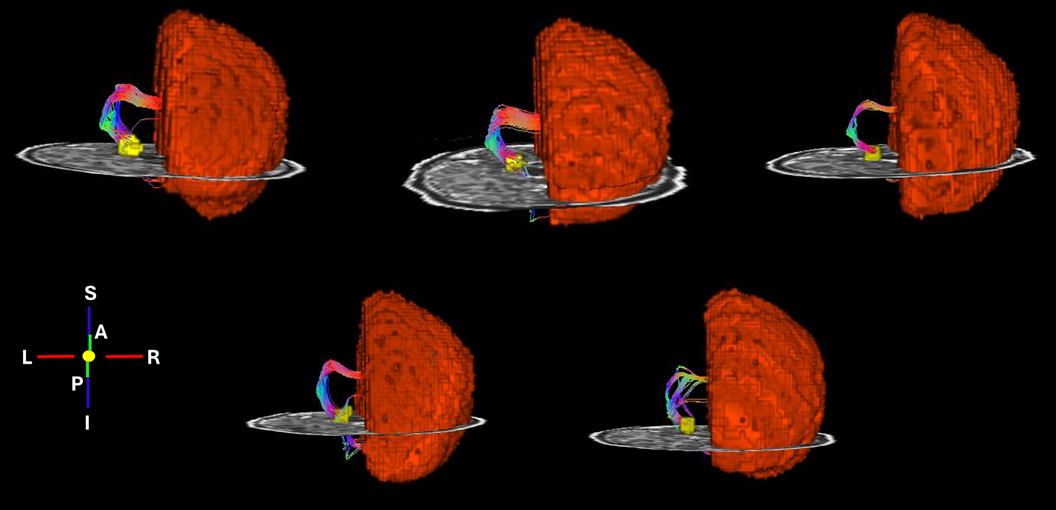

**Supplementary figure 2:** Figures showing the R CC-L MGN tract for five infants (2 males; 3 iHU and 2 iHEU (1 iHEU-pre and 1 iHEU-post)).

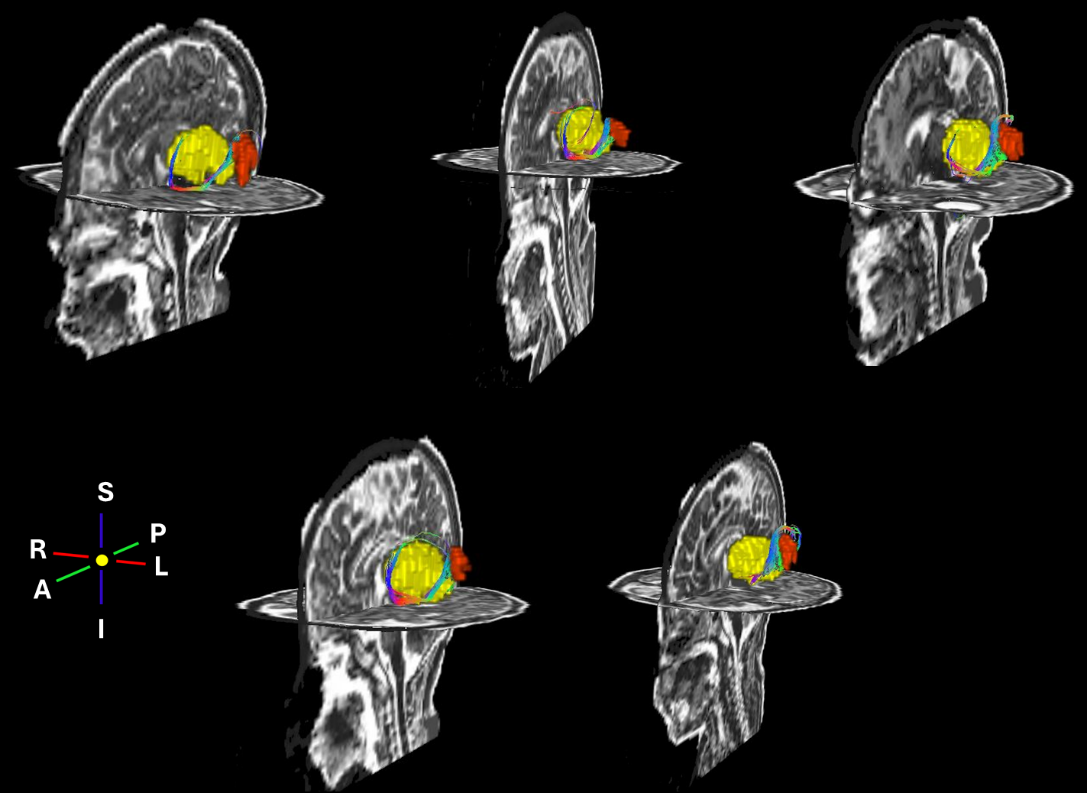

**Supplementary figure 3:** Figures showing the L Thal – L HG tract for five infants (2 males; 3 iHU and 2 iHEU (1 iHEU-pre and 1 iHEU-post)).

**Supplementary table 1:** A summary of means and standard deviations of FA, MD, RD and AD in iHU and iHEU for all auditory connections; results of linear regression comparing DTI measures between iHEU and iHU, showing standardised beta (Std β) and standardised standard error (Std SE), p values and p values adjusted for multiple comparisons (q values). For each predictor variable (HIV exposure group, maternal education and maternal weight) r and r^2^ values are provided.

| **DTI measure** | **Mean HU** | **sd HU** | **Mean HEU** | **sd HEU** | **Group % difference** | **Std β** | **Std SE** | **p value** | **q value** | **Lower CI** | **Upper CI** | **r HIV exp** | **r^2^ HIV exp** | **r mat ed** | **r^2^ mat ed** | **r mat weight** | **r^2^ mat weight** |
| --- | --- | --- | --- | --- | --- | --- | --- | --- | --- | --- | --- | --- | --- | --- | --- | --- | --- |
| L CC - L Inferior colliculus | | | | | | | | | | | | | | | | | |
| FA | 0.233 | 0.018 | 0.225 | 0.021 | -3.463 | -0.157 | 0.115 | 0.174 | 0.964 | -0.385 | 0.071 | -0.179 | 0.032 | 0.047 | 0.002 | 0.132 | 0.017 |
| MD | 1.321 | 0.063 | 1.367 | 0.057 | 3.488 | 0.293 | 0.109 | **0.009** | 0.233 | 0.076 | 0.510 | 0.340 | 0.115 | -0.116 | 0.013 | -0.258 | 0.067 |
| RD | 1.158 | 0.061 | 1.205 | 0.058 | 4.028 | 0.287 | 0.108 | **0.010** | 0.167 | 0.071 | 0.502 | 0.343 | 0.118 | -0.143 | 0.021 | -0.270 | 0.073 |
| AD | 1.648 | 0.076 | 1.693 | 0.065 | 2.729 | 0.266 | 0.112 | **0.020** | 0.294 | 0.043 | 0.490 | 0.290 | 0.084 | -0.053 | 0.003 | -0.203 | 0.041 |
| L CC - L Heschl's gyrus | | | | | | | | | | | | | | | | | |
| FA | 0.201 | 0.014 | 0.199 | 0.014 | -0.997 | 0.015 | 0.113 | 0.891 | 0.984 | -0.208 | 0.239 | -0.068 | 0.005 | 0.148 | 0.022 | 0.228 | 0.052 |
| MD | 1.306 | 0.053 | 1.341 | 0.048 | 2.653 | 0.255 | 0.109 | **0.021** | 0.206 | 0.039 | 0.471 | 0.306 | 0.093 | -0.100 | 0.010 | -0.260 | 0.067 |
| RD | 1.167 | 0.055 | 1.200 | 0.051 | 2.801 | 0.217 | 0.109 | **0.050** | 0.278 | 0.000 | 0.434 | 0.278 | 0.077 | -0.116 | 0.013 | -0.273 | 0.075 |
| AD | 1.584 | 0.056 | 1.623 | 0.047 | 2.434 | 0.312 | 0.109 | **0.005** | 0.275 | 0.096 | 0.529 | 0.337 | 0.114 | -0.059 | 0.004 | -0.210 | 0.044 |
| L CC - L Cochlear nucleus | | | | | | | | | | | | | | | | | |
| FA | 0.244 | 0.015 | 0.238 | 0.013 | -2.453 | -0.183 | 0.116 | 0.118 | 0.964 | -0.413 | 0.048 | -0.192 | 0.037 | -0.074 | 0.005 | 0.237 | 0.056 |
| MD | 1.239 | 0.063 | 1.269 | 0.063 | 2.389 | 0.217 | 0.114 | 0.062 | 0.233 | -0.011 | 0.444 | 0.211 | 0.045 | -0.020 | 0.000 | -0.055 | 0.003 |
| RD | 1.079 | 0.059 | 1.101 | 0.043 | 2.105 | 0.201 | 0.115 | 0.086 | 0.291 | -0.029 | 0.431 | 0.211 | 0.045 | 0.024 | 0.001 | -0.187 | 0.035 |
| AD | 1.570 | 0.071 | 1.590 | 0.067 | 1.319 | 0.157 | 0.116 | 0.178 | 0.523 | -0.073 | 0.387 | 0.139 | 0.019 | -0.004 | 0.000 | 0.019 | 0.000 |
| L CC - L Medial geniculate nucleus | | | | | | | | | | | | | | | | | |
| FA | 0.227 | 0.013 | 0.219 | 0.012 | -3.381 | -0.193 | 0.109 | 0.080 | 0.849 | -0.410 | 0.024 | -0.272 | 0.074 | 0.212 | 0.045 | 0.226 | 0.051 |
| MD | 1.318 | 0.052 | 1.349 | 0.049 | 2.423 | 0.255 | 0.112 | **0.025** | 0.206 | 0.033 | 0.478 | 0.283 | 0.080 | -0.043 | 0.002 | -0.219 | 0.048 |
| RD | 1.158 | 0.051 | 1.194 | 0.051 | 3.149 | 0.252 | 0.108 | **0.022** | 0.167 | 0.037 | 0.467 | 0.313 | 0.098 | -0.116 | 0.013 | -0.304 | 0.092 |
| AD | 1.648 | 0.078 | 1.671 | 0.066 | 1.421 | 0.161 | 0.114 | 0.162 | 0.518 | -0.066 | 0.388 | 0.153 | 0.023 | 0.076 | 0.006 | -0.136 | 0.018 |
| L CC - R Medial geniculate nucleus | | | | | | | | | | | | | | | | | |
| FA | 0.257 | 0.015 | 0.249 | 0.011 | -3.121 | -0.283 | 0.111 | **0.013** | 0.682 | -0.505 | -0.062 | -0.292 | 0.085 | 0.034 | 0.001 | 0.143 | 0.021 |
| MD | 1.325 | 0.061 | 1.354 | 0.054 | 2.138 | 0.255 | 0.113 | **0.027** | 0.206 | 0.030 | 0.481 | 0.228 | 0.052 | 0.034 | 0.001 | -0.031 | 0.001 |
| RD | 1.139 | 0.056 | 1.171 | 0.049 | 2.765 | 0.294 | 0.112 | **0.010** | 0.167 | 0.071 | 0.517 | 0.272 | 0.074 | 0.021 | 0.000 | -0.064 | 0.004 |
| AD | 1.698 | 0.077 | 1.720 | 0.067 | 1.297 | 0.178 | 0.115 | 0.126 | 0.477 | -0.051 | 0.407 | 0.143 | 0.020 | 0.052 | 0.003 | 0.022 | 0.001 |
| L Lat Vent - L Inferior colliculus | | | | | | | | | | | | | | | | | |
| FA | 0.242 | 0.018 | 0.242 | 0.017 | 0.187 | 0.121 | 0.110 | 0.277 | 0.964 | -0.099 | 0.341 | 0.012 | 0.000 | 0.265 | 0.070 | 0.155 | 0.024 |
| MD | 1.350 | 0.097 | 1.379 | 0.100 | 2.165 | 0.108 | 0.115 | 0.347 | 0.565 | -0.119 | 0.336 | 0.135 | 0.018 | -0.009 | 0.000 | -0.178 | 0.032 |
| RD | 1.177 | 0.089 | 1.203 | 0.094 | 2.186 | 0.083 | 0.114 | 0.469 | 0.681 | -0.144 | 0.310 | 0.127 | 0.016 | -0.054 | 0.003 | -0.200 | 0.040 |
| AD | 1.696 | 0.118 | 1.732 | 0.119 | 2.135 | 0.143 | 0.115 | 0.217 | 0.558 | -0.085 | 0.371 | 0.139 | 0.019 | 0.062 | 0.004 | -0.135 | 0.018 |
| L Lat Vent - L Heschl's gyrus | | | | | | | | | | | | | | | | | |
| FA | 0.250 | 0.018 | 0.243 | 0.021 | -2.848 | -0.072 | 0.111 | 0.517 | 0.964 | -0.293 | 0.148 | -0.159 | 0.025 | 0.180 | 0.032 | 0.251 | 0.063 |
| MD | 1.364 | 0.075 | 1.419 | 0.090 | 3.969 | 0.288 | 0.112 | **0.012** | 0.206 | 0.065 | 0.511 | 0.278 | 0.078 | -0.003 | 0.000 | -0.092 | 0.008 |
| RD | 1.181 | 0.067 | 1.235 | 0.089 | 4.618 | 0.280 | 0.112 | **0.014** | 0.167 | 0.058 | 0.502 | 0.288 | 0.083 | -0.038 | 0.001 | -0.132 | 0.018 |
| AD | 1.732 | 0.100 | 1.786 | 0.100 | 3.083 | 0.278 | 0.113 | **0.016** | 0.294 | 0.054 | 0.502 | 0.239 | 0.057 | 0.057 | 0.003 | -0.015 | 0.000 |
| L Lat Vent - L Medial geniculate nucleus | | | | | | | | | | | | | | | | | |
| FA | 0.252 | 0.019 | 0.244 | 0.020 | -3.079 | -0.108 | 0.112 | 0.338 | 0.964 | -0.331 | 0.115 | -0.180 | 0.033 | 0.191 | 0.036 | 0.180 | 0.032 |
| MD | 1.354 | 0.091 | 1.407 | 0.087 | 3.921 | 0.281 | 0.111 | **0.014** | 0.206 | 0.059 | 0.502 | 0.268 | 0.072 | 0.057 | 0.003 | -0.149 | 0.022 |
| RD | 1.170 | 0.083 | 1.224 | 0.084 | 4.589 | 0.281 | 0.111 | **0.013** | 0.167 | 0.060 | 0.502 | 0.283 | 0.080 | 0.017 | 0.000 | -0.175 | 0.030 |
| AD | 1.720 | 0.117 | 1.772 | 0.100 | 3.014 | 0.258 | 0.112 | **0.024** | 0.294 | 0.035 | 0.481 | 0.222 | 0.049 | 0.118 | 0.014 | -0.096 | 0.009 |
| L Lat Vent - R Medial geniculate nucleus | | | | | | | | | | | | | | | | | |
| FA | 0.277 | 0.019 | 0.273 | 0.018 | -1.181 | 0.028 | 0.110 | 0.802 | 0.974 | -0.191 | 0.246 | -0.082 | 0.007 | 0.230 | 0.053 | 0.252 | 0.064 |
| MD | 1.410 | 0.090 | 1.451 | 0.082 | 2.916 | 0.238 | 0.114 | **0.040** | 0.210 | 0.011 | 0.465 | 0.219 | 0.048 | 0.019 | 0.000 | -0.046 | 0.002 |
| RD | 1.196 | 0.082 | 1.235 | 0.076 | 3.263 | 0.222 | 0.114 | 0.054 | 0.278 | -0.004 | 0.449 | 0.225 | 0.050 | -0.024 | 0.001 | -0.094 | 0.009 |
| AD | 1.836 | 0.113 | 1.876 | 0.097 | 2.215 | 0.220 | 0.116 | 0.061 | 0.447 | -0.010 | 0.450 | 0.182 | 0.033 | 0.053 | 0.003 | 0.002 | 0.000 |
| L Cb - L Inferior colliculus | | | | | | | | | | | | | | | | | |
| FA | 0.178 | 0.016 | 0.174 | 0.014 | -2.587 | -0.095 | 0.114 | 0.407 | 0.964 | -0.323 | 0.132 | -0.144 | 0.021 | 0.121 | 0.015 | 0.139 | 0.019 |
| MD | 1.216 | 0.083 | 1.229 | 0.071 | 1.079 | 0.078 | 0.117 | 0.505 | 0.741 | -0.154 | 0.310 | 0.081 | 0.007 | -0.015 | 0.000 | -0.035 | 0.001 |
| RD | 1.104 | 0.082 | 1.119 | 0.068 | 1.361 | 0.085 | 0.116 | 0.467 | 0.681 | -0.146 | 0.317 | 0.095 | 0.009 | -0.032 | 0.001 | -0.052 | 0.003 |
| AD | 1.439 | 0.088 | 1.449 | 0.080 | 0.647 | 0.061 | 0.117 | 0.600 | 0.826 | -0.171 | 0.294 | 0.052 | 0.003 | 0.016 | 0.000 | -0.003 | 0.000 |
| L Cb - L Cochlear nucleus | | | | | | | | | | | | | | | | | |
| FA | 0.175 | 0.017 | 0.173 | 0.019 | -1.230 | -0.032 | 0.116 | 0.781 | 0.974 | -0.262 | 0.198 | -0.054 | 0.003 | -0.018 | 0.000 | 0.150 | 0.022 |
| MD | 1.259 | 0.092 | 1.278 | 0.091 | 1.544 | 0.088 | 0.116 | 0.448 | 0.679 | -0.143 | 0.319 | 0.097 | 0.009 | 0.003 | 0.000 | -0.095 | 0.009 |
| RD | 1.146 | 0.091 | 1.166 | 0.091 | 1.745 | 0.091 | 0.116 | 0.436 | 0.681 | -0.140 | 0.321 | 0.101 | 0.010 | 0.011 | 0.000 | -0.110 | 0.012 |
| AD | 1.485 | 0.098 | 1.503 | 0.096 | 1.235 | 0.080 | 0.116 | 0.493 | 0.747 | -0.152 | 0.312 | 0.087 | 0.008 | -0.011 | 0.000 | -0.062 | 0.004 |
| L Cb - L Medial geniculate nucleus | | | | | | | | | | | | | | | | | |
| FA | 0.201 | 0.023 | 0.199 | 0.019 | -1.226 | -0.012 | 0.116 | 0.915 | 0.986 | -0.243 | 0.218 | -0.056 | 0.003 | 0.084 | 0.007 | 0.121 | 0.015 |
| MD | 1.184 | 0.065 | 1.186 | 0.048 | 0.218 | 0.053 | 0.116 | 0.650 | 0.822 | -0.178 | 0.284 | 0.022 | 0.000 | 0.115 | 0.013 | -0.017 | 0.000 |
| RD | 1.057 | 0.063 | 1.061 | 0.045 | 0.368 | 0.052 | 0.116 | 0.656 | 0.812 | -0.179 | 0.283 | 0.035 | 0.001 | 0.088 | 0.008 | -0.054 | 0.003 |
| AD | 1.437 | 0.081 | 1.437 | 0.068 | -0.003 | 0.045 | 0.116 | 0.700 | 0.911 | -0.186 | 0.275 | 0.000 | 0.000 | 0.132 | 0.018 | 0.038 | 0.001 |
| L Cb - R Inferior colliculus | | | | | | | | | | | | | | | | | |
| FA | 0.197 | 0.022 | 0.194 | 0.019 | -1.648 | -0.073 | 0.116 | 0.532 | 0.964 | -0.305 | 0.158 | -0.074 | 0.006 | -0.023 | 0.001 | 0.073 | 0.005 |
| MD | 1.195 | 0.084 | 1.199 | 0.053 | 0.336 | 0.049 | 0.117 | 0.675 | 0.822 | -0.183 | 0.281 | 0.029 | 0.001 | 0.045 | 0.002 | 0.020 | 0.000 |
| RD | 1.070 | 0.077 | 1.075 | 0.047 | 0.491 | 0.063 | 0.117 | 0.592 | 0.775 | -0.169 | 0.295 | 0.042 | 0.002 | 0.055 | 0.003 | 0.003 | 0.000 |
| AD | 1.445 | 0.108 | 1.447 | 0.076 | 0.106 | 0.025 | 0.117 | 0.830 | 0.956 | -0.207 | 0.258 | 0.008 | 0.000 | 0.027 | 0.001 | 0.039 | 0.002 |
| L Cb - R Medial geniculate nucleus | | | | | | | | | | | | | | | | | |
| FA | 0.221 | 0.022 | 0.218 | 0.020 | -1.525 | -0.074 | 0.117 | 0.528 | 0.964 | -0.306 | 0.158 | -0.075 | 0.006 | 0.011 | 0.000 | 0.026 | 0.001 |
| MD | 1.168 | 0.066 | 1.199 | 0.050 | 2.584 | 0.195 | 0.112 | 0.084 | 0.270 | -0.027 | 0.417 | 0.244 | 0.060 | -0.107 | 0.012 | -0.213 | 0.045 |
| RD | 1.032 | 0.053 | 1.057 | 0.041 | 2.397 | 0.197 | 0.112 | 0.081 | 0.285 | -0.025 | 0.419 | 0.243 | 0.059 | -0.095 | 0.009 | -0.219 | 0.048 |
| AD | 1.450 | 0.095 | 1.482 | 0.079 | 2.249 | 0.137 | 0.114 | 0.232 | 0.558 | -0.089 | 0.364 | 0.176 | 0.031 | -0.080 | 0.006 | -0.165 | 0.027 |
| L Thal - L Inferior colliculus | | | | | | | | | | | | | | | | | |
| FA | 0.222 | 0.014 | 0.223 | 0.020 | 0.847 | 0.067 | 0.116 | 0.564 | 0.964 | -0.164 | 0.298 | 0.047 | 0.002 | 0.092 | 0.008 | -0.049 | 0.002 |
| MD | 1.217 | 0.067 | 1.217 | 0.056 | 0.057 | 0.028 | 0.117 | 0.812 | 0.927 | -0.204 | 0.260 | 0.005 | 0.000 | 0.059 | 0.003 | 0.025 | 0.001 |
| RD | 1.076 | 0.063 | 1.075 | 0.051 | -0.113 | 0.005 | 0.117 | 0.968 | 0.989 | -0.228 | 0.237 | -0.010 | 0.000 | 0.035 | 0.001 | 0.029 | 0.001 |
| AD | 1.498 | 0.082 | 1.502 | 0.075 | 0.300 | 0.057 | 0.116 | 0.623 | 0.838 | -0.174 | 0.289 | 0.027 | 0.001 | 0.086 | 0.007 | 0.017 | 0.000 |
| L Thal - L Heschl's gyrus | | | | | | | | | | | | | | | | | |
| FA | 0.250 | 0.013 | 0.241 | 0.013 | -3.606 | -0.217 | 0.107 | **0.046** | 0.849 | -0.431 | -0.004 | -0.304 | 0.092 | 0.268 | 0.072 | 0.209 | 0.043 |
| MD | 1.235 | 0.054 | 1.273 | 0.066 | 3.036 | 0.259 | 0.112 | **0.023** | 0.206 | 0.036 | 0.482 | 0.267 | 0.071 | -0.014 | 0.000 | -0.154 | 0.024 |
| RD | 1.068 | 0.049 | 1.108 | 0.061 | 3.748 | 0.285 | 0.110 | **0.012** | 0.167 | 0.065 | 0.505 | 0.306 | 0.094 | -0.057 | 0.003 | -0.181 | 0.033 |
| AD | 1.570 | 0.069 | 1.603 | 0.081 | 2.067 | 0.203 | 0.114 | 0.079 | 0.447 | -0.024 | 0.430 | 0.189 | 0.036 | 0.053 | 0.003 | -0.102 | 0.010 |
| L Thal - L Cochlear nucleus | | | | | | | | | | | | | | | | | |
| FA | 0.230 | 0.015 | 0.231 | 0.022 | 0.630 | 0.036 | 0.117 | 0.760 | 0.974 | -0.197 | 0.268 | 0.033 | 0.001 | 0.016 | 0.000 | -0.024 | 0.001 |
| MD | 1.132 | 0.045 | 1.151 | 0.040 | 1.634 | 0.247 | 0.113 | **0.033** | 0.206 | 0.021 | 0.472 | 0.202 | 0.041 | 0.079 | 0.006 | 0.003 | 0.000 |
| RD | 0.994 | 0.042 | 1.008 | 0.041 | 1.451 | 0.198 | 0.115 | 0.088 | 0.291 | -0.030 | 0.426 | 0.159 | 0.025 | 0.069 | 0.005 | 0.010 | 0.000 |
| AD | 1.409 | 0.059 | 1.435 | 0.055 | 1.893 | 0.256 | 0.113 | **0.026** | 0.294 | 0.031 | 0.481 | 0.214 | 0.046 | 0.073 | 0.005 | -0.007 | 0.000 |
| L Thal - L Medial geniculate nucleus | | | | | | | | | | | | | | | | | |
| FA | 0.234 | 0.017 | 0.230 | 0.017 | -1.702 | -0.019 | 0.112 | 0.864 | 0.974 | -0.242 | 0.204 | -0.106 | 0.011 | 0.253 | 0.064 | 0.131 | 0.017 |
| MD | 1.212 | 0.051 | 1.239 | 0.051 | 2.224 | 0.270 | 0.112 | **0.018** | 0.206 | 0.047 | 0.493 | 0.236 | 0.055 | 0.110 | 0.012 | -0.087 | 0.008 |
| RD | 1.060 | 0.043 | 1.087 | 0.048 | 2.567 | 0.267 | 0.112 | **0.020** | 0.167 | 0.043 | 0.490 | 0.258 | 0.066 | 0.036 | 0.001 | -0.128 | 0.016 |
| AD | 1.518 | 0.076 | 1.544 | 0.069 | 1.745 | 0.232 | 0.112 | **0.041** | 0.378 | 0.009 | 0.455 | 0.169 | 0.028 | 0.192 | 0.037 | -0.017 | 0.000 |
| L Thal - R Inferior colliculus | | | | | | | | | | | | | | | | | |
| FA | 0.244 | 0.027 | 0.249 | 0.027 | 1.959 | 0.103 | 0.116 | 0.377 | 0.964 | -0.127 | 0.333 | 0.082 | 0.007 | 0.094 | 0.009 | -0.068 | 0.005 |
| MD | 1.232 | 0.083 | 1.242 | 0.074 | 0.825 | 0.062 | 0.117 | 0.596 | 0.800 | -0.170 | 0.294 | 0.061 | 0.004 | 0.013 | 0.000 | -0.044 | 0.002 |
| RD | 1.068 | 0.075 | 1.074 | 0.058 | 0.498 | 0.034 | 0.117 | 0.773 | 0.907 | -0.199 | 0.267 | 0.039 | 0.002 | -0.013 | 0.000 | -0.025 | 0.001 |
| AD | 1.560 | 0.113 | 1.580 | 0.119 | 1.271 | 0.085 | 0.116 | 0.468 | 0.719 | -0.147 | 0.316 | 0.078 | 0.006 | 0.040 | 0.002 | -0.058 | 0.003 |
| L Thal - R Medial geniculate nucleus | | | | | | | | | | | | | | | | | |
| FA | 0.263 | 0.019 | 0.265 | 0.015 | 0.783 | 0.119 | 0.115 | 0.302 | 0.964 | -0.109 | 0.347 | 0.057 | 0.003 | 0.134 | 0.018 | 0.084 | 0.007 |
| MD | 1.267 | 0.067 | 1.283 | 0.055 | 1.309 | 0.142 | 0.116 | 0.222 | 0.441 | -0.088 | 0.373 | 0.130 | 0.017 | 0.028 | 0.001 | -0.044 | 0.002 |
| RD | 1.084 | 0.054 | 1.097 | 0.051 | 1.207 | 0.112 | 0.116 | 0.337 | 0.586 | -0.119 | 0.343 | 0.115 | 0.013 | -0.003 | 0.000 | -0.070 | 0.005 |
| AD | 1.633 | 0.099 | 1.656 | 0.069 | 1.444 | 0.168 | 0.115 | 0.149 | 0.518 | -0.061 | 0.397 | 0.136 | 0.019 | 0.066 | 0.004 | -0.006 | 0.000 |
| L Caud - L Inferior colliculus | | | | | | | | | | | | | | | | | |
| FA | 0.226 | 0.020 | 0.228 | 0.020 | 0.728 | 0.022 | 0.117 | 0.851 | 0.974 | -0.210 | 0.254 | 0.038 | 0.001 | -0.053 | 0.003 | -0.023 | 0.001 |
| MD | 1.190 | 0.090 | 1.194 | 0.078 | 0.311 | 0.004 | 0.115 | 0.976 | 0.995 | -0.226 | 0.233 | 0.021 | 0.000 | 0.049 | 0.002 | -0.155 | 0.024 |
| RD | 1.047 | 0.082 | 1.049 | 0.075 | 0.183 | -0.003 | 0.116 | 0.979 | 0.989 | -0.233 | 0.227 | 0.011 | 0.000 | 0.056 | 0.003 | -0.145 | 0.021 |
| AD | 1.476 | 0.112 | 1.483 | 0.089 | 0.492 | 0.014 | 0.115 | 0.904 | 0.960 | -0.216 | 0.244 | 0.035 | 0.001 | 0.035 | 0.001 | -0.160 | 0.025 |
| L Caud - L Heschl's gyrus | | | | | | | | | | | | | | | | | |
| FA | 0.235 | 0.014 | 0.235 | 0.016 | -0.139 | 0.025 | 0.116 | 0.830 | 0.974 | -0.206 | 0.257 | -0.010 | 0.000 | 0.097 | 0.009 | 0.042 | 0.002 |
| MD | 1.209 | 0.042 | 1.229 | 0.063 | 1.600 | 0.133 | 0.115 | 0.253 | 0.470 | -0.097 | 0.362 | 0.153 | 0.023 | -0.054 | 0.003 | -0.102 | 0.010 |
| RD | 1.055 | 0.038 | 1.073 | 0.060 | 1.703 | 0.124 | 0.115 | 0.286 | 0.542 | -0.106 | 0.353 | 0.150 | 0.023 | -0.072 | 0.005 | -0.105 | 0.011 |
| AD | 1.517 | 0.058 | 1.540 | 0.074 | 1.455 | 0.135 | 0.116 | 0.247 | 0.558 | -0.095 | 0.365 | 0.144 | 0.021 | -0.022 | 0.000 | -0.089 | 0.008 |
| L Caud - L Cochlear nucleus | | | | | | | | | | | | | | | | | |
| FA | 0.238 | 0.018 | 0.234 | 0.015 | -1.873 | -0.153 | 0.116 | 0.190 | 0.964 | -0.383 | 0.077 | -0.128 | 0.016 | -0.058 | 0.003 | 0.024 | 0.001 |
| MD | 1.135 | 0.040 | 1.157 | 0.037 | 1.914 | 0.242 | 0.115 | **0.038** | 0.210 | 0.014 | 0.470 | 0.252 | 0.064 | -0.092 | 0.008 | -0.049 | 0.002 |
| RD | 0.995 | 0.055 | 1.010 | 0.035 | 1.567 | 0.167 | 0.115 | 0.152 | 0.394 | -0.063 | 0.396 | 0.170 | 0.029 | -0.074 | 0.005 | -0.008 | 0.000 |
| AD | 1.437 | 0.066 | 1.450 | 0.050 | 0.916 | 0.087 | 0.116 | 0.452 | 0.705 | -0.143 | 0.318 | 0.110 | 0.012 | -0.127 | 0.016 | 0.004 | 0.000 |
| L Caud - L Medial geniculate nucleus | | | | | | | | | | | | | | | | | |
| FA | 0.226 | 0.018 | 0.225 | 0.016 | -0.557 | 0.017 | 0.115 | 0.884 | 0.984 | -0.212 | 0.246 | -0.036 | 0.001 | 0.082 | 0.007 | 0.156 | 0.024 |
| MD | 1.192 | 0.053 | 1.222 | 0.051 | 2.545 | 0.285 | 0.112 | **0.013** | 0.206 | 0.062 | 0.507 | 0.260 | 0.068 | 0.091 | 0.008 | -0.117 | 0.014 |
| RD | 1.047 | 0.048 | 1.075 | 0.049 | 2.646 | 0.267 | 0.112 | **0.020** | 0.167 | 0.044 | 0.490 | 0.254 | 0.065 | 0.072 | 0.005 | -0.151 | 0.023 |
| AD | 1.482 | 0.072 | 1.518 | 0.065 | 2.402 | 0.275 | 0.113 | **0.017** | 0.294 | 0.051 | 0.499 | 0.237 | 0.056 | 0.106 | 0.011 | -0.053 | 0.003 |
| L Put - L Heschl's gyrus | | | | | | | | | | | | | | | | | |
| FA | 0.242 | 0.015 | 0.242 | 0.016 | -0.302 | 0.039 | 0.115 | 0.736 | 0.974 | -0.190 | 0.268 | -0.021 | 0.000 | 0.112 | 0.013 | 0.143 | 0.020 |
| MD | 1.188 | 0.043 | 1.207 | 0.042 | 1.613 | 0.183 | 0.114 | 0.112 | 0.296 | -0.044 | 0.409 | 0.204 | 0.041 | -0.039 | 0.002 | -0.148 | 0.022 |
| RD | 1.033 | 0.043 | 1.051 | 0.040 | 1.700 | 0.158 | 0.113 | 0.166 | 0.413 | -0.067 | 0.384 | 0.196 | 0.039 | -0.070 | 0.005 | -0.184 | 0.034 |
| AD | 1.497 | 0.054 | 1.520 | 0.059 | 1.493 | 0.184 | 0.115 | 0.112 | 0.477 | -0.044 | 0.413 | 0.177 | 0.031 | 0.012 | 0.000 | -0.071 | 0.005 |
| L Put - L Medial geniculate nucleus | | | | | | | | | | | | | | | | | |
| FA | 0.270 | 0.018 | 0.262 | 0.022 | -3.007 | -0.097 | 0.111 | 0.384 | 0.964 | -0.319 | 0.124 | -0.178 | 0.032 | 0.185 | 0.034 | 0.227 | 0.052 |
| MD | 1.187 | 0.076 | 1.203 | 0.060 | 1.318 | 0.149 | 0.115 | 0.201 | 0.427 | -0.081 | 0.378 | 0.110 | 0.012 | 0.080 | 0.006 | 0.019 | 0.000 |
| RD | 1.011 | 0.072 | 1.031 | 0.059 | 1.979 | 0.159 | 0.116 | 0.173 | 0.413 | -0.071 | 0.389 | 0.145 | 0.021 | 0.025 | 0.001 | -0.043 | 0.002 |
| AD | 1.540 | 0.091 | 1.547 | 0.075 | 0.450 | 0.113 | 0.114 | 0.327 | 0.584 | -0.114 | 0.340 | 0.040 | 0.002 | 0.156 | 0.024 | 0.114 | 0.013 |
| L Put - R Medial geniculate nucleus | | | | | | | | | | | | | | | | | |
| FA | 0.293 | 0.018 | 0.287 | 0.022 | -2.165 | -0.091 | 0.114 | 0.424 | 0.964 | -0.317 | 0.135 | -0.136 | 0.019 | 0.221 | 0.049 | -0.008 | 0.000 |
| MD | 1.230 | 0.073 | 1.236 | 0.063 | 0.523 | 0.055 | 0.117 | 0.642 | 0.822 | -0.178 | 0.287 | 0.045 | 0.002 | -0.019 | 0.000 | 0.048 | 0.002 |
| RD | 1.028 | 0.063 | 1.039 | 0.057 | 1.032 | 0.080 | 0.116 | 0.493 | 0.695 | -0.151 | 0.311 | 0.083 | 0.007 | -0.081 | 0.007 | 0.048 | 0.002 |
| AD | 1.633 | 0.100 | 1.631 | 0.090 | -0.120 | 0.015 | 0.117 | 0.896 | 0.960 | -0.217 | 0.248 | -0.010 | 0.000 | 0.061 | 0.004 | 0.042 | 0.002 |
| L Pal - L Cochlear nucleus | | | | | | | | | | | | | | | | | |
| FA | 0.239 | 0.019 | 0.238 | 0.019 | -0.793 | -0.024 | 0.116 | 0.839 | 0.974 | -0.255 | 0.208 | -0.046 | 0.002 | 0.009 | 0.000 | 0.113 | 0.013 |
| MD | 1.144 | 0.053 | 1.167 | 0.054 | 2.011 | 0.136 | 0.112 | 0.228 | 0.441 | -0.087 | 0.360 | 0.194 | 0.038 | -0.243 | 0.059 | -0.053 | 0.003 |
| RD | 0.998 | 0.051 | 1.019 | 0.049 | 2.113 | 0.131 | 0.113 | 0.249 | 0.489 | -0.093 | 0.355 | 0.191 | 0.037 | -0.228 | 0.052 | -0.082 | 0.007 |
| AD | 1.436 | 0.065 | 1.462 | 0.073 | 1.869 | 0.125 | 0.113 | 0.272 | 0.558 | -0.100 | 0.350 | 0.172 | 0.029 | -0.229 | 0.053 | -0.005 | 0.000 |
| 3rd Vent - L Inferior colliculus | | | | | | | | | | | | | | | | | |
| FA | 0.168 | 0.030 | 0.162 | 0.026 | -3.244 | -0.008 | 0.110 | 0.940 | 0.986 | -0.227 | 0.211 | -0.092 | 0.008 | 0.339 | 0.115 | -0.004 | 0.000 |
| MD | 1.361 | 0.156 | 1.436 | 0.138 | 5.513 | 0.189 | 0.112 | 0.096 | 0.274 | -0.034 | 0.412 | 0.234 | 0.055 | -0.222 | 0.050 | -0.041 | 0.002 |
| RD | 1.244 | 0.151 | 1.316 | 0.135 | 5.823 | 0.177 | 0.111 | 0.115 | 0.339 | -0.044 | 0.398 | 0.232 | 0.054 | -0.256 | 0.066 | -0.041 | 0.002 |
| AD | 1.596 | 0.178 | 1.677 | 0.152 | 5.030 | 0.200 | 0.113 | 0.082 | 0.447 | -0.026 | 0.426 | 0.226 | 0.051 | -0.152 | 0.023 | -0.038 | 0.001 |
| 4th Vent - L Inferior colliculus | | | | | | | | | | | | | | | | | |
| FA | 0.183 | 0.014 | 0.181 | 0.023 | -0.854 | 0.031 | 0.114 | 0.786 | 0.974 | -0.197 | 0.259 | -0.034 | 0.001 | 0.115 | 0.013 | 0.170 | 0.029 |
| MD | 1.337 | 0.126 | 1.373 | 0.148 | 2.668 | 0.100 | 0.116 | 0.393 | 0.631 | -0.131 | 0.330 | 0.115 | 0.013 | -0.020 | 0.000 | -0.104 | 0.011 |
| RD | 1.212 | 0.116 | 1.244 | 0.140 | 2.686 | 0.089 | 0.116 | 0.445 | 0.681 | -0.141 | 0.319 | 0.111 | 0.012 | -0.030 | 0.001 | -0.122 | 0.015 |
| AD | 1.588 | 0.149 | 1.630 | 0.170 | 2.640 | 0.114 | 0.116 | 0.330 | 0.584 | -0.117 | 0.345 | 0.117 | 0.014 | -0.002 | 0.000 | -0.071 | 0.005 |
| 4th Vent - L Cochlear nucleus | | | | | | | | | | | | | | | | | |
| FA | 0.188 | 0.019 | 0.177 | 0.023 | -5.981 | -0.240 | 0.118 | **0.045** | 0.849 | -0.474 | -0.006 | -0.234 | 0.055 | 0.018 | 0.000 | 0.090 | 0.008 |
| MD | 1.372 | 0.128 | 1.464 | 0.198 | 6.734 | 0.216 | 0.114 | 0.061 | 0.233 | -0.010 | 0.442 | 0.229 | 0.053 | -0.053 | 0.003 | -0.111 | 0.012 |
| RD | 1.238 | 0.120 | 1.330 | 0.195 | 7.471 | 0.222 | 0.113 | 0.054 | 0.278 | -0.004 | 0.448 | 0.235 | 0.055 | -0.046 | 0.002 | -0.118 | 0.014 |
| AD | 1.641 | 0.150 | 1.733 | 0.210 | 5.621 | 0.198 | 0.114 | 0.086 | 0.447 | -0.029 | 0.426 | 0.213 | 0.046 | -0.063 | 0.004 | -0.096 | 0.009 |
| 4th Vent - R Inferior colliculus | | | | | | | | | | | | | | | | | |
| FA | 0.187 | 0.022 | 0.180 | 0.019 | -3.801 | -0.164 | 0.115 | 0.160 | 0.964 | -0.393 | 0.066 | -0.164 | 0.027 | 0.011 | 0.000 | 0.069 | 0.005 |
| MD | 1.359 | 0.149 | 1.390 | 0.131 | 2.223 | 0.091 | 0.116 | 0.436 | 0.679 | -0.140 | 0.323 | 0.102 | 0.010 | -0.038 | 0.001 | -0.052 | 0.003 |
| RD | 1.228 | 0.141 | 1.261 | 0.124 | 2.648 | 0.104 | 0.116 | 0.375 | 0.631 | -0.127 | 0.335 | 0.115 | 0.013 | -0.042 | 0.002 | -0.059 | 0.003 |
| AD | 1.622 | 0.171 | 1.648 | 0.148 | 1.579 | 0.068 | 0.117 | 0.564 | 0.786 | -0.164 | 0.300 | 0.076 | 0.006 | -0.029 | 0.001 | -0.039 | 0.002 |
| 4th Vent - R Cochlear nucleus | | | | | | | | | | | | | | | | | |
| FA | 0.183 | 0.019 | 0.174 | 0.019 | -5.046 | -0.239 | 0.119 | **0.048** | 0.849 | -0.476 | -0.002 | -0.225 | 0.051 | -0.057 | 0.003 | 0.135 | 0.018 |
| MD | 1.378 | 0.118 | 1.454 | 0.151 | 5.517 | 0.227 | 0.113 | **0.049** | 0.233 | 0.002 | 0.452 | 0.239 | 0.057 | -0.135 | 0.018 | -0.004 | 0.000 |
| RD | 1.244 | 0.114 | 1.323 | 0.153 | 6.400 | 0.236 | 0.113 | **0.040** | 0.252 | 0.011 | 0.460 | 0.249 | 0.062 | -0.136 | 0.019 | -0.010 | 0.000 |
| AD | 1.647 | 0.138 | 1.716 | 0.155 | 4.182 | 0.197 | 0.114 | 0.089 | 0.447 | -0.030 | 0.423 | 0.207 | 0.043 | -0.125 | 0.016 | 0.007 | 0.000 |
| L Hipp - L Heschl's gyrus | | | | | | | | | | | | | | | | | |
| FA | 0.267 | 0.028 | 0.257 | 0.026 | -3.698 | -0.108 | 0.113 | 0.345 | 0.964 | -0.333 | 0.118 | -0.169 | 0.029 | 0.159 | 0.025 | 0.165 | 0.027 |
| MD | 1.292 | 0.089 | 1.333 | 0.065 | 3.135 | 0.225 | 0.114 | 0.052 | 0.233 | -0.002 | 0.452 | 0.248 | 0.062 | -0.139 | 0.019 | -0.052 | 0.003 |
| RD | 1.111 | 0.076 | 1.146 | 0.068 | 3.127 | 0.190 | 0.115 | 0.103 | 0.312 | -0.039 | 0.418 | 0.221 | 0.049 | -0.139 | 0.019 | -0.080 | 0.006 |
| AD | 1.671 | 0.109 | 1.719 | 0.122 | 2.856 | 0.216 | 0.114 | 0.063 | 0.447 | -0.012 | 0.443 | 0.183 | 0.033 | 0.017 | 0.000 | 0.038 | 0.001 |
| L Hipp - L Medial geniculate nucleus | | | | | | | | | | | | | | | | | |
| FA | 0.224 | 0.033 | 0.215 | 0.032 | -4.039 | -0.104 | 0.115 | 0.369 | 0.964 | -0.334 | 0.125 | -0.127 | 0.016 | 0.036 | 0.001 | 0.123 | 0.015 |
| MD | 1.345 | 0.083 | 1.379 | 0.113 | 2.511 | 0.191 | 0.112 | 0.093 | 0.274 | -0.033 | 0.415 | 0.146 | 0.021 | 0.186 | 0.035 | -0.107 | 0.012 |
| RD | 1.184 | 0.082 | 1.221 | 0.116 | 3.146 | 0.193 | 0.113 | 0.091 | 0.292 | -0.031 | 0.417 | 0.158 | 0.025 | 0.158 | 0.025 | -0.125 | 0.016 |
| AD | 1.668 | 0.109 | 1.694 | 0.124 | 1.608 | 0.159 | 0.113 | 0.162 | 0.518 | -0.065 | 0.384 | 0.103 | 0.011 | 0.209 | 0.044 | -0.060 | 0.004 |
| L VDC - L Inferior colliculus | | | | | | | | | | | | | | | | | |
| FA | 0.210 | 0.038 | 0.205 | 0.039 | -2.511 | 0.003 | 0.113 | 0.982 | 0.986 | -0.223 | 0.228 | -0.063 | 0.004 | 0.073 | 0.005 | 0.240 | 0.057 |
| MD | 1.206 | 0.068 | 1.239 | 0.082 | 2.707 | 0.189 | 0.114 | 0.101 | 0.275 | -0.038 | 0.416 | 0.188 | 0.035 | 0.035 | 0.001 | -0.138 | 0.019 |
| RD | 1.072 | 0.062 | 1.105 | 0.082 | 3.087 | 0.170 | 0.113 | 0.134 | 0.385 | -0.054 | 0.394 | 0.196 | 0.038 | 0.004 | 0.000 | -0.222 | 0.049 |
| AD | 1.475 | 0.110 | 1.506 | 0.113 | 2.155 | 0.166 | 0.115 | 0.155 | 0.518 | -0.064 | 0.395 | 0.129 | 0.017 | 0.069 | 0.005 | 0.015 | 0.000 |
| L VDC - L Cochlear nucleus | | | | | | | | | | | | | | | | | |
| FA | 0.213 | 0.024 | 0.207 | 0.028 | -2.461 | -0.085 | 0.116 | 0.469 | 0.964 | -0.316 | 0.147 | -0.089 | 0.008 | -0.013 | 0.000 | 0.082 | 0.007 |
| MD | 1.221 | 0.069 | 1.252 | 0.074 | 2.552 | 0.173 | 0.115 | 0.135 | 0.340 | -0.055 | 0.401 | 0.193 | 0.037 | -0.097 | 0.009 | -0.067 | 0.004 |
| RD | 1.085 | 0.067 | 1.117 | 0.078 | 2.876 | 0.170 | 0.115 | 0.143 | 0.394 | -0.059 | 0.398 | 0.189 | 0.036 | -0.079 | 0.006 | -0.082 | 0.007 |
| AD | 1.492 | 0.087 | 1.523 | 0.082 | 2.079 | 0.150 | 0.115 | 0.197 | 0.558 | -0.079 | 0.379 | 0.169 | 0.028 | -0.113 | 0.013 | -0.027 | 0.001 |
| L VDC - L Medial geniculate nucleus | | | | | | | | | | | | | | | | | |
| FA | 0.249 | 0.027 | 0.244 | 0.026 | -1.928 | -0.004 | 0.113 | 0.971 | 0.986 | -0.229 | 0.221 | -0.083 | 0.007 | 0.228 | 0.052 | 0.115 | 0.013 |
| MD | 1.227 | 0.076 | 1.264 | 0.067 | 2.954 | 0.243 | 0.114 | **0.035** | 0.209 | 0.017 | 0.470 | 0.234 | 0.055 | -0.036 | 0.001 | -0.029 | 0.001 |
| RD | 1.061 | 0.072 | 1.096 | 0.064 | 3.311 | 0.221 | 0.114 | 0.056 | 0.278 | -0.005 | 0.447 | 0.237 | 0.056 | -0.099 | 0.010 | -0.066 | 0.004 |
| AD | 1.560 | 0.102 | 1.599 | 0.089 | 2.470 | 0.231 | 0.114 | **0.046** | 0.378 | 0.005 | 0.458 | 0.187 | 0.035 | 0.061 | 0.004 | 0.030 | 0.001 |
| L VDC - R Inferior colliculus | | | | | | | | | | | | | | | | | |
| FA | 0.223 | 0.045 | 0.224 | 0.041 | 0.627 | 0.035 | 0.117 | 0.767 | 0.974 | -0.198 | 0.267 | 0.015 | 0.000 | 0.032 | 0.001 | 0.042 | 0.002 |
| MD | 1.226 | 0.095 | 1.241 | 0.105 | 1.202 | 0.021 | 0.116 | 0.858 | 0.943 | -0.209 | 0.251 | 0.066 | 0.004 | -0.117 | 0.014 | -0.093 | 0.009 |
| RD | 1.078 | 0.088 | 1.091 | 0.097 | 1.158 | 0.012 | 0.116 | 0.916 | 0.989 | -0.218 | 0.242 | 0.061 | 0.004 | -0.122 | 0.015 | -0.098 | 0.010 |
| AD | 1.522 | 0.141 | 1.542 | 0.148 | 1.265 | 0.028 | 0.116 | 0.810 | 0.943 | -0.203 | 0.259 | 0.061 | 0.004 | -0.088 | 0.008 | -0.068 | 0.005 |
| R CC - L Inferior colliculus | | | | | | | | | | | | | | | | | |
| FA | 0.264 | 0.025 | 0.256 | 0.017 | -3.004 | -0.125 | 0.113 | 0.272 | 0.964 | -0.349 | 0.100 | -0.183 | 0.033 | 0.225 | 0.051 | 0.070 | 0.005 |
| MD | 1.327 | 0.102 | 1.320 | 0.071 | -0.459 | -0.087 | 0.114 | 0.448 | 0.679 | -0.313 | 0.140 | -0.034 | 0.001 | -0.002 | 0.000 | -0.221 | 0.049 |
| RD | 1.136 | 0.099 | 1.137 | 0.065 | 0.110 | -0.055 | 0.114 | 0.629 | 0.806 | -0.281 | 0.171 | 0.008 | 0.000 | -0.051 | 0.003 | -0.228 | 0.052 |
| AD | 1.708 | 0.117 | 1.687 | 0.093 | -1.217 | -0.127 | 0.114 | 0.268 | 0.558 | -0.353 | 0.100 | -0.095 | 0.009 | 0.072 | 0.005 | -0.192 | 0.037 |
| R CC - L Medial geniculate nucleus | | | | | | | | | | | | | | | | | |
| FA | 0.257 | 0.015 | 0.247 | 0.012 | -3.808 | -0.285 | 0.110 | **0.011** | 0.682 | -0.503 | -0.066 | -0.323 | 0.104 | 0.183 | 0.033 | 0.105 | 0.011 |
| MD | 1.317 | 0.053 | 1.350 | 0.054 | 2.497 | 0.265 | 0.112 | **0.020** | 0.206 | 0.042 | 0.488 | 0.269 | 0.072 | 0.011 | 0.000 | -0.176 | 0.031 |
| RD | 1.139 | 0.059 | 1.171 | 0.053 | 2.808 | 0.256 | 0.111 | **0.024** | 0.173 | 0.034 | 0.477 | 0.258 | 0.067 | 0.031 | 0.001 | -0.184 | 0.034 |
| AD | 1.697 | 0.087 | 1.715 | 0.071 | 1.033 | 0.126 | 0.114 | 0.273 | 0.558 | -0.101 | 0.353 | 0.105 | 0.011 | 0.133 | 0.018 | -0.129 | 0.017 |
| R CC - R Inferior colliculus | | | | | | | | | | | | | | | | | |
| FA | 0.230 | 0.018 | 0.224 | 0.014 | -2.500 | -0.133 | 0.114 | 0.245 | 0.964 | -0.360 | 0.093 | -0.173 | 0.030 | 0.066 | 0.004 | 0.185 | 0.034 |
| MD | 1.331 | 0.073 | 1.337 | 0.062 | 0.514 | 0.063 | 0.115 | 0.586 | 0.796 | -0.166 | 0.293 | 0.048 | 0.002 | 0.118 | 0.014 | -0.109 | 0.012 |
| RD | 1.168 | 0.073 | 1.179 | 0.060 | 0.934 | 0.084 | 0.115 | 0.469 | 0.681 | -0.145 | 0.312 | 0.079 | 0.006 | 0.096 | 0.009 | -0.141 | 0.020 |
| AD | 1.656 | 0.079 | 1.655 | 0.071 | -0.078 | 0.022 | 0.116 | 0.848 | 0.957 | -0.208 | 0.252 | -0.008 | 0.000 | 0.146 | 0.021 | -0.045 | 0.002 |
| R CC - R Heschl's gyrus | | | | | | | | | | | | | | | | | |
| FA | 0.202 | 0.013 | 0.198 | 0.012 | -2.101 | -0.121 | 0.113 | 0.288 | 0.964 | -0.345 | 0.104 | -0.160 | 0.026 | 0.017 | 0.000 | 0.240 | 0.058 |
| MD | 1.303 | 0.052 | 1.318 | 0.042 | 1.127 | 0.117 | 0.114 | 0.308 | 0.535 | -0.110 | 0.344 | 0.149 | 0.022 | -0.021 | 0.000 | -0.195 | 0.038 |
| RD | 1.164 | 0.055 | 1.181 | 0.043 | 1.408 | 0.123 | 0.113 | 0.283 | 0.542 | -0.103 | 0.348 | 0.159 | 0.025 | -0.025 | 0.001 | -0.218 | 0.048 |
| AD | 1.581 | 0.049 | 1.592 | 0.045 | 0.713 | 0.094 | 0.116 | 0.417 | 0.681 | -0.136 | 0.324 | 0.112 | 0.013 | -0.011 | 0.000 | -0.128 | 0.016 |
| R CC - R Medial geniculate nucleus | | | | | | | | | | | | | | | | | |
| FA | 0.221 | 0.016 | 0.216 | 0.011 | -2.463 | -0.107 | 0.109 | 0.330 | 0.964 | -0.324 | 0.110 | -0.197 | 0.039 | 0.161 | 0.026 | 0.308 | 0.095 |
| MD | 1.325 | 0.055 | 1.341 | 0.053 | 1.252 | 0.144 | 0.114 | 0.212 | 0.432 | -0.084 | 0.371 | 0.142 | 0.020 | 0.069 | 0.005 | -0.153 | 0.024 |
| RD | 1.168 | 0.056 | 1.187 | 0.050 | 1.612 | 0.148 | 0.113 | 0.196 | 0.430 | -0.078 | 0.373 | 0.165 | 0.027 | 0.032 | 0.001 | -0.204 | 0.042 |
| AD | 1.638 | 0.061 | 1.650 | 0.061 | 0.740 | 0.125 | 0.115 | 0.282 | 0.558 | -0.104 | 0.354 | 0.091 | 0.008 | 0.127 | 0.016 | -0.054 | 0.003 |
| R Lat Vent - R Heschl's gyrus | | | | | | | | | | | | | | | | | |
| FA | 0.246 | 0.021 | 0.246 | 0.026 | -0.055 | 0.002 | 0.116 | 0.986 | 0.986 | -0.229 | 0.233 | -0.003 | 0.000 | -0.071 | 0.005 | 0.114 | 0.013 |
| MD | 1.357 | 0.074 | 1.372 | 0.097 | 1.085 | 0.114 | 0.115 | 0.327 | 0.543 | -0.116 | 0.343 | 0.075 | 0.006 | 0.137 | 0.019 | -0.034 | 0.001 |
| RD | 1.177 | 0.076 | 1.191 | 0.095 | 1.146 | 0.103 | 0.115 | 0.374 | 0.631 | -0.126 | 0.332 | 0.069 | 0.005 | 0.138 | 0.019 | -0.057 | 0.003 |
| AD | 1.718 | 0.079 | 1.735 | 0.113 | 1.001 | 0.119 | 0.115 | 0.304 | 0.566 | -0.110 | 0.349 | 0.075 | 0.006 | 0.121 | 0.015 | 0.009 | 0.000 |
| R Lat Vent - R Medial geniculate nucleus | | | | | | | | | | | | | | | | | |
| FA | 0.249 | 0.014 | 0.246 | 0.019 | -1.058 | 0.065 | 0.107 | 0.548 | 0.964 | -0.148 | 0.277 | -0.067 | 0.004 | 0.281 | 0.079 | 0.279 | 0.078 |
| MD | 1.339 | 0.083 | 1.385 | 0.076 | 3.390 | 0.248 | 0.113 | **0.031** | 0.206 | 0.023 | 0.472 | 0.259 | 0.067 | -0.117 | 0.014 | -0.030 | 0.001 |
| RD | 1.159 | 0.076 | 1.202 | 0.074 | 3.646 | 0.215 | 0.112 | 0.059 | 0.278 | -0.008 | 0.439 | 0.252 | 0.064 | -0.165 | 0.027 | -0.086 | 0.007 |
| AD | 1.699 | 0.102 | 1.751 | 0.089 | 3.041 | 0.281 | 0.113 | **0.015** | 0.294 | 0.057 | 0.504 | 0.249 | 0.062 | -0.031 | 0.001 | 0.062 | 0.004 |
| R Cb - L Inferior colliculus | | | | | | | | | | | | | | | | | |
| FA | 0.189 | 0.023 | 0.187 | 0.022 | -1.002 | -0.011 | 0.116 | 0.928 | 0.986 | -0.242 | 0.221 | -0.040 | 0.002 | 0.094 | 0.009 | 0.034 | 0.001 |
| MD | 1.176 | 0.070 | 1.195 | 0.057 | 1.595 | 0.140 | 0.116 | 0.229 | 0.441 | -0.090 | 0.371 | 0.140 | 0.020 | -0.041 | 0.002 | -0.015 | 0.000 |
| RD | 1.057 | 0.059 | 1.076 | 0.053 | 1.775 | 0.148 | 0.116 | 0.203 | 0.430 | -0.081 | 0.378 | 0.156 | 0.024 | -0.071 | 0.005 | -0.024 | 0.001 |
| AD | 1.415 | 0.101 | 1.434 | 0.079 | 1.327 | 0.111 | 0.116 | 0.341 | 0.584 | -0.120 | 0.343 | 0.100 | 0.010 | 0.003 | 0.000 | -0.001 | 0.000 |
| R Cb - L Cochlear nucleus | | | | | | | | | | | | | | | | | |
| FA | 0.195 | 0.028 | 0.184 | 0.032 | -5.744 | -0.133 | 0.115 | 0.251 | 0.964 | -0.361 | 0.096 | -0.165 | 0.027 | 0.088 | 0.008 | 0.122 | 0.015 |
| MD | 1.264 | 0.134 | 1.321 | 0.192 | 4.487 | 0.138 | 0.116 | 0.236 | 0.446 | -0.092 | 0.368 | 0.146 | 0.021 | -0.074 | 0.006 | -0.015 | 0.000 |
| RD | 1.134 | 0.125 | 1.195 | 0.191 | 5.399 | 0.149 | 0.115 | 0.199 | 0.430 | -0.080 | 0.379 | 0.160 | 0.026 | -0.079 | 0.006 | -0.028 | 0.001 |
| AD | 1.524 | 0.160 | 1.572 | 0.200 | 3.130 | 0.112 | 0.116 | 0.337 | 0.584 | -0.119 | 0.343 | 0.115 | 0.013 | -0.064 | 0.004 | 0.010 | 0.000 |
| R Cb - L Medial geniculate nucleus | | | | | | | | | | | | | | | | | |
| FA | 0.214 | 0.021 | 0.217 | 0.025 | 1.424 | 0.092 | 0.116 | 0.430 | 0.964 | -0.139 | 0.322 | 0.058 | 0.003 | 0.115 | 0.013 | -0.024 | 0.001 |
| MD | 1.173 | 0.068 | 1.198 | 0.053 | 2.120 | 0.189 | 0.114 | 0.100 | 0.275 | -0.037 | 0.416 | 0.196 | 0.038 | 0.015 | 0.000 | -0.147 | 0.022 |
| RD | 1.036 | 0.061 | 1.056 | 0.041 | 1.886 | 0.166 | 0.114 | 0.150 | 0.394 | -0.061 | 0.393 | 0.185 | 0.034 | -0.028 | 0.001 | -0.147 | 0.022 |
| AD | 1.447 | 0.092 | 1.482 | 0.090 | 2.455 | 0.187 | 0.114 | 0.105 | 0.477 | -0.040 | 0.414 | 0.178 | 0.032 | 0.058 | 0.003 | -0.126 | 0.016 |
| R Cb - R Inferior colliculus | | | | | | | | | | | | | | | | | |
| FA | 0.174 | 0.016 | 0.173 | 0.013 | -0.738 | -0.060 | 0.115 | 0.604 | 0.974 | -0.288 | 0.169 | -0.042 | 0.002 | -0.139 | 0.019 | 0.119 | 0.014 |
| MD | 1.208 | 0.065 | 1.217 | 0.060 | 0.663 | 0.051 | 0.116 | 0.659 | 0.822 | -0.180 | 0.282 | 0.060 | 0.004 | 0.033 | 0.001 | -0.113 | 0.013 |
| RD | 1.099 | 0.065 | 1.107 | 0.057 | 0.804 | 0.063 | 0.116 | 0.588 | 0.775 | -0.167 | 0.293 | 0.068 | 0.005 | 0.053 | 0.003 | -0.127 | 0.016 |
| AD | 1.428 | 0.071 | 1.435 | 0.069 | 0.446 | 0.028 | 0.117 | 0.810 | 0.943 | -0.204 | 0.260 | 0.042 | 0.002 | -0.003 | 0.000 | -0.082 | 0.007 |
| R Cb - R Cochlear nucleus | | | | | | | | | | | | | | | | | |
| FA | 0.170 | 0.016 | 0.167 | 0.019 | -2.127 | -0.067 | 0.115 | 0.563 | 0.964 | -0.295 | 0.162 | -0.091 | 0.008 | -0.015 | 0.000 | 0.176 | 0.031 |
| MD | 1.216 | 0.090 | 1.243 | 0.076 | 2.231 | 0.151 | 0.116 | 0.196 | 0.425 | -0.079 | 0.380 | 0.153 | 0.024 | -0.067 | 0.004 | -0.005 | 0.000 |
| RD | 1.108 | 0.086 | 1.137 | 0.076 | 2.599 | 0.158 | 0.115 | 0.175 | 0.413 | -0.072 | 0.388 | 0.165 | 0.027 | -0.062 | 0.004 | -0.035 | 0.001 |
| AD | 1.433 | 0.101 | 1.456 | 0.083 | 1.661 | 0.128 | 0.116 | 0.273 | 0.558 | -0.102 | 0.358 | 0.123 | 0.015 | -0.072 | 0.005 | 0.051 | 0.003 |
| R Cb - R Medial geniculate nucleus | | | | | | | | | | | | | | | | | |
| FA | 0.198 | 0.020 | 0.194 | 0.017 | -1.981 | -0.082 | 0.116 | 0.484 | 0.964 | -0.312 | 0.149 | -0.102 | 0.010 | 0.032 | 0.001 | 0.103 | 0.011 |
| MD | 1.181 | 0.058 | 1.188 | 0.045 | 0.614 | 0.049 | 0.117 | 0.675 | 0.822 | -0.183 | 0.281 | 0.068 | 0.005 | -0.054 | 0.003 | -0.050 | 0.003 |
| RD | 1.056 | 0.057 | 1.065 | 0.042 | 0.917 | 0.072 | 0.116 | 0.538 | 0.731 | -0.159 | 0.303 | 0.095 | 0.009 | -0.062 | 0.004 | -0.077 | 0.006 |
| AD | 1.431 | 0.072 | 1.433 | 0.062 | 0.167 | 0.008 | 0.117 | 0.943 | 0.960 | -0.224 | 0.241 | 0.017 | 0.000 | -0.034 | 0.001 | -0.004 | 0.000 |
| R Thal - L Inferior colliculus | | | | | | | | | | | | | | | | | |
| FA | 0.239 | 0.031 | 0.232 | 0.029 | -2.963 | -0.073 | 0.113 | 0.524 | 0.964 | -0.298 | 0.153 | -0.110 | 0.012 | 0.233 | 0.054 | -0.074 | 0.005 |
| MD | 1.260 | 0.104 | 1.267 | 0.095 | 0.535 | -0.010 | 0.116 | 0.931 | 0.987 | -0.240 | 0.220 | 0.032 | 0.001 | -0.053 | 0.003 | -0.141 | 0.020 |
| RD | 1.098 | 0.096 | 1.109 | 0.090 | 1.018 | 0.006 | 0.115 | 0.958 | 0.989 | -0.224 | 0.236 | 0.056 | 0.003 | -0.112 | 0.013 | -0.115 | 0.013 |
| AD | 1.585 | 0.139 | 1.582 | 0.121 | -0.136 | -0.032 | 0.115 | 0.781 | 0.943 | -0.262 | 0.197 | -0.008 | 0.000 | 0.040 | 0.002 | -0.161 | 0.026 |
| R Thal - L Medial geniculate nucleus | | | | | | | | | | | | | | | | | |
| FA | 0.267 | 0.021 | 0.258 | 0.022 | -3.484 | -0.115 | 0.111 | 0.305 | 0.964 | -0.336 | 0.106 | -0.193 | 0.037 | 0.259 | 0.067 | 0.125 | 0.016 |
| MD | 1.272 | 0.066 | 1.313 | 0.056 | 3.168 | 0.307 | 0.110 | **0.007** | 0.206 | 0.087 | 0.526 | 0.299 | 0.090 | 0.037 | 0.001 | -0.165 | 0.027 |
| RD | 1.085 | 0.057 | 1.128 | 0.057 | 3.985 | 0.314 | 0.109 | **0.005** | 0.167 | 0.096 | 0.531 | 0.331 | 0.109 | -0.044 | 0.002 | -0.190 | 0.036 |
| AD | 1.657 | 0.086 | 1.682 | 0.071 | 1.530 | 0.206 | 0.112 | 0.069 | 0.447 | -0.017 | 0.430 | 0.152 | 0.023 | 0.198 | 0.039 | -0.065 | 0.004 |
| R Thal - R Inferior colliculus | | | | | | | | | | | | | | | | | |
| FA | 0.226 | 0.017 | 0.220 | 0.015 | -2.542 | -0.164 | 0.115 | 0.159 | 0.964 | -0.394 | 0.065 | -0.165 | 0.027 | 0.062 | 0.004 | 0.007 | 0.000 |
| MD | 1.214 | 0.057 | 1.217 | 0.052 | 0.254 | -0.021 | 0.114 | 0.855 | 0.943 | -0.249 | 0.207 | 0.027 | 0.001 | -0.021 | 0.000 | -0.207 | 0.043 |
| RD | 1.069 | 0.055 | 1.076 | 0.050 | 0.705 | 0.023 | 0.115 | 0.839 | 0.956 | -0.205 | 0.251 | 0.068 | 0.005 | -0.031 | 0.001 | -0.199 | 0.040 |
| AD | 1.506 | 0.069 | 1.500 | 0.065 | -0.385 | -0.087 | 0.114 | 0.447 | 0.705 | -0.315 | 0.140 | -0.040 | 0.002 | -0.003 | 0.000 | -0.193 | 0.037 |
| R Thal - R Heschl's gyrus | | | | | | | | | | | | | | | | | |
| FA | 0.250 | 0.015 | 0.246 | 0.018 | -1.387 | -0.070 | 0.115 | 0.544 | 0.964 | -0.300 | 0.159 | -0.093 | 0.009 | 0.002 | 0.000 | 0.147 | 0.022 |
| MD | 1.230 | 0.049 | 1.254 | 0.066 | 1.951 | 0.213 | 0.114 | 0.067 | 0.236 | -0.015 | 0.440 | 0.178 | 0.032 | 0.051 | 0.003 | 0.007 | 0.000 |
| RD | 1.063 | 0.049 | 1.087 | 0.060 | 2.247 | 0.217 | 0.114 | 0.062 | 0.278 | -0.011 | 0.444 | 0.189 | 0.036 | 0.044 | 0.002 | -0.027 | 0.001 |
| AD | 1.565 | 0.057 | 1.589 | 0.086 | 1.550 | 0.184 | 0.115 | 0.112 | 0.477 | -0.044 | 0.413 | 0.141 | 0.020 | 0.057 | 0.003 | 0.054 | 0.003 |
| R Thal - R Cochlear nucleus | | | | | | | | | | | | | | | | | |
| FA | 0.230 | 0.015 | 0.228 | 0.016 | -0.779 | -0.057 | 0.117 | 0.629 | 0.974 | -0.289 | 0.176 | -0.052 | 0.003 | 0.041 | 0.002 | -0.047 | 0.002 |
| MD | 1.121 | 0.050 | 1.143 | 0.050 | 1.909 | 0.179 | 0.115 | 0.122 | 0.316 | -0.049 | 0.407 | 0.195 | 0.038 | -0.092 | 0.008 | -0.054 | 0.003 |
| RD | 0.984 | 0.039 | 1.004 | 0.046 | 2.047 | 0.191 | 0.114 | 0.099 | 0.309 | -0.037 | 0.418 | 0.207 | 0.043 | -0.105 | 0.011 | -0.043 | 0.002 |
| AD | 1.397 | 0.076 | 1.421 | 0.065 | 1.716 | 0.145 | 0.115 | 0.212 | 0.558 | -0.084 | 0.375 | 0.159 | 0.025 | -0.065 | 0.004 | -0.062 | 0.004 |
| R Thal - R Medial geniculate nucleus | | | | | | | | | | | | | | | | | |
| FA | 0.235 | 0.014 | 0.233 | 0.018 | -0.727 | 0.023 | 0.114 | 0.842 | 0.974 | -0.205 | 0.250 | -0.047 | 0.002 | 0.161 | 0.026 | 0.137 | 0.019 |
| MD | 1.223 | 0.044 | 1.237 | 0.050 | 1.135 | 0.151 | 0.116 | 0.195 | 0.425 | -0.079 | 0.382 | 0.132 | 0.017 | 0.027 | 0.001 | -0.013 | 0.000 |
| RD | 1.068 | 0.040 | 1.081 | 0.045 | 1.265 | 0.135 | 0.116 | 0.246 | 0.489 | -0.095 | 0.366 | 0.140 | 0.020 | -0.023 | 0.001 | -0.062 | 0.004 |
| AD | 1.533 | 0.059 | 1.548 | 0.070 | 0.954 | 0.148 | 0.115 | 0.202 | 0.558 | -0.081 | 0.378 | 0.101 | 0.010 | 0.090 | 0.008 | 0.053 | 0.003 |
| R Caud - R Inferior colliculus | | | | | | | | | | | | | | | | | |
| FA | 0.226 | 0.023 | 0.228 | 0.021 | 0.820 | 0.006 | 0.116 | 0.960 | 0.986 | -0.225 | 0.237 | 0.039 | 0.002 | -0.118 | 0.014 | -0.020 | 0.000 |
| MD | 1.156 | 0.067 | 1.174 | 0.057 | 1.550 | 0.165 | 0.115 | 0.158 | 0.371 | -0.065 | 0.394 | 0.136 | 0.019 | 0.046 | 0.002 | 0.003 | 0.000 |
| RD | 1.017 | 0.064 | 1.031 | 0.056 | 1.394 | 0.145 | 0.116 | 0.212 | 0.441 | -0.085 | 0.375 | 0.111 | 0.012 | 0.076 | 0.006 | 0.004 | 0.000 |
| AD | 1.435 | 0.084 | 1.461 | 0.069 | 1.771 | 0.172 | 0.115 | 0.140 | 0.513 | -0.058 | 0.402 | 0.156 | 0.024 | -0.007 | 0.000 | 0.002 | 0.000 |
| R Caud - R Heschl's gyrus | | | | | | | | | | | | | | | | | |
| FA | 0.239 | 0.027 | 0.237 | 0.014 | -0.889 | -0.041 | 0.114 | 0.720 | 0.974 | -0.267 | 0.186 | -0.052 | 0.003 | -0.101 | 0.010 | 0.208 | 0.043 |
| MD | 1.223 | 0.055 | 1.216 | 0.060 | -0.506 | -0.001 | 0.114 | 0.995 | 0.995 | -0.228 | 0.226 | -0.049 | 0.002 | 0.218 | 0.047 | -0.036 | 0.001 |
| RD | 1.064 | 0.065 | 1.060 | 0.057 | -0.299 | 0.014 | 0.114 | 0.901 | 0.988 | -0.212 | 0.241 | -0.025 | 0.001 | 0.214 | 0.046 | -0.085 | 0.007 |
| AD | 1.541 | 0.051 | 1.528 | 0.071 | -0.791 | -0.028 | 0.115 | 0.810 | 0.943 | -0.256 | 0.200 | -0.085 | 0.007 | 0.193 | 0.037 | 0.058 | 0.003 |
| R Caud - R Medial geniculate nucleus | | | | | | | | | | | | | | | | | |
| FA | 0.234 | 0.016 | 0.227 | 0.017 | -3.183 | -0.132 | 0.111 | 0.239 | 0.964 | -0.354 | 0.089 | -0.201 | 0.040 | 0.140 | 0.019 | 0.239 | 0.057 |
| MD | 1.194 | 0.046 | 1.220 | 0.048 | 2.235 | 0.257 | 0.115 | **0.028** | 0.206 | 0.029 | 0.485 | 0.249 | 0.062 | -0.046 | 0.002 | -0.037 | 0.001 |
| RD | 1.047 | 0.048 | 1.071 | 0.044 | 2.268 | 0.211 | 0.114 | 0.067 | 0.278 | -0.015 | 0.437 | 0.234 | 0.055 | -0.111 | 0.012 | -0.081 | 0.007 |
| AD | 1.504 | 0.073 | 1.519 | 0.066 | 0.958 | 0.122 | 0.116 | 0.297 | 0.562 | -0.109 | 0.352 | 0.097 | 0.009 | -0.018 | 0.000 | 0.089 | 0.008 |
| R Put - R Inferior colliculus | | | | | | | | | | | | | | | | | |
| FA | 0.261 | 0.019 | 0.257 | 0.016 | -1.401 | -0.116 | 0.116 | 0.321 | 0.964 | -0.347 | 0.115 | -0.100 | 0.010 | -0.031 | 0.001 | 0.018 | 0.000 |
| MD | 1.153 | 0.061 | 1.159 | 0.061 | 0.544 | 0.072 | 0.116 | 0.538 | 0.770 | -0.159 | 0.303 | 0.048 | 0.002 | 0.100 | 0.010 | -0.041 | 0.002 |
| RD | 0.989 | 0.062 | 0.997 | 0.059 | 0.772 | 0.083 | 0.116 | 0.476 | 0.681 | -0.148 | 0.314 | 0.059 | 0.003 | 0.099 | 0.010 | -0.046 | 0.002 |
| AD | 1.481 | 0.068 | 1.484 | 0.072 | 0.241 | 0.045 | 0.116 | 0.698 | 0.911 | -0.186 | 0.277 | 0.023 | 0.001 | 0.093 | 0.009 | -0.029 | 0.001 |
| R Put - R Heschl's gyrus | | | | | | | | | | | | | | | | | |
| FA | 0.245 | 0.017 | 0.240 | 0.016 | -2.143 | -0.121 | 0.114 | 0.289 | 0.964 | -0.347 | 0.105 | -0.149 | 0.022 | -0.004 | 0.000 | 0.212 | 0.045 |
| MD | 1.185 | 0.043 | 1.186 | 0.039 | 0.083 | 0.026 | 0.116 | 0.824 | 0.929 | -0.205 | 0.257 | 0.011 | 0.000 | 0.095 | 0.009 | -0.061 | 0.004 |
| RD | 1.028 | 0.044 | 1.033 | 0.037 | 0.485 | 0.063 | 0.116 | 0.590 | 0.775 | -0.167 | 0.293 | 0.059 | 0.003 | 0.085 | 0.007 | -0.121 | 0.015 |
| AD | 1.499 | 0.050 | 1.492 | 0.053 | -0.469 | -0.034 | 0.116 | 0.769 | 0.943 | -0.266 | 0.197 | -0.062 | 0.004 | 0.091 | 0.008 | 0.040 | 0.002 |
| R Put - R Medial geniculate nucleus | | | | | | | | | | | | | | | | | |
| FA | 0.268 | 0.018 | 0.262 | 0.017 | -2.496 | -0.070 | 0.108 | 0.518 | 0.964 | -0.286 | 0.145 | -0.178 | 0.032 | 0.322 | 0.103 | 0.175 | 0.031 |
| MD | 1.198 | 0.050 | 1.192 | 0.054 | -0.469 | -0.017 | 0.116 | 0.881 | 0.943 | -0.249 | 0.214 | -0.049 | 0.002 | 0.061 | 0.004 | 0.092 | 0.008 |
| RD | 1.021 | 0.047 | 1.021 | 0.047 | -0.030 | -0.005 | 0.117 | 0.964 | 0.989 | -0.238 | 0.227 | -0.003 | 0.000 | -0.036 | 0.001 | 0.038 | 0.001 |
| AD | 1.550 | 0.068 | 1.534 | 0.076 | -1.048 | -0.031 | 0.114 | 0.788 | 0.943 | -0.257 | 0.196 | -0.101 | 0.010 | 0.176 | 0.031 | 0.149 | 0.022 |
| R Pal - R Medial geniculate nucleus | | | | | | | | | | | | | | | | | |
| FA | 0.271 | 0.031 | 0.261 | 0.025 | -3.442 | -0.136 | 0.115 | 0.243 | 0.964 | -0.365 | 0.094 | -0.157 | 0.025 | 0.063 | 0.004 | 0.098 | 0.010 |
| MD | 1.179 | 0.080 | 1.169 | 0.070 | -0.833 | -0.030 | 0.116 | 0.794 | 0.925 | -0.261 | 0.200 | -0.062 | 0.004 | 0.029 | 0.001 | 0.140 | 0.020 |
| RD | 1.003 | 0.062 | 1.001 | 0.059 | -0.169 | 0.009 | 0.116 | 0.938 | 0.989 | -0.222 | 0.241 | -0.013 | 0.000 | 0.003 | 0.000 | 0.106 | 0.011 |
| AD | 1.531 | 0.130 | 1.505 | 0.106 | -1.703 | -0.068 | 0.115 | 0.555 | 0.784 | -0.297 | 0.161 | -0.106 | 0.011 | 0.053 | 0.003 | 0.158 | 0.025 |
| R Hipp - R Medial geniculate nucleus | | | | | | | | | | | | | | | | | |
| FA | 0.221 | 0.037 | 0.219 | 0.034 | -1.092 | 0.026 | 0.114 | 0.820 | 0.974 | -0.201 | 0.253 | -0.032 | 0.001 | 0.060 | 0.004 | 0.208 | 0.043 |
| MD | 1.374 | 0.092 | 1.357 | 0.076 | -1.198 | -0.093 | 0.114 | 0.415 | 0.657 | -0.320 | 0.133 | -0.093 | 0.009 | 0.151 | 0.023 | -0.148 | 0.022 |
| RD | 1.213 | 0.100 | 1.199 | 0.078 | -1.106 | -0.092 | 0.114 | 0.419 | 0.681 | -0.319 | 0.134 | -0.072 | 0.005 | 0.108 | 0.012 | -0.195 | 0.038 |
| AD | 1.696 | 0.102 | 1.673 | 0.098 | -1.331 | -0.070 | 0.115 | 0.546 | 0.782 | -0.298 | 0.159 | -0.104 | 0.011 | 0.186 | 0.034 | -0.026 | 0.001 |
| R Amygdala - R Medial geniculate nucleus | | | | | | | | | | | | | | | | | |
| FA | 0.227 | 0.030 | 0.221 | 0.027 | -2.777 | -0.031 | 0.111 | 0.779 | 0.974 | -0.252 | 0.190 | -0.105 | 0.011 | 0.070 | 0.005 | 0.302 | 0.091 |
| MD | 1.287 | 0.082 | 1.256 | 0.067 | -2.357 | -0.168 | 0.113 | 0.142 | 0.343 | -0.393 | 0.058 | -0.192 | 0.037 | 0.190 | 0.036 | -0.036 | 0.001 |
| RD | 1.129 | 0.084 | 1.106 | 0.059 | -2.024 | -0.154 | 0.113 | 0.177 | 0.413 | -0.380 | 0.071 | -0.155 | 0.024 | 0.160 | 0.026 | -0.128 | 0.016 |
| AD | 1.602 | 0.098 | 1.557 | 0.097 | -2.827 | -0.158 | 0.113 | 0.166 | 0.518 | -0.382 | 0.067 | -0.210 | 0.044 | 0.197 | 0.039 | 0.096 | 0.009 |
| R VDC - L Inferior colliculus | | | | | | | | | | | | | | | | | |
| FA | 0.220 | 0.049 | 0.212 | 0.039 | -3.536 | -0.043 | 0.115 | 0.712 | 0.974 | -0.272 | 0.187 | -0.085 | 0.007 | 0.082 | 0.007 | 0.132 | 0.017 |
| MD | 1.241 | 0.081 | 1.244 | 0.102 | 0.235 | -0.057 | 0.114 | 0.618 | 0.818 | -0.283 | 0.169 | 0.014 | 0.000 | -0.098 | 0.010 | -0.209 | 0.044 |
| RD | 1.094 | 0.081 | 1.103 | 0.096 | 0.825 | -0.037 | 0.112 | 0.745 | 0.907 | -0.260 | 0.187 | 0.045 | 0.002 | -0.122 | 0.015 | -0.244 | 0.059 |
| AD | 1.534 | 0.126 | 1.524 | 0.138 | -0.607 | -0.072 | 0.116 | 0.538 | 0.782 | -0.302 | 0.159 | -0.032 | 0.001 | -0.043 | 0.002 | -0.113 | 0.013 |
| R VDC - L Cochlear nucleus | | | | | | | | | | | | | | | | | |
| FA | 0.219 | 0.036 | 0.211 | 0.032 | -3.808 | -0.111 | 0.115 | 0.341 | 0.964 | -0.340 | 0.119 | -0.115 | 0.013 | 0.109 | 0.012 | -0.064 | 0.004 |
| MD | 1.250 | 0.067 | 1.288 | 0.086 | 3.053 | 0.217 | 0.114 | 0.061 | 0.233 | -0.010 | 0.445 | 0.213 | 0.045 | -0.044 | 0.002 | -0.032 | 0.001 |
| RD | 1.106 | 0.071 | 1.146 | 0.091 | 3.637 | 0.211 | 0.114 | 0.068 | 0.278 | -0.016 | 0.439 | 0.211 | 0.044 | -0.073 | 0.005 | -0.011 | 0.000 |
| AD | 1.538 | 0.090 | 1.573 | 0.093 | 2.212 | 0.179 | 0.115 | 0.124 | 0.477 | -0.050 | 0.408 | 0.168 | 0.028 | 0.020 | 0.000 | -0.063 | 0.004 |
| R VDC - R Inferior colliculus | | | | | | | | | | | | | | | | | |
| FA | 0.216 | 0.023 | 0.214 | 0.031 | -1.053 | -0.055 | 0.116 | 0.638 | 0.974 | -0.286 | 0.176 | -0.036 | 0.001 | -0.087 | 0.008 | 0.044 | 0.002 |
| MD | 1.219 | 0.090 | 1.216 | 0.070 | -0.263 | -0.110 | 0.112 | 0.328 | 0.543 | -0.334 | 0.113 | -0.019 | 0.000 | -0.156 | 0.024 | -0.210 | 0.044 |
| RD | 1.078 | 0.086 | 1.077 | 0.067 | -0.081 | -0.086 | 0.113 | 0.451 | 0.681 | -0.310 | 0.139 | -0.006 | 0.000 | -0.121 | 0.015 | -0.213 | 0.045 |
| AD | 1.501 | 0.108 | 1.493 | 0.098 | -0.524 | -0.127 | 0.112 | 0.263 | 0.558 | -0.350 | 0.097 | -0.036 | 0.001 | -0.180 | 0.032 | -0.170 | 0.029 |
| R VDC - R Heschl's gyrus | | | | | | | | | | | | | | | | | |
| FA | 0.271 | 0.015 | 0.266 | 0.017 | -1.932 | -0.088 | 0.112 | 0.434 | 0.964 | -0.311 | 0.135 | -0.150 | 0.022 | 0.076 | 0.006 | 0.263 | 0.069 |
| MD | 1.218 | 0.049 | 1.224 | 0.056 | 0.484 | 0.033 | 0.116 | 0.776 | 0.914 | -0.198 | 0.265 | 0.050 | 0.002 | 0.000 | 0.000 | -0.103 | 0.011 |
| RD | 1.037 | 0.048 | 1.047 | 0.054 | 0.967 | 0.056 | 0.115 | 0.631 | 0.806 | -0.174 | 0.285 | 0.087 | 0.008 | -0.024 | 0.001 | -0.160 | 0.026 |
| AD | 1.582 | 0.058 | 1.579 | 0.069 | -0.149 | -0.007 | 0.117 | 0.951 | 0.960 | -0.240 | 0.225 | -0.017 | 0.000 | 0.038 | 0.001 | 0.002 | 0.000 |
| R VDC - R Cochlear nucleus | | | | | | | | | | | | | | | | | |
| FA | 0.214 | 0.024 | 0.209 | 0.024 | -2.166 | -0.075 | 0.116 | 0.520 | 0.964 | -0.307 | 0.156 | -0.089 | 0.008 | 0.032 | 0.001 | 0.066 | 0.004 |
| MD | 1.213 | 0.062 | 1.242 | 0.062 | 2.358 | 0.200 | 0.114 | 0.084 | 0.270 | -0.027 | 0.427 | 0.208 | 0.043 | -0.109 | 0.012 | -0.001 | 0.000 |
| RD | 1.076 | 0.058 | 1.106 | 0.063 | 2.746 | 0.204 | 0.114 | 0.077 | 0.283 | -0.023 | 0.431 | 0.216 | 0.047 | -0.113 | 0.013 | -0.021 | 0.000 |
| AD | 1.486 | 0.082 | 1.513 | 0.076 | 1.795 | 0.157 | 0.115 | 0.178 | 0.523 | -0.073 | 0.386 | 0.156 | 0.024 | -0.082 | 0.007 | 0.030 | 0.001 |
| R VDC - R Medial geniculate nucleus | | | | | | | | | | | | | | | | | |
| FA | 0.253 | 0.022 | 0.247 | 0.024 | -2.471 | -0.071 | 0.114 | 0.537 | 0.964 | -0.297 | 0.156 | -0.124 | 0.015 | 0.084 | 0.007 | 0.201 | 0.040 |
| MD | 1.245 | 0.061 | 1.245 | 0.066 | -0.001 | -0.019 | 0.117 | 0.872 | 0.943 | -0.251 | 0.213 | 0.000 | 0.000 | -0.011 | 0.000 | -0.072 | 0.005 |
| RD | 1.072 | 0.055 | 1.078 | 0.059 | 0.476 | 0.002 | 0.116 | 0.984 | 0.989 | -0.228 | 0.233 | 0.040 | 0.002 | -0.046 | 0.002 | -0.138 | 0.019 |
| AD | 1.590 | 0.088 | 1.580 | 0.094 | -0.646 | -0.043 | 0.117 | 0.715 | 0.913 | -0.275 | 0.190 | -0.051 | 0.003 | 0.034 | 0.001 | 0.022 | 0.000 |
| Vermis - L Inferior colliculus | | | | | | | | | | | | | | | | | |
| FA | 0.173 | 0.021 | 0.168 | 0.020 | -3.083 | -0.068 | 0.115 | 0.554 | 0.964 | -0.297 | 0.160 | -0.118 | 0.014 | 0.156 | 0.024 | 0.090 | 0.008 |
| MD | 1.198 | 0.069 | 1.199 | 0.073 | 0.028 | 0.018 | 0.116 | 0.878 | 0.943 | -0.213 | 0.248 | 0.002 | 0.000 | 0.116 | 0.014 | -0.079 | 0.006 |
| RD | 1.090 | 0.064 | 1.094 | 0.070 | 0.404 | 0.033 | 0.116 | 0.779 | 0.907 | -0.198 | 0.264 | 0.030 | 0.001 | 0.076 | 0.006 | -0.099 | 0.010 |
| AD | 1.416 | 0.089 | 1.408 | 0.088 | -0.551 | -0.007 | 0.115 | 0.949 | 0.960 | -0.237 | 0.222 | -0.041 | 0.002 | 0.166 | 0.027 | -0.039 | 0.002 |
| Vermis - L Cochlear nucleus | | | | | | | | | | | | | | | | | |
| FA | 0.175 | 0.028 | 0.169 | 0.032 | -3.225 | -0.069 | 0.116 | 0.550 | 0.964 | -0.300 | 0.161 | -0.085 | 0.007 | -0.010 | 0.000 | 0.127 | 0.016 |
| MD | 1.276 | 0.165 | 1.290 | 0.175 | 1.040 | 0.041 | 0.116 | 0.727 | 0.876 | -0.191 | 0.272 | 0.036 | 0.001 | 0.066 | 0.004 | -0.080 | 0.006 |
| RD | 1.160 | 0.158 | 1.178 | 0.174 | 1.529 | 0.052 | 0.116 | 0.658 | 0.812 | -0.180 | 0.283 | 0.048 | 0.002 | 0.064 | 0.004 | -0.091 | 0.008 |
| AD | 1.508 | 0.185 | 1.512 | 0.184 | 0.287 | 0.019 | 0.117 | 0.872 | 0.960 | -0.213 | 0.251 | 0.011 | 0.000 | 0.068 | 0.005 | -0.056 | 0.003 |
| Vermis - L Medial geniculate nucleus | | | | | | | | | | | | | | | | | |
| FA | 0.206 | 0.027 | 0.206 | 0.026 | -0.267 | 0.029 | 0.116 | 0.806 | 0.974 | -0.202 | 0.260 | -0.010 | 0.000 | 0.063 | 0.004 | 0.101 | 0.010 |
| MD | 1.149 | 0.058 | 1.163 | 0.057 | 1.205 | 0.146 | 0.115 | 0.208 | 0.432 | -0.083 | 0.375 | 0.110 | 0.012 | 0.115 | 0.013 | -0.040 | 0.002 |
| RD | 1.021 | 0.049 | 1.033 | 0.049 | 1.240 | 0.142 | 0.115 | 0.221 | 0.451 | -0.087 | 0.371 | 0.118 | 0.014 | 0.097 | 0.009 | -0.076 | 0.006 |
| AD | 1.406 | 0.092 | 1.423 | 0.090 | 1.155 | 0.124 | 0.115 | 0.286 | 0.558 | -0.106 | 0.354 | 0.082 | 0.007 | 0.113 | 0.013 | 0.006 | 0.000 |
| Vermis - R Inferior colliculus | | | | | | | | | | | | | | | | | |
| FA | 0.175 | 0.023 | 0.171 | 0.019 | -1.891 | -0.056 | 0.116 | 0.630 | 0.974 | -0.288 | 0.175 | -0.076 | 0.006 | 0.052 | 0.003 | 0.062 | 0.004 |
| MD | 1.211 | 0.078 | 1.215 | 0.073 | 0.345 | 0.027 | 0.115 | 0.813 | 0.927 | -0.202 | 0.257 | 0.026 | 0.001 | 0.103 | 0.011 | -0.138 | 0.019 |
| RD | 1.100 | 0.074 | 1.106 | 0.070 | 0.555 | 0.036 | 0.115 | 0.754 | 0.907 | -0.193 | 0.266 | 0.039 | 0.002 | 0.084 | 0.007 | -0.144 | 0.021 |
| AD | 1.433 | 0.096 | 1.433 | 0.087 | 0.022 | 0.011 | 0.115 | 0.928 | 0.960 | -0.219 | 0.240 | 0.002 | 0.000 | 0.122 | 0.015 | -0.117 | 0.014 |
| Vermis - R Medial geniculate nucleus | | | | | | | | | | | | | | | | | |
| FA | 0.212 | 0.025 | 0.207 | 0.025 | -2.272 | -0.075 | 0.116 | 0.519 | 0.964 | -0.307 | 0.156 | -0.088 | 0.008 | 0.074 | 0.005 | 0.006 | 0.000 |
| MD | 1.164 | 0.063 | 1.178 | 0.059 | 1.204 | 0.065 | 0.114 | 0.571 | 0.792 | -0.163 | 0.293 | 0.107 | 0.011 | -0.039 | 0.002 | -0.196 | 0.039 |
| RD | 1.030 | 0.056 | 1.046 | 0.051 | 1.543 | 0.092 | 0.114 | 0.422 | 0.681 | -0.135 | 0.318 | 0.139 | 0.019 | -0.065 | 0.004 | -0.203 | 0.041 |
| AD | 1.433 | 0.093 | 1.443 | 0.091 | 0.717 | 0.023 | 0.116 | 0.842 | 0.957 | -0.207 | 0.253 | 0.052 | 0.003 | -0.002 | 0.000 | -0.156 | 0.024 |
| Midbrain - L Inferior colliculus | | | | | | | | | | | | | | | | | |
| FA | 0.207 | 0.032 | 0.201 | 0.028 | -3.106 | -0.072 | 0.116 | 0.537 | 0.964 | -0.302 | 0.158 | -0.100 | 0.010 | 0.043 | 0.002 | 0.129 | 0.017 |
| MD | 1.239 | 0.080 | 1.251 | 0.068 | 0.984 | 0.075 | 0.113 | 0.510 | 0.741 | -0.151 | 0.301 | 0.078 | 0.006 | 0.119 | 0.014 | -0.206 | 0.042 |
| RD | 1.105 | 0.079 | 1.120 | 0.066 | 1.397 | 0.088 | 0.113 | 0.436 | 0.681 | -0.136 | 0.313 | 0.101 | 0.010 | 0.098 | 0.010 | -0.236 | 0.056 |
| AD | 1.508 | 0.107 | 1.514 | 0.093 | 0.380 | 0.039 | 0.115 | 0.737 | 0.930 | -0.191 | 0.268 | 0.027 | 0.001 | 0.121 | 0.015 | -0.116 | 0.014 |
| Midbrain - L Cochlear nucleus | | | | | | | | | | | | | | | | | |
| FA | 0.213 | 0.028 | 0.211 | 0.026 | -0.891 | -0.020 | 0.117 | 0.863 | 0.974 | -0.253 | 0.212 | -0.032 | 0.001 | 0.028 | 0.001 | 0.037 | 0.001 |
| MD | 1.208 | 0.052 | 1.225 | 0.068 | 1.462 | 0.132 | 0.116 | 0.260 | 0.474 | -0.099 | 0.363 | 0.127 | 0.016 | -0.002 | 0.000 | -0.040 | 0.002 |
| RD | 1.073 | 0.051 | 1.090 | 0.068 | 1.531 | 0.118 | 0.116 | 0.312 | 0.570 | -0.113 | 0.349 | 0.118 | 0.014 | -0.011 | 0.000 | -0.047 | 0.002 |
| AD | 1.477 | 0.078 | 1.497 | 0.083 | 1.361 | 0.124 | 0.116 | 0.290 | 0.558 | -0.107 | 0.355 | 0.113 | 0.013 | 0.011 | 0.000 | -0.020 | 0.000 |
| Midbrain - L Medial geniculate nucleus | | | | | | | | | | | | | | | | | |
| FA | 0.240 | 0.026 | 0.235 | 0.024 | -2.048 | -0.038 | 0.115 | 0.744 | 0.974 | -0.266 | 0.191 | -0.091 | 0.008 | 0.184 | 0.034 | 0.058 | 0.003 |
| MD | 1.241 | 0.057 | 1.269 | 0.051 | 2.229 | 0.266 | 0.111 | **0.018** | 0.206 | 0.046 | 0.486 | 0.236 | 0.056 | 0.137 | 0.019 | -0.157 | 0.024 |
| RD | 1.081 | 0.053 | 1.109 | 0.047 | 2.638 | 0.270 | 0.111 | **0.017** | 0.167 | 0.049 | 0.491 | 0.262 | 0.069 | 0.064 | 0.004 | -0.176 | 0.031 |
| AD | 1.563 | 0.086 | 1.589 | 0.078 | 1.664 | 0.197 | 0.112 | 0.083 | 0.447 | -0.026 | 0.420 | 0.147 | 0.022 | 0.195 | 0.038 | -0.095 | 0.009 |
| Midbrain - R Inferior colliculus | | | | | | | | | | | | | | | | | |
| FA | 0.207 | 0.028 | 0.209 | 0.027 | 0.792 | 0.026 | 0.117 | 0.823 | 0.974 | -0.206 | 0.259 | 0.027 | 0.001 | -0.048 | 0.002 | 0.042 | 0.002 |
| MD | 1.231 | 0.067 | 1.234 | 0.057 | 0.269 | -0.001 | 0.115 | 0.995 | 0.995 | -0.230 | 0.228 | 0.025 | 0.001 | 0.036 | 0.001 | -0.180 | 0.033 |
| RD | 1.096 | 0.064 | 1.098 | 0.053 | 0.122 | -0.014 | 0.115 | 0.903 | 0.988 | -0.242 | 0.214 | 0.011 | 0.000 | 0.053 | 0.003 | -0.189 | 0.036 |
| AD | 1.501 | 0.092 | 1.508 | 0.088 | 0.485 | 0.016 | 0.116 | 0.888 | 0.960 | -0.215 | 0.247 | 0.038 | 0.001 | 0.008 | 0.000 | -0.127 | 0.016 |
| Midbrain - R Heschl's gyrus | | | | | | | | | | | | | | | | | |
| FA | 0.264 | 0.016 | 0.254 | 0.017 | -4.026 | -0.230 | 0.112 | **0.044** | 0.849 | -0.453 | -0.007 | -0.287 | 0.083 | 0.174 | 0.030 | 0.175 | 0.031 |
| MD | 1.221 | 0.062 | 1.227 | 0.045 | 0.526 | 0.055 | 0.116 | 0.637 | 0.822 | -0.177 | 0.287 | 0.059 | 0.003 | 0.029 | 0.001 | -0.084 | 0.007 |
| RD | 1.044 | 0.058 | 1.056 | 0.046 | 1.121 | 0.086 | 0.115 | 0.460 | 0.681 | -0.144 | 0.315 | 0.107 | 0.011 | -0.011 | 0.000 | -0.139 | 0.019 |
| AD | 1.574 | 0.076 | 1.570 | 0.063 | -0.263 | -0.004 | 0.117 | 0.970 | 0.970 | -0.236 | 0.228 | -0.028 | 0.001 | 0.082 | 0.007 | 0.019 | 0.000 |
| Midbrain - R Cochlear nucleus | | | | | | | | | | | | | | | | | |
| FA | 0.214 | 0.028 | 0.213 | 0.026 | -0.823 | -0.043 | 0.117 | 0.716 | 0.974 | -0.275 | 0.189 | -0.031 | 0.001 | -0.061 | 0.004 | 0.039 | 0.001 |
| MD | 1.208 | 0.052 | 1.231 | 0.065 | 1.871 | 0.159 | 0.115 | 0.171 | 0.395 | -0.070 | 0.388 | 0.168 | 0.028 | -0.092 | 0.009 | -0.005 | 0.000 |
| RD | 1.071 | 0.054 | 1.093 | 0.066 | 2.063 | 0.155 | 0.115 | 0.184 | 0.414 | -0.075 | 0.385 | 0.159 | 0.025 | -0.068 | 0.005 | -0.014 | 0.000 |
| AD | 1.483 | 0.072 | 1.507 | 0.079 | 1.593 | 0.128 | 0.115 | 0.272 | 0.558 | -0.102 | 0.357 | 0.140 | 0.020 | -0.110 | 0.012 | 0.012 | 0.000 |
| Midbrain - R Medial geniculate nucleus | | | | | | | | | | | | | | | | | |
| FA | 0.239 | 0.026 | 0.238 | 0.022 | -0.436 | 0.021 | 0.116 | 0.855 | 0.974 | -0.209 | 0.252 | -0.021 | 0.000 | 0.058 | 0.003 | 0.129 | 0.017 |
| MD | 1.251 | 0.048 | 1.258 | 0.052 | 0.577 | 0.050 | 0.114 | 0.666 | 0.822 | -0.178 | 0.277 | 0.065 | 0.004 | 0.072 | 0.005 | -0.196 | 0.039 |
| RD | 1.090 | 0.046 | 1.096 | 0.045 | 0.605 | 0.034 | 0.113 | 0.767 | 0.907 | -0.191 | 0.259 | 0.067 | 0.004 | 0.051 | 0.003 | -0.253 | 0.064 |
| AD | 1.572 | 0.077 | 1.581 | 0.082 | 0.537 | 0.057 | 0.116 | 0.625 | 0.838 | -0.174 | 0.288 | 0.048 | 0.002 | 0.081 | 0.007 | -0.090 | 0.008 |
| Pons - L Inferior colliculus | | | | | | | | | | | | | | | | | |
| FA | 0.208 | 0.026 | 0.205 | 0.023 | -1.622 | -0.031 | 0.116 | 0.789 | 0.974 | -0.262 | 0.200 | -0.065 | 0.004 | 0.065 | 0.004 | 0.106 | 0.011 |
| MD | 1.308 | 0.106 | 1.307 | 0.094 | -0.109 | -0.002 | 0.116 | 0.988 | 0.995 | -0.232 | 0.229 | -0.007 | 0.000 | 0.095 | 0.009 | -0.095 | 0.009 |
| RD | 1.167 | 0.107 | 1.168 | 0.092 | 0.073 | 0.002 | 0.116 | 0.989 | 0.989 | -0.229 | 0.233 | 0.004 | 0.000 | 0.073 | 0.005 | -0.108 | 0.012 |
| AD | 1.591 | 0.116 | 1.585 | 0.109 | -0.376 | -0.007 | 0.116 | 0.950 | 0.960 | -0.238 | 0.223 | -0.025 | 0.001 | 0.121 | 0.015 | -0.064 | 0.004 |
| Pons - L Cochlear nucleus | | | | | | | | | | | | | | | | | |
| FA | 0.215 | 0.029 | 0.210 | 0.029 | -2.282 | -0.057 | 0.116 | 0.626 | 0.974 | -0.289 | 0.175 | -0.079 | 0.006 | 0.069 | 0.005 | 0.053 | 0.003 |
| MD | 1.268 | 0.086 | 1.315 | 0.097 | 3.743 | 0.230 | 0.117 | 0.054 | 0.233 | -0.004 | 0.464 | 0.228 | 0.052 | -0.060 | 0.004 | -0.030 | 0.001 |
| RD | 1.137 | 0.105 | 1.172 | 0.101 | 3.028 | 0.161 | 0.119 | 0.179 | 0.413 | -0.076 | 0.399 | 0.155 | 0.024 | -0.037 | 0.001 | -0.010 | 0.000 |
| AD | 1.567 | 0.118 | 1.619 | 0.120 | 3.310 | 0.259 | 0.113 | **0.024** | 0.294 | 0.035 | 0.483 | 0.196 | 0.038 | 0.099 | 0.010 | 0.062 | 0.004 |
| Pons - L Medial geniculate nucleus | | | | | | | | | | | | | | | | | |
| FA | 0.234 | 0.026 | 0.231 | 0.023 | -1.138 | -0.006 | 0.116 | 0.962 | 0.986 | -0.235 | 0.224 | -0.050 | 0.003 | 0.154 | 0.024 | 0.033 | 0.001 |
| MD | 1.287 | 0.080 | 1.296 | 0.050 | 0.704 | 0.082 | 0.116 | 0.481 | 0.718 | -0.149 | 0.314 | 0.069 | 0.005 | 0.062 | 0.004 | -0.051 | 0.003 |
| RD | 1.127 | 0.080 | 1.137 | 0.051 | 0.893 | 0.074 | 0.117 | 0.529 | 0.729 | -0.158 | 0.306 | 0.076 | 0.006 | 0.008 | 0.000 | -0.058 | 0.003 |
| AD | 1.607 | 0.098 | 1.614 | 0.069 | 0.438 | 0.076 | 0.116 | 0.512 | 0.764 | -0.154 | 0.306 | 0.041 | 0.002 | 0.130 | 0.017 | -0.028 | 0.001 |
| Pons - R Inferior colliculus | | | | | | | | | | | | | | | | | |
| FA | 0.209 | 0.024 | 0.210 | 0.023 | 0.736 | 0.030 | 0.117 | 0.797 | 0.974 | -0.202 | 0.262 | 0.030 | 0.001 | -0.060 | 0.004 | 0.061 | 0.004 |
| MD | 1.267 | 0.068 | 1.277 | 0.060 | 0.742 | 0.065 | 0.115 | 0.575 | 0.792 | -0.164 | 0.294 | 0.069 | 0.005 | 0.077 | 0.006 | -0.154 | 0.024 |
| RD | 1.128 | 0.067 | 1.136 | 0.059 | 0.673 | 0.051 | 0.115 | 0.658 | 0.812 | -0.178 | 0.280 | 0.056 | 0.003 | 0.084 | 0.007 | -0.162 | 0.026 |
| AD | 1.545 | 0.084 | 1.558 | 0.079 | 0.842 | 0.073 | 0.116 | 0.532 | 0.782 | -0.158 | 0.303 | 0.074 | 0.006 | 0.051 | 0.003 | -0.110 | 0.012 |
| Pons - R Heschl's gyrus | | | | | | | | | | | | | | | | | |
| FA | 0.253 | 0.018 | 0.243 | 0.016 | -3.905 | -0.210 | 0.111 | 0.062 | 0.849 | -0.431 | 0.011 | -0.264 | 0.070 | 0.146 | 0.021 | 0.197 | 0.039 |
| MD | 1.251 | 0.067 | 1.251 | 0.055 | -0.020 | -0.036 | 0.116 | 0.758 | 0.903 | -0.268 | 0.196 | -0.002 | 0.000 | -0.063 | 0.004 | -0.075 | 0.006 |
| RD | 1.081 | 0.063 | 1.088 | 0.050 | 0.679 | 0.014 | 0.115 | 0.905 | 0.988 | -0.216 | 0.244 | 0.062 | 0.004 | -0.099 | 0.010 | -0.126 | 0.016 |
| AD | 1.592 | 0.083 | 1.577 | 0.074 | -0.968 | -0.102 | 0.116 | 0.384 | 0.637 | -0.334 | 0.130 | -0.092 | 0.008 | -0.004 | 0.000 | 0.007 | 0.000 |
| Pons - R Cochlear nucleus | | | | | | | | | | | | | | | | | |
| FA | 0.221 | 0.032 | 0.213 | 0.027 | -3.566 | -0.122 | 0.116 | 0.295 | 0.964 | -0.353 | 0.109 | -0.125 | 0.016 | 0.019 | 0.000 | 0.053 | 0.003 |
| MD | 1.259 | 0.105 | 1.295 | 0.097 | 2.882 | 0.204 | 0.114 | 0.079 | 0.268 | -0.024 | 0.432 | 0.166 | 0.028 | 0.011 | 0.000 | 0.075 | 0.006 |
| RD | 1.113 | 0.111 | 1.152 | 0.099 | 3.494 | 0.205 | 0.115 | 0.078 | 0.283 | -0.023 | 0.433 | 0.172 | 0.030 | 0.007 | 0.000 | 0.057 | 0.003 |
| AD | 1.551 | 0.105 | 1.582 | 0.103 | 2.004 | 0.180 | 0.115 | 0.121 | 0.477 | -0.049 | 0.408 | 0.137 | 0.019 | 0.019 | 0.000 | 0.103 | 0.011 |
| Pons - R Medial geniculate nucleus | | | | | | | | | | | | | | | | | |
| FA | 0.235 | 0.023 | 0.232 | 0.020 | -1.425 | -0.049 | 0.116 | 0.674 | 0.974 | -0.280 | 0.182 | -0.073 | 0.005 | 0.041 | 0.002 | 0.095 | 0.009 |
| MD | 1.274 | 0.050 | 1.283 | 0.051 | 0.721 | 0.065 | 0.115 | 0.574 | 0.792 | -0.164 | 0.293 | 0.083 | 0.007 | 0.041 | 0.002 | -0.182 | 0.033 |
| RD | 1.113 | 0.048 | 1.124 | 0.045 | 0.958 | 0.078 | 0.114 | 0.498 | 0.695 | -0.149 | 0.304 | 0.106 | 0.011 | 0.022 | 0.001 | -0.217 | 0.047 |
| AD | 1.595 | 0.072 | 1.601 | 0.076 | 0.389 | 0.036 | 0.116 | 0.758 | 0.943 | -0.195 | 0.267 | 0.038 | 0.001 | 0.056 | 0.003 | -0.103 | 0.011 |
| Medulla - L Cochlear nucleus | | | | | | | | | | | | | | | | | |
| FA | 0.193 | 0.028 | 0.184 | 0.029 | -4.432 | -0.145 | 0.115 | 0.213 | 0.964 | -0.374 | 0.085 | -0.137 | 0.019 | -0.043 | 0.002 | 0.090 | 0.008 |
| MD | 1.251 | 0.121 | 1.338 | 0.127 | 6.942 | 0.333 | 0.110 | **0.003** | 0.206 | 0.113 | 0.552 | 0.305 | 0.093 | 0.056 | 0.003 | -0.101 | 0.010 |
| RD | 1.128 | 0.121 | 1.213 | 0.130 | 7.573 | 0.323 | 0.110 | **0.005** | 0.167 | 0.103 | 0.542 | 0.296 | 0.088 | 0.063 | 0.004 | -0.107 | 0.011 |
| AD | 1.498 | 0.129 | 1.588 | 0.128 | 5.992 | 0.333 | 0.110 | **0.003** | 0.275 | 0.113 | 0.553 | 0.306 | 0.094 | 0.041 | 0.002 | -0.083 | 0.007 |
| Medulla - L Medial geniculate nucleus | | | | | | | | | | | | | | | | | |
| FA | 0.224 | 0.027 | 0.224 | 0.028 | -0.112 | 0.012 | 0.116 | 0.920 | 0.986 | -0.220 | 0.243 | -0.004 | 0.000 | 0.100 | 0.010 | -0.053 | 0.003 |
| MD | 1.178 | 0.071 | 1.220 | 0.063 | 3.541 | 0.282 | 0.110 | **0.012** | 0.206 | 0.063 | 0.502 | 0.283 | 0.080 | 0.041 | 0.002 | -0.198 | 0.039 |
| RD | 1.037 | 0.065 | 1.075 | 0.060 | 3.621 | 0.264 | 0.112 | **0.021** | 0.167 | 0.042 | 0.485 | 0.269 | 0.073 | 0.006 | 0.000 | -0.171 | 0.029 |
| AD | 1.461 | 0.099 | 1.511 | 0.090 | 3.428 | 0.249 | 0.111 | **0.028** | 0.294 | 0.028 | 0.470 | 0.242 | 0.059 | 0.080 | 0.006 | -0.193 | 0.037 |
| Medulla - R Inferior colliculus | | | | | | | | | | | | | | | | | |
| FA | 0.201 | 0.025 | 0.196 | 0.026 | -2.363 | -0.070 | 0.116 | 0.549 | 0.964 | -0.300 | 0.161 | -0.084 | 0.007 | -0.023 | 0.001 | 0.137 | 0.019 |
| MD | 1.235 | 0.072 | 1.271 | 0.067 | 2.866 | 0.254 | 0.117 | **0.033** | 0.206 | 0.021 | 0.487 | 0.230 | 0.053 | 0.041 | 0.002 | -0.099 | 0.010 |
| RD | 1.132 | 0.122 | 1.152 | 0.110 | 1.783 | 0.026 | 0.115 | 0.819 | 0.944 | -0.202 | 0.255 | 0.082 | 0.007 | -0.118 | 0.014 | -0.144 | 0.021 |
| AD | 1.531 | 0.152 | 1.544 | 0.127 | 0.852 | -0.008 | 0.115 | 0.942 | 0.960 | -0.238 | 0.221 | 0.045 | 0.002 | -0.130 | 0.017 | -0.100 | 0.010 |
| Medulla - R Cochlear nucleus | | | | | | | | | | | | | | | | | |
| FA | 0.193 | 0.030 | 0.179 | 0.032 | -7.369 | -0.208 | 0.114 | 0.071 | 0.849 | -0.434 | 0.018 | -0.205 | 0.042 | -0.032 | 0.001 | 0.134 | 0.018 |
| MD | 1.263 | 0.106 | 1.301 | 0.114 | 3.054 | 0.153 | 0.117 | 0.194 | 0.425 | -0.079 | 0.385 | 0.160 | 0.025 | 0.123 | 0.015 | -0.247 | 0.061 |
| RD | 1.136 | 0.108 | 1.202 | 0.140 | 5.821 | 0.274 | 0.111 | **0.016** | 0.167 | 0.052 | 0.496 | 0.225 | 0.051 | 0.141 | 0.020 | -0.071 | 0.005 |
| AD | 1.517 | 0.113 | 1.543 | 0.114 | 1.700 | 0.106 | 0.118 | 0.373 | 0.627 | -0.130 | 0.342 | 0.105 | 0.011 | 0.130 | 0.017 | -0.199 | 0.040 |
| Medulla - R Medial geniculate nucleus | | | | | | | | | | | | | | | | | |
| FA | 0.228 | 0.030 | 0.216 | 0.026 | -5.058 | -0.182 | 0.114 | 0.116 | 0.964 | -0.409 | 0.046 | -0.189 | 0.036 | 0.010 | 0.000 | 0.118 | 0.014 |
| MD | 1.189 | 0.064 | 1.218 | 0.057 | 2.451 | 0.217 | 0.113 | 0.059 | 0.233 | -0.009 | 0.443 | 0.220 | 0.048 | 0.005 | 0.000 | -0.133 | 0.018 |
| RD | 1.042 | 0.057 | 1.078 | 0.057 | 3.386 | 0.269 | 0.111 | **0.018** | 0.167 | 0.048 | 0.491 | 0.276 | 0.076 | -0.002 | 0.000 | -0.168 | 0.028 |
| AD | 1.482 | 0.099 | 1.499 | 0.080 | 1.134 | 0.092 | 0.116 | 0.432 | 0.694 | -0.140 | 0.324 | 0.090 | 0.008 | 0.013 | 0.000 | -0.052 | 0.003 |
| L Inferior colliculus - L Cochlear nucleus | | | | | | | | | | | | | | | | | |
| FA | 0.185 | 0.016 | 0.183 | 0.019 | -0.768 | 0.009 | 0.116 | 0.941 | 0.986 | -0.221 | 0.239 | -0.036 | 0.001 | 0.064 | 0.004 | 0.143 | 0.020 |
| MD | 1.336 | 0.115 | 1.404 | 0.194 | 5.100 | 0.127 | 0.114 | 0.267 | 0.480 | -0.099 | 0.354 | 0.177 | 0.031 | -0.163 | 0.027 | -0.108 | 0.012 |
| RD | 1.211 | 0.107 | 1.274 | 0.184 | 5.169 | 0.120 | 0.114 | 0.295 | 0.549 | -0.106 | 0.346 | 0.172 | 0.030 | -0.164 | 0.027 | -0.116 | 0.013 |
| AD | 1.585 | 0.133 | 1.665 | 0.218 | 4.994 | 0.138 | 0.114 | 0.231 | 0.558 | -0.089 | 0.364 | 0.182 | 0.033 | -0.159 | 0.025 | -0.093 | 0.009 |
| L Inferior colliculus - L Medial geniculate nucleus | | | | | | | | | | | | | | | | | |
| FA | 0.189 | 0.027 | 0.184 | 0.032 | -2.799 | -0.051 | 0.113 | 0.652 | 0.974 | -0.275 | 0.173 | -0.079 | 0.006 | -0.070 | 0.005 | 0.261 | 0.068 |
| MD | 1.325 | 0.127 | 1.356 | 0.126 | 2.387 | 0.112 | 0.113 | 0.324 | 0.543 | -0.113 | 0.338 | 0.115 | 0.013 | 0.106 | 0.011 | -0.206 | 0.042 |
| RD | 1.200 | 0.128 | 1.234 | 0.133 | 2.786 | 0.109 | 0.113 | 0.335 | 0.586 | -0.115 | 0.333 | 0.116 | 0.013 | 0.108 | 0.012 | -0.231 | 0.053 |
| AD | 1.574 | 0.134 | 1.602 | 0.122 | 1.778 | 0.110 | 0.115 | 0.342 | 0.584 | -0.119 | 0.338 | 0.102 | 0.010 | 0.093 | 0.009 | -0.137 | 0.019 |
| L Inferior colliculus - R Inferior colliculus | | | | | | | | | | | | | | | | | |
| FA | 0.183 | 0.034 | 0.180 | 0.033 | -1.744 | -0.034 | 0.117 | 0.773 | 0.974 | -0.266 | 0.199 | -0.044 | 0.002 | 0.024 | 0.001 | 0.040 | 0.002 |
| MD | 1.220 | 0.100 | 1.217 | 0.083 | -0.244 | -0.001 | 0.116 | 0.991 | 0.995 | -0.233 | 0.230 | -0.016 | 0.000 | 0.100 | 0.010 | -0.055 | 0.003 |
| RD | 1.104 | 0.093 | 1.103 | 0.079 | -0.102 | 0.005 | 0.116 | 0.967 | 0.989 | -0.226 | 0.236 | -0.006 | 0.000 | 0.093 | 0.009 | -0.066 | 0.004 |
| AD | 1.451 | 0.133 | 1.444 | 0.114 | -0.461 | -0.010 | 0.116 | 0.935 | 0.960 | -0.241 | 0.222 | -0.026 | 0.001 | 0.091 | 0.008 | -0.030 | 0.001 |
| L Heschl's gyrus - L Medial geniculate nucleus | | | | | | | | | | | | | | | | | |
| FA | 0.285 | 0.029 | 0.280 | 0.033 | -2.016 | -0.079 | 0.116 | 0.496 | 0.964 | -0.310 | 0.151 | -0.083 | 0.007 | 0.096 | 0.009 | -0.062 | 0.004 |
| MD | 1.185 | 0.049 | 1.208 | 0.068 | 1.975 | 0.171 | 0.115 | 0.141 | 0.343 | -0.058 | 0.401 | 0.168 | 0.028 | -0.058 | 0.003 | 0.009 | 0.000 |
| RD | 1.000 | 0.055 | 1.024 | 0.073 | 2.484 | 0.167 | 0.115 | 0.151 | 0.394 | -0.062 | 0.396 | 0.165 | 0.027 | -0.083 | 0.007 | 0.031 | 0.001 |
| AD | 1.555 | 0.063 | 1.576 | 0.084 | 1.319 | 0.127 | 0.116 | 0.279 | 0.558 | -0.104 | 0.358 | 0.120 | 0.014 | 0.002 | 0.000 | -0.033 | 0.001 |
| L Cochlear nucleus - R Cochlear nucleus | | | | | | | | | | | | | | | | | |
| FA | 0.185 | 0.036 | 0.169 | 0.050 | -8.686 | -0.134 | 0.113 | 0.239 | 0.964 | -0.360 | 0.091 | -0.158 | 0.025 | -0.022 | 0.000 | 0.218 | 0.048 |
| MD | 1.335 | 0.170 | 1.506 | 0.373 | 12.821 | 0.208 | 0.111 | 0.065 | 0.236 | -0.013 | 0.429 | 0.235 | 0.055 | 0.001 | 0.000 | -0.242 | 0.059 |
| RD | 1.210 | 0.173 | 1.380 | 0.372 | 14.060 | 0.205 | 0.111 | 0.068 | 0.278 | -0.016 | 0.426 | 0.233 | 0.054 | 0.001 | 0.000 | -0.247 | 0.061 |
| AD | 1.586 | 0.173 | 1.667 | 0.178 | 5.151 | 0.236 | 0.117 | **0.046** | 0.378 | 0.004 | 0.468 | 0.213 | 0.045 | 0.080 | 0.006 | -0.097 | 0.009 |
| L Medial geniculate nucleus - R Medial geniculate nucleus | | | | | | | | | | | | | | | | | |
| FA | 0.262 | 0.031 | 0.258 | 0.029 | -1.694 | -0.026 | 0.116 | 0.822 | 0.974 | -0.256 | 0.204 | -0.069 | 0.005 | 0.140 | 0.020 | 0.052 | 0.003 |
| MD | 1.245 | 0.072 | 1.276 | 0.065 | 2.518 | 0.193 | 0.113 | 0.092 | 0.274 | -0.032 | 0.419 | 0.210 | 0.044 | -0.008 | 0.000 | -0.171 | 0.029 |
| RD | 1.063 | 0.050 | 1.095 | 0.053 | 2.948 | 0.231 | 0.111 | **0.040** | 0.252 | 0.010 | 0.452 | 0.267 | 0.071 | -0.066 | 0.004 | -0.215 | 0.046 |
| AD | 1.607 | 0.127 | 1.638 | 0.108 | 1.949 | 0.130 | 0.115 | 0.265 | 0.558 | -0.100 | 0.359 | 0.125 | 0.016 | 0.047 | 0.002 | -0.105 | 0.011 |
| R Inferior colliculus - R Cochlear nucleus | | | | | | | | | | | | | | | | | |
| FA | 0.187 | 0.016 | 0.180 | 0.017 | -4.046 | -0.208 | 0.115 | 0.073 | 0.849 | -0.436 | 0.020 | -0.203 | 0.041 | 0.046 | 0.002 | 0.020 | 0.000 |
| MD | 1.320 | 0.147 | 1.414 | 0.204 | 7.155 | 0.195 | 0.112 | 0.087 | 0.270 | -0.029 | 0.418 | 0.224 | 0.050 | -0.213 | 0.045 | 0.026 | 0.001 |
| RD | 1.194 | 0.138 | 1.285 | 0.192 | 7.636 | 0.201 | 0.112 | 0.077 | 0.283 | -0.022 | 0.424 | 0.229 | 0.053 | -0.213 | 0.045 | 0.026 | 0.001 |
| AD | 1.571 | 0.165 | 1.672 | 0.230 | 6.426 | 0.183 | 0.113 | 0.107 | 0.477 | -0.041 | 0.407 | 0.213 | 0.045 | -0.210 | 0.044 | 0.026 | 0.001 |
| R Inferior colliculus - R Medial geniculate nucleus | | | | | | | | | | | | | | | | | |
| FA | 0.190 | 0.030 | 0.194 | 0.030 | 1.872 | 0.069 | 0.116 | 0.550 | 0.964 | -0.161 | 0.300 | 0.055 | 0.003 | -0.064 | 0.004 | 0.121 | 0.015 |
| MD | 1.343 | 0.135 | 1.316 | 0.117 | -1.992 | -0.121 | 0.116 | 0.297 | 0.524 | -0.351 | 0.109 | -0.100 | 0.010 | 0.042 | 0.002 | -0.102 | 0.010 |
| RD | 1.214 | 0.140 | 1.188 | 0.123 | -2.140 | -0.113 | 0.115 | 0.330 | 0.586 | -0.343 | 0.117 | -0.093 | 0.009 | 0.050 | 0.002 | -0.111 | 0.012 |
| AD | 1.602 | 0.133 | 1.574 | 0.113 | -1.767 | -0.130 | 0.116 | 0.264 | 0.558 | -0.361 | 0.100 | -0.109 | 0.012 | 0.022 | 0.001 | -0.074 | 0.005 |
| R Heschl's gyrus - R Medial geniculate nucleus | | | | | | | | | | | | | | | | | |
| FA | 0.278 | 0.026 | 0.280 | 0.029 | 0.452 | 0.076 | 0.115 | 0.512 | 0.964 | -0.154 | 0.305 | 0.020 | 0.000 | 0.138 | 0.019 | 0.068 | 0.005 |
| MD | 1.208 | 0.067 | 1.206 | 0.064 | -0.157 | 0.006 | 0.117 | 0.962 | 0.995 | -0.226 | 0.238 | -0.013 | 0.000 | 0.082 | 0.007 | -0.011 | 0.000 |
| RD | 1.024 | 0.069 | 1.022 | 0.069 | -0.215 | -0.016 | 0.117 | 0.891 | 0.988 | -0.249 | 0.217 | -0.015 | 0.000 | 0.025 | 0.001 | -0.031 | 0.001 |
| AD | 1.574 | 0.079 | 1.573 | 0.073 | -0.082 | 0.044 | 0.115 | 0.704 | 0.911 | -0.185 | 0.273 | -0.008 | 0.000 | 0.167 | 0.028 | 0.027 | 0.001 |

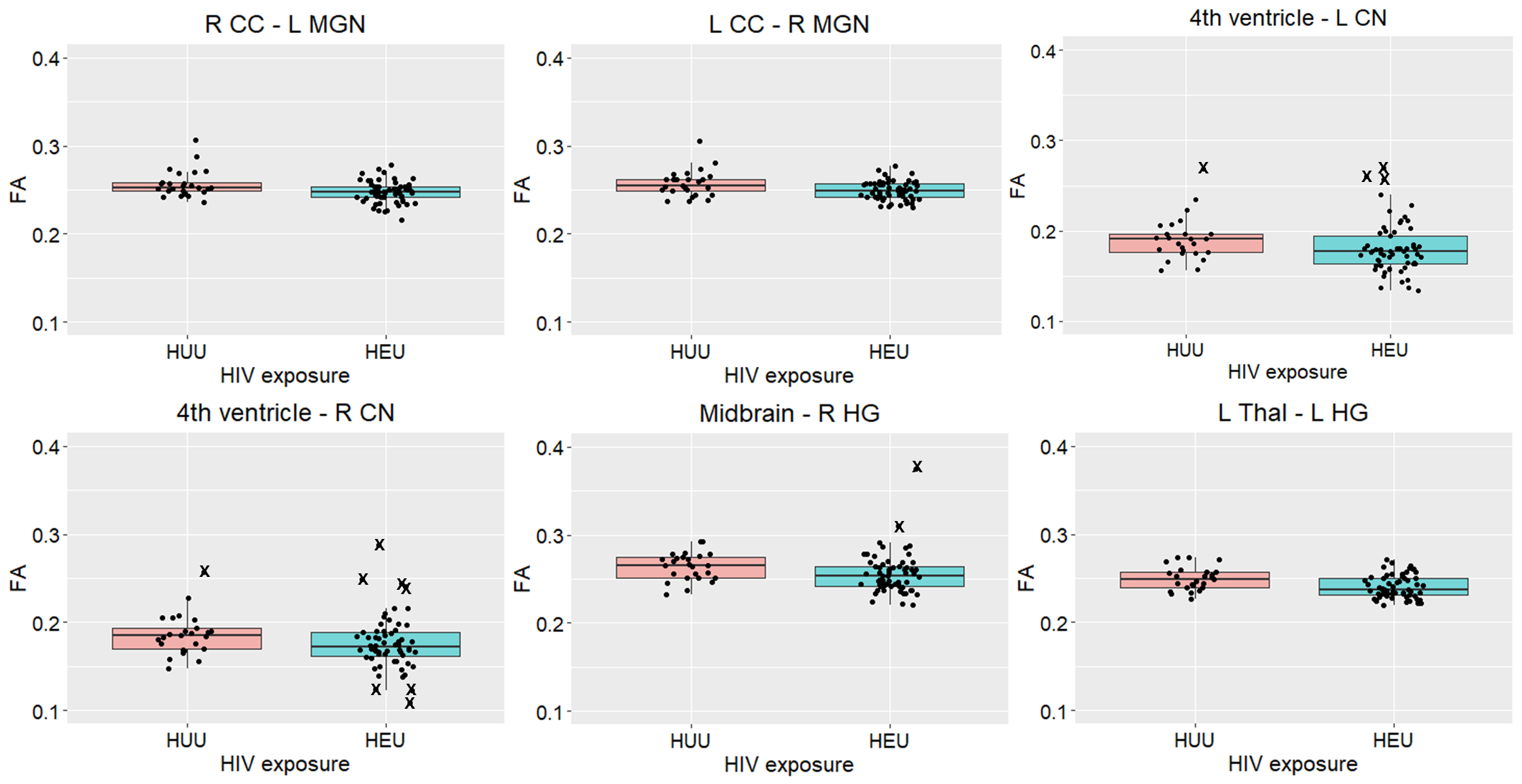

**Supplementary figure 4:** Boxplots showing fractional anisotropy (FA) according to HIV exposure groups for auditory tracts in which we observed groupwise FA differences. Influential outliers are shown as X’s.

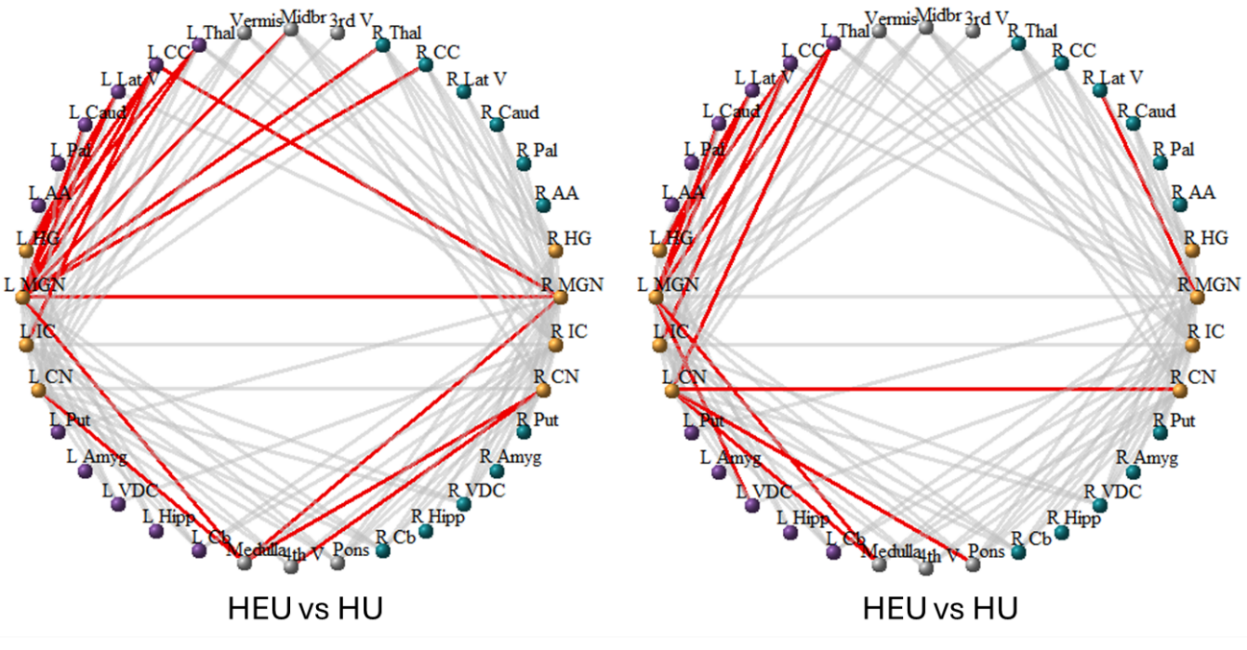

(a)

(b)

**Supplementary figure 5:** Auditory connections for which differences in (a) RD and (b) AD were observed for HEU compared to HU infants. Gray lines indicate auditory connections in which no differences were observed; Elevated RD and AD with unadjusted p values ≤ 0.05 = red.

*Cb=cerebellar cortex; Hipp=hippocampus; VDC=ventral diencephalon; Amyg=*Amygdala*; Put=putamen; AA= accumbens area; Pal= pallidum; Caud=caudate; Lat V=lateral ventricle; CC=cerebral cortex; Thal=thalamus; CN=cochlear nucleus; IC=inferior colliculus; MGN=medial geniculate nucleus; HG=Heschl’s gyrus; 3^rd^ V = 3^rd^ ventricle; 4^th^ V = 4^th^ ventricle.*

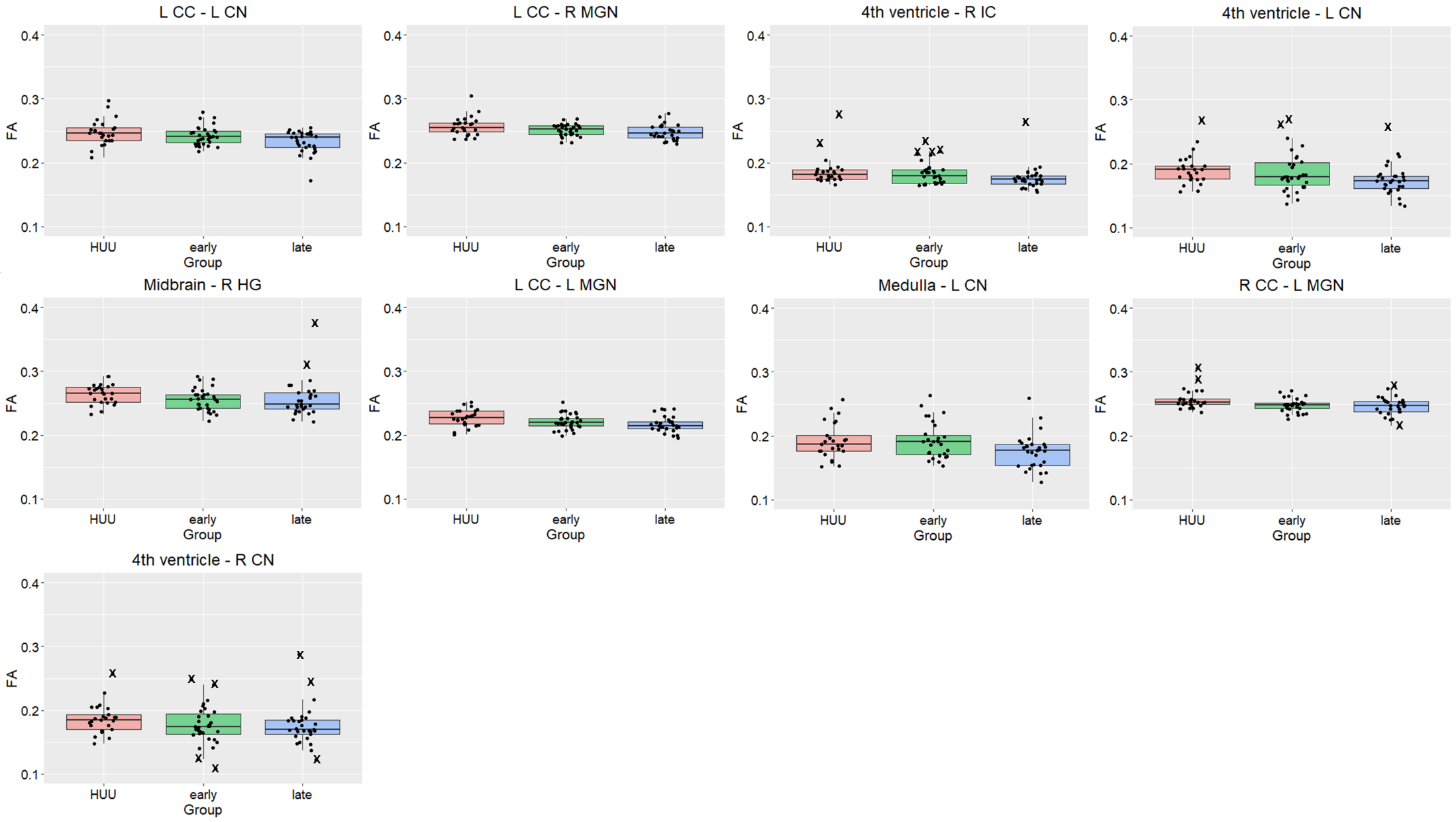

**Supplementary figure 6:** Boxplots showing fractional anisotropy (FA) according to ART exposure groups for auditory tracts in which we observed groupwise FA differences. Influential outliers are shown as X’s.

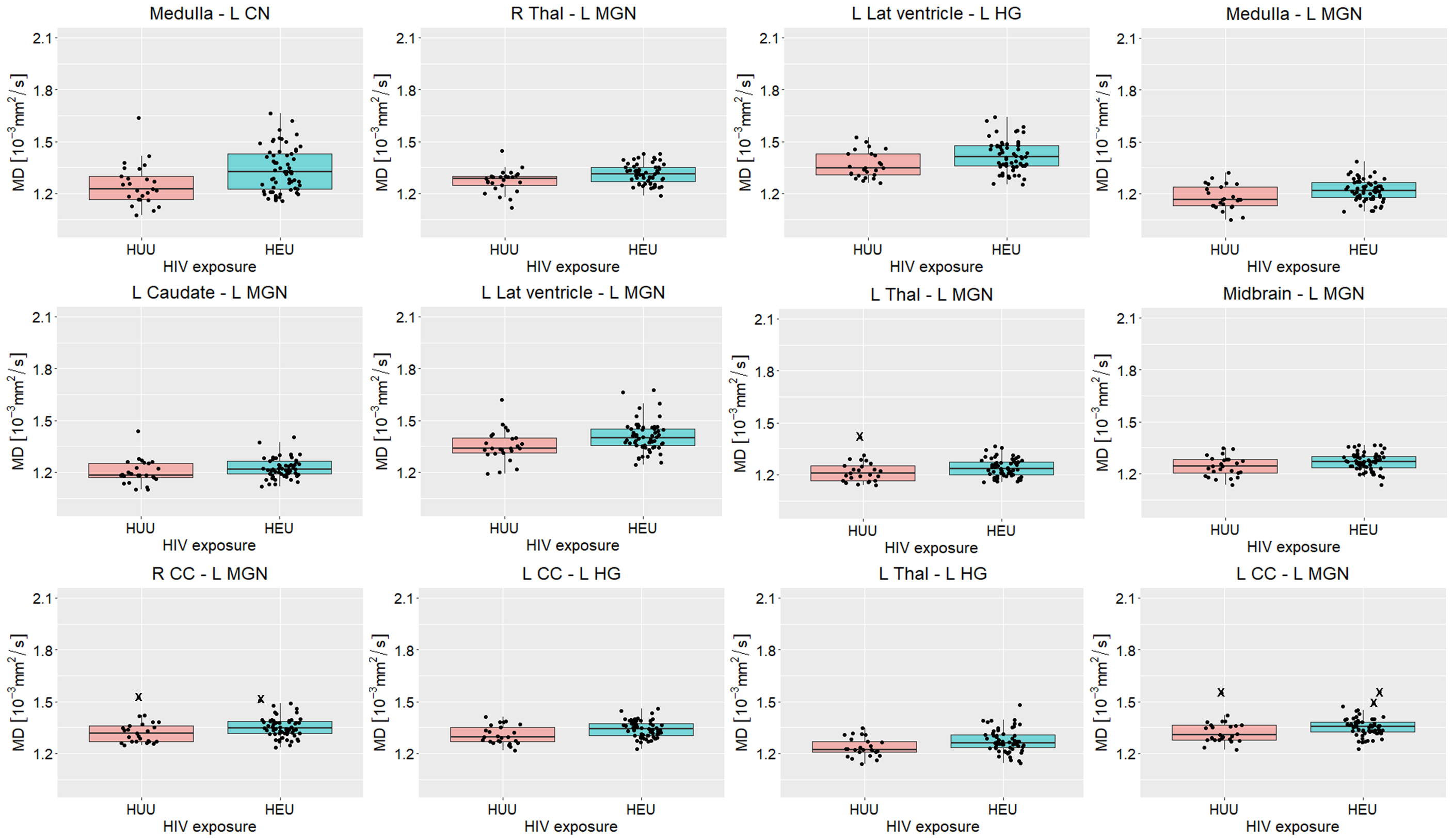

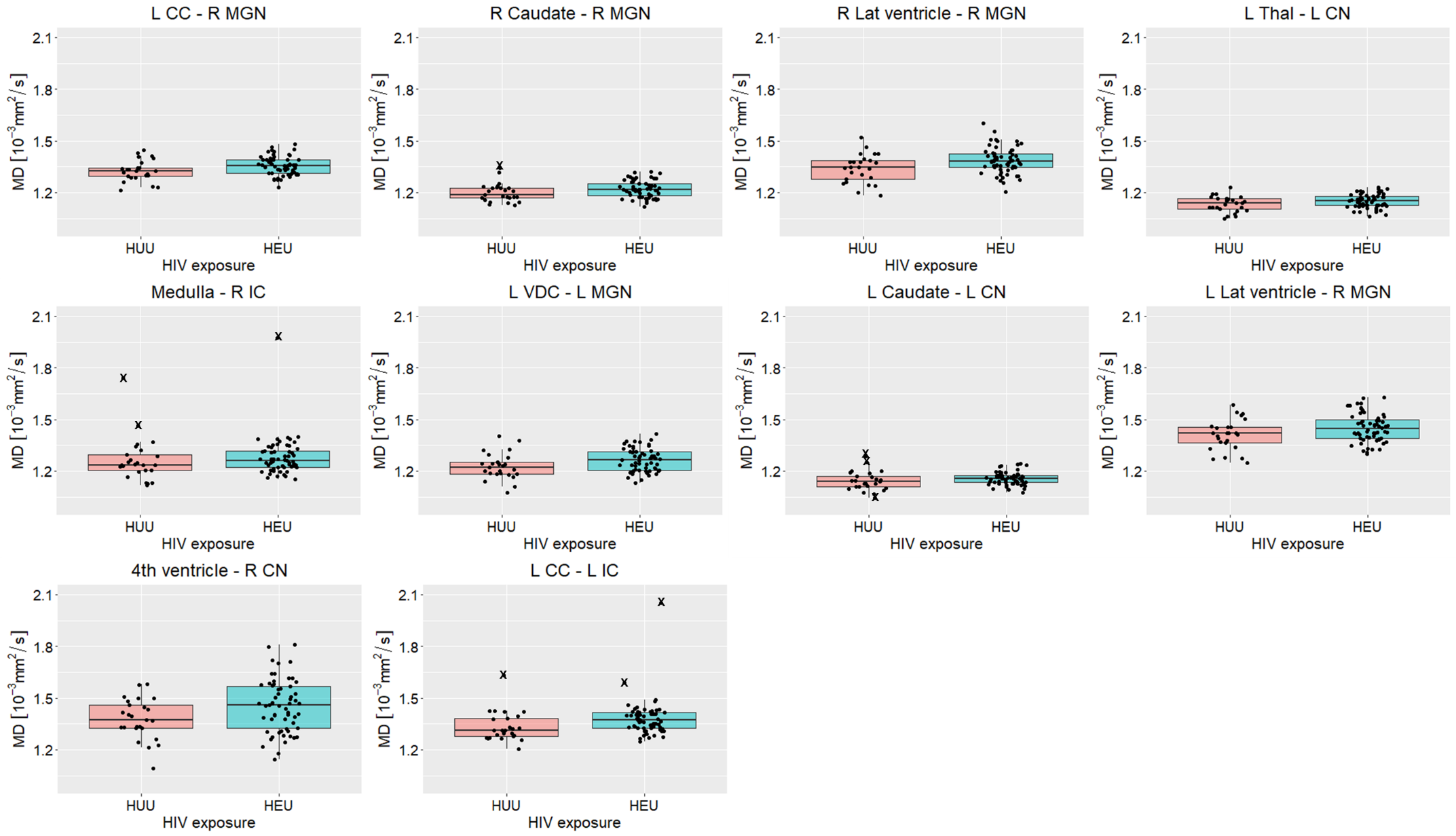
**Supplementary figure 7:** Boxplots showing mean diffusivity (MD) according to HIV exposure groups for auditory tracts in which we observed groupwise MD differences. Influential outliers are shown as X’s.

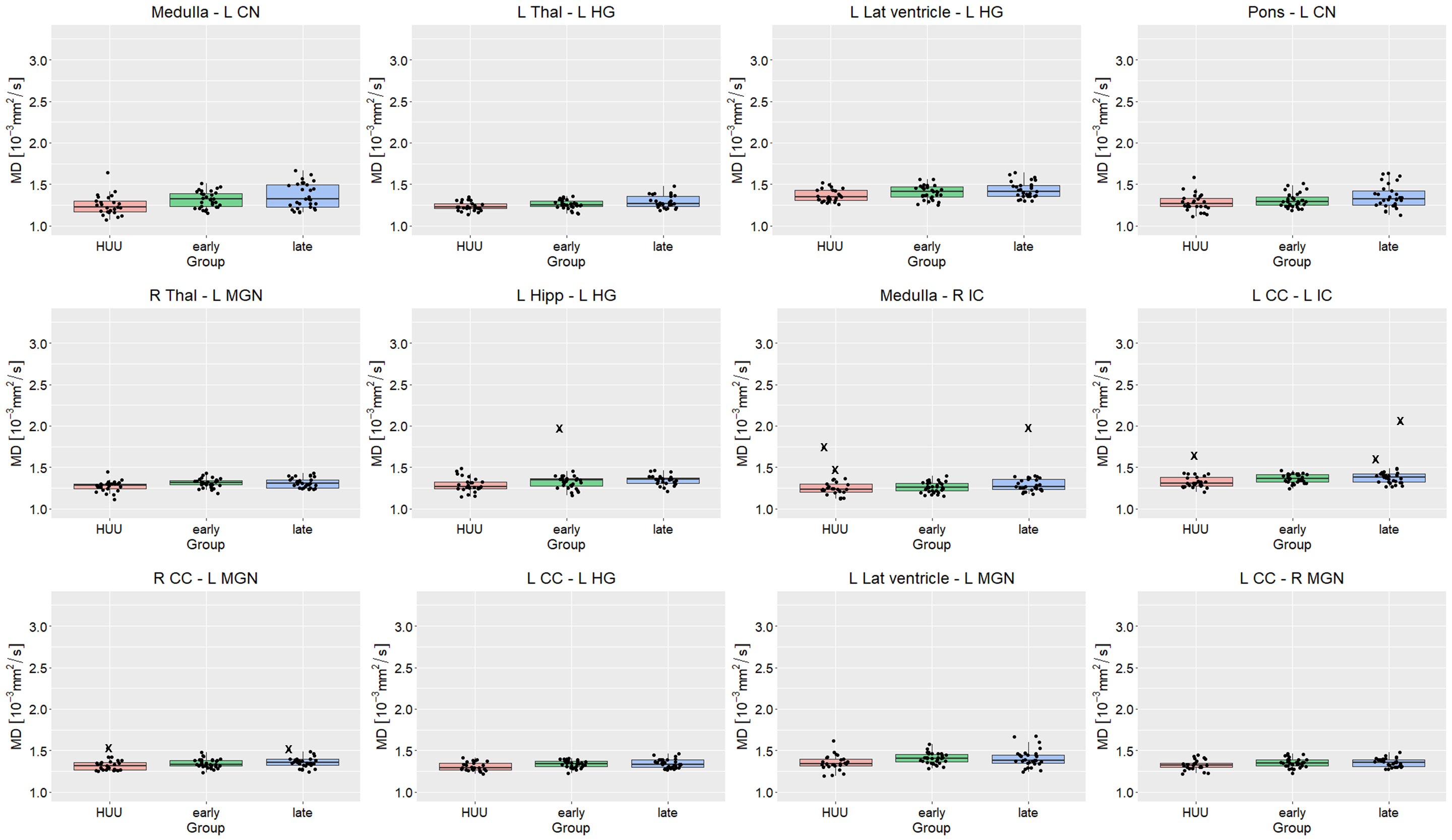

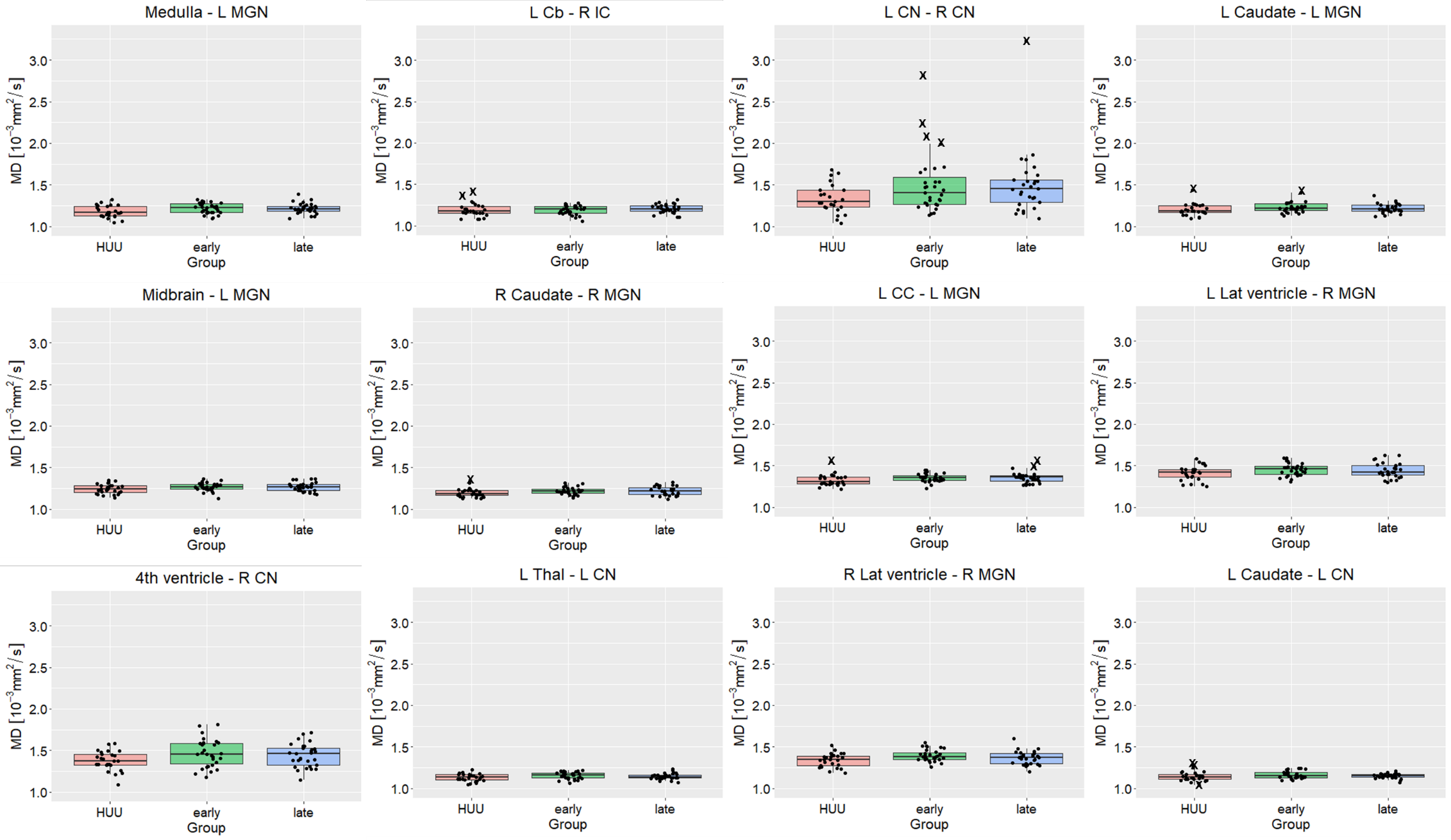

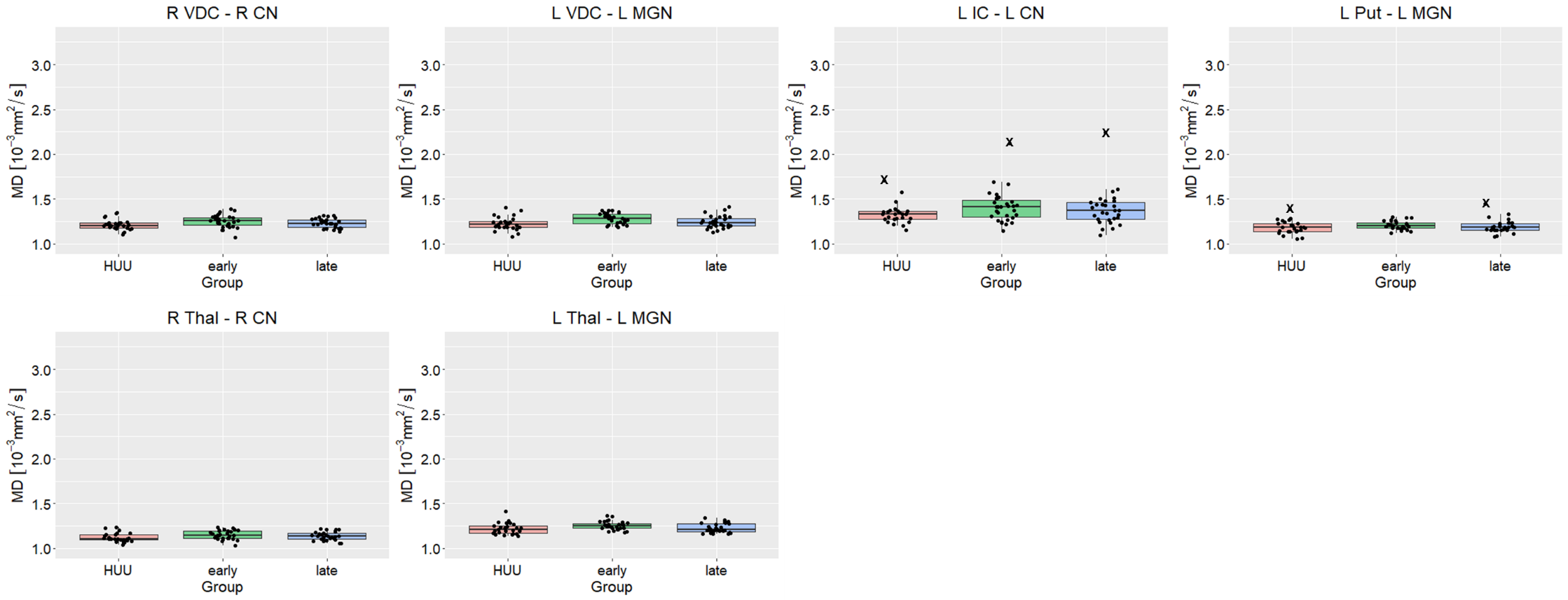

**Supplementary figure 8:** Boxplots showing mean diffusivity (MD) according to ART exposure groups for auditory tracts in which we observed groupwise MD differences. Influential outliers are shown as X’s.

**Supplementary table 2:** A summary of auditory WM tracts in which differences in FA, MD or both FA and MD have been seen in iHEU (All HEU) compared to iHU, or in iHEU-pre or iHEU-post compared to iHU. Bold indicates groups for which we observe differences in both FA and MD.

| **Auditory tracts** | **FA differences** | **MD differences** |
| --- | --- | --- |
| L CC - L Inferior colliculus |  | All HEU; iHEU-pre; iHEU-post |
| L CC - L Heschl's gyrus |  | All HEU; iHEU-post |
| L CC - L Cochlear nucleus | iHEU-post |  |
| L CC - L Medial geniculate nucleus | iHEU-post | All HEU; iHEU-pre |
| L CC - R Medial geniculate nucleus | **All HEU; iHEU-post** | **All HEU; iHEU-post** |
| L Lat Vent - L Heschl's gyrus |  | All HEU; iHEU-post |
| L Lat Vent - L Medial geniculate nucleus |  | All HEU; iHEU-pre; iHEU-post |
| L Lat Vent - R Medial geniculate nucleus |  | All HEU; iHEU-pre |
| L Cb - R Inferior colliculus |  | iHEU-post |
| L Thal - L Heschl's gyrus | **All HEU** | **All HEU**; iHEU-post |
| L Thal - L Cochlear nucleus |  | All HEU; iHEU-pre |
| L Thal - L Medial geniculate nucleus |  | All HEU; iHEU-pre |
| L Caud - L Cochlear nucleus |  | All HEU; iHEU-pre |
| L Caud - L Medial geniculate nucleus |  | All HEU; iHEU-pre; iHEU-post |
| L Put - L Medial geniculate nucleus |  | iHEU-pre |
| 4th Vent - L Cochlear nucleus | All HEU; iHEU-post |  |
| 4th Vent - R Inferior colliculus | iHEU-post |  |
| 4th Vent - R Cochlear nucleus | **All HEU**; iHEU-post | **All HEU**; iHEU-pre |
| L Hipp - L Heschl's gyrus |  | iHEU-post |
| L VDC - L Medial geniculate nucleus |  | All HEU; iHEU-pre |
| R CC - L Medial geniculate nucleus | **All HEU; iHEU-post** | **All HEU; iHEU-post** |
| R Lat Vent - R Medial geniculate nucleus |  | All HEU; iHEU-pre |
| R Thal - L Medial geniculate nucleus |  | All HEU; iHEU-pre; iHEU-post |
| R Thal - R Cochlear nucleus |  | iHEU-pre |
| R Caud - R Medial geniculate nucleus |  | All HEU; iHEU-pre |
| R VDC - R Cochlear nucleus |  | iHEU-pre |
| Midbrain - L Medial geniculate nucleus |  | All HEU; iHEU-pre |
| Midbrain - R Heschl's gyrus | All HEU; iHEU-post |  |
| Pons - L Cochlear nucleus |  | iHEU-post |
| Medulla - L Cochlear nucleus | **iHEU-post** | All HEU; iHEU-pre; **iHEU-post** |
| Medulla - L Medial geniculate nucleus |  | All HEU; iHEU-pre; iHEU-post |
| Medulla - R Inferior colliculus |  | All HEU; iHEU-post |
| L Inferior colliculus - L Cochlear nucleus |  | iHEU-pre |
| L Cochlear nucleus - R Cochlear nucleus |  | iHEU-post |

*Cb=cerebellar cortex; Hipp=hippocampus; VDC=ventral diencephalon; Amyg=*Amygdala*; Put=putamen; AA= accumbens area; Pal= pallidum; Caud=caudate; Lat V=lateral ventricle; CC=cerebral cortex; Thal=thalamus; CN=cochlear nucleus; IC=inferior colliculus; MGN=medial geniculate nucleus; HG=Heschl’s gyrus; 3^rd^ V = 3^rd^ ventricle; 4^th^ V = 4^th^ ventricle.*

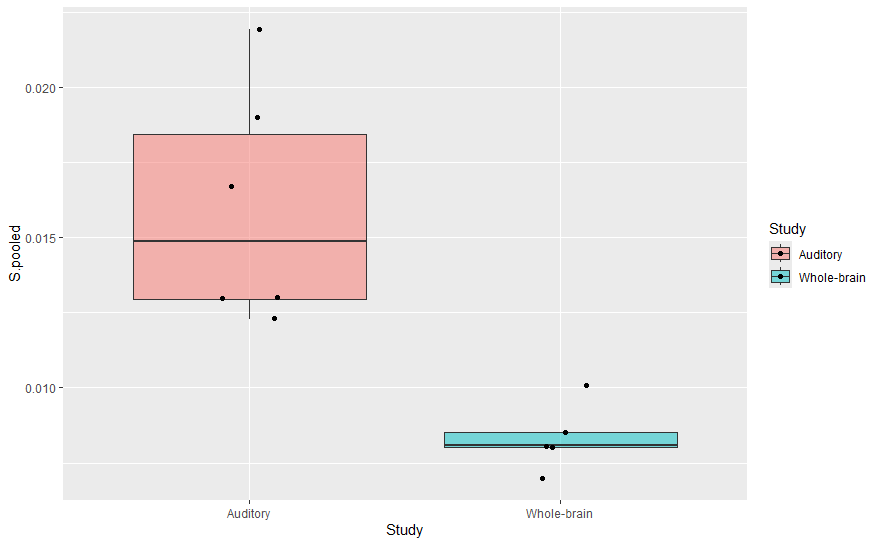

**Supplementary figure 9:** A plot showing **FA** variation in this study (auditory) compared to that of Magondo et al. (2024) (whole-brain).

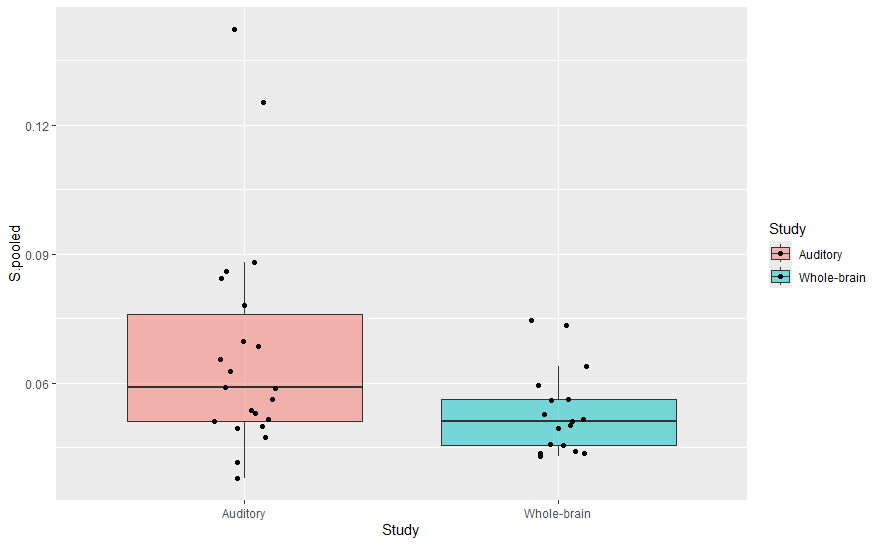

**Supplementary figure 10:** A plot showing **MD** variation in this study (auditory) compared to that of Magondo et al. (2024) (whole-brain).

**Supplementary table 3:** Summary of linear regression analysis of associations between FA and maternal CD4 counts, showing standardised beta (Std β) and standardised standard error (Std SE), p values and p values adjusted for multiple comparisons (q values) and the r and r^2^ values for these relationships.

| **Structures** | | **Std β** | **Std SE** | **p value** | **q value** | **Lower CI** | **Upper CI** | **r** | **r^2^** |
| --- | --- | --- | --- | --- | --- | --- | --- | --- | --- |
| L CC | R Medial geniculate nucleus | 0.107 | 0.132 | 0.420 | 0.898 | -0.157 | 0.371 | 0.107 | 0.011 |
| L Thal | L Heschl's gyrus | -0.100 | 0.132 | 0.449 | 0.898 | -0.364 | 0.164 | -0.100 | 0.010 |
| 4^th^ Vent | L Cochlear nucleus | -0.110 | 0.132 | 0.405 | 0.898 | -0.374 | 0.153 | -0.110 | 0.012 |
| 4^th^ Vent | R Cochlear nucleus | -0.010 | 0.141 | 0.945 | 0.945 | -0.294 | 0.274 | -0.010 | 0.000 |
| R CC | L Medial geniculate nucleus | -0.038 | 0.132 | 0.777 | 0.932 | -0.303 | 0.227 | -0.038 | 0.001 |
| Midbrain | R Heschl's gyrus | 0.070 | 0.132 | 0.599 | 0.898 | -0.195 | 0.335 | 0.070 | 0.005 |

**Supplementary table 4:** Summary of linear regression analysis of associations between MD and maternal CD4 counts, showing standardised beta (Std β) and standardised standard error (Std SE), p values and p values adjusted for multiple comparisons (q values) and the r and r^2^ values for these relationships.

| **Structures** | | **Std β** | **Std SE** | **p value** | **q value** | **Lower CI** | **Upper CI** | **r** | **r^2^** |
| --- | --- | --- | --- | --- | --- | --- | --- | --- | --- |
| L CC | L Inferior colliculus | -0.231 | 0.129 | 0.079 | 0.974 | -0.489 | 0.027 | -0.231 | 0.053 |
| L CC | L Heschl's gyrus | -0.033 | 0.132 | 0.804 | 0.974 | -0.298 | 0.232 | -0.033 | 0.001 |
| L CC | L Medial geniculate nucleus | -0.136 | 0.131 | 0.303 | 0.974 | -0.399 | 0.126 | -0.136 | 0.019 |
| L CC | R Medial geniculate nucleus | -0.096 | 0.132 | 0.467 | 0.974 | -0.360 | 0.168 | -0.096 | 0.009 |
| L Lat Vent | L Heschl's gyrus | -0.075 | 0.132 | 0.570 | 0.974 | -0.340 | 0.189 | -0.075 | 0.006 |
| L Lat Vent | L Medial geniculate nucleus | 0.004 | 0.132 | 0.974 | 0.974 | -0.261 | 0.270 | 0.004 | 0.000 |
| L Lat Vent | R Medial geniculate nucleus | -0.012 | 0.132 | 0.929 | 0.974 | -0.277 | 0.253 | -0.012 | 0.000 |
| L Thal | L Heschl's gyrus | -0.159 | 0.132 | 0.235 | 0.974 | -0.423 | 0.106 | -0.159 | 0.025 |
| L Thal | L Cochlear nucleus | 0.043 | 0.132 | 0.745 | 0.974 | -0.222 | 0.308 | 0.043 | 0.002 |
| L Thal | L Medial geniculate nucleus | 0.007 | 0.132 | 0.958 | 0.974 | -0.258 | 0.272 | 0.007 | 0.000 |
| L Caud | L Cochlear nucleus | 0.088 | 0.132 | 0.509 | 0.974 | -0.177 | 0.352 | 0.088 | 0.008 |
| L Caud | L Medial geniculate nucleus | -0.065 | 0.132 | 0.625 | 0.974 | -0.330 | 0.200 | -0.065 | 0.004 |
| 4^th^ Vent | R Cochlear nucleus | 0.197 | 0.130 | 0.136 | 0.974 | -0.063 | 0.457 | 0.197 | 0.039 |
| L VDC | L Medial geniculate nucleus | -0.072 | 0.132 | 0.589 | 0.974 | -0.336 | 0.193 | -0.072 | 0.005 |
| R CC | L Medial geniculate nucleus | -0.103 | 0.132 | 0.436 | 0.974 | -0.367 | 0.160 | -0.103 | 0.011 |
| R Lat Vent | R Medial geniculate nucleus | -0.037 | 0.132 | 0.782 | 0.974 | -0.302 | 0.228 | -0.037 | 0.001 |
| R Thal | L Medial geniculate nucleus | -0.222 | 0.129 | 0.091 | 0.974 | -0.481 | 0.037 | -0.222 | 0.049 |
| R Caud | R Medial geniculate nucleus | -0.029 | 0.132 | 0.829 | 0.974 | -0.294 | 0.236 | -0.029 | 0.001 |
| Midbrain | L Medial geniculate nucleus | -0.051 | 0.132 | 0.700 | 0.974 | -0.316 | 0.214 | -0.051 | 0.003 |
| Medulla | L Cochlear nucleus | -0.026 | 0.132 | 0.848 | 0.974 | -0.291 | 0.240 | -0.026 | 0.001 |
| Medulla | L Medial geniculate nucleus | -0.048 | 0.132 | 0.720 | 0.974 | -0.313 | 0.217 | -0.048 | 0.002 |
| Medulla | R Inferior colliculus | 0.072 | 0.132 | 0.586 | 0.974 | -0.192 | 0.337 | 0.072 | 0.005 |

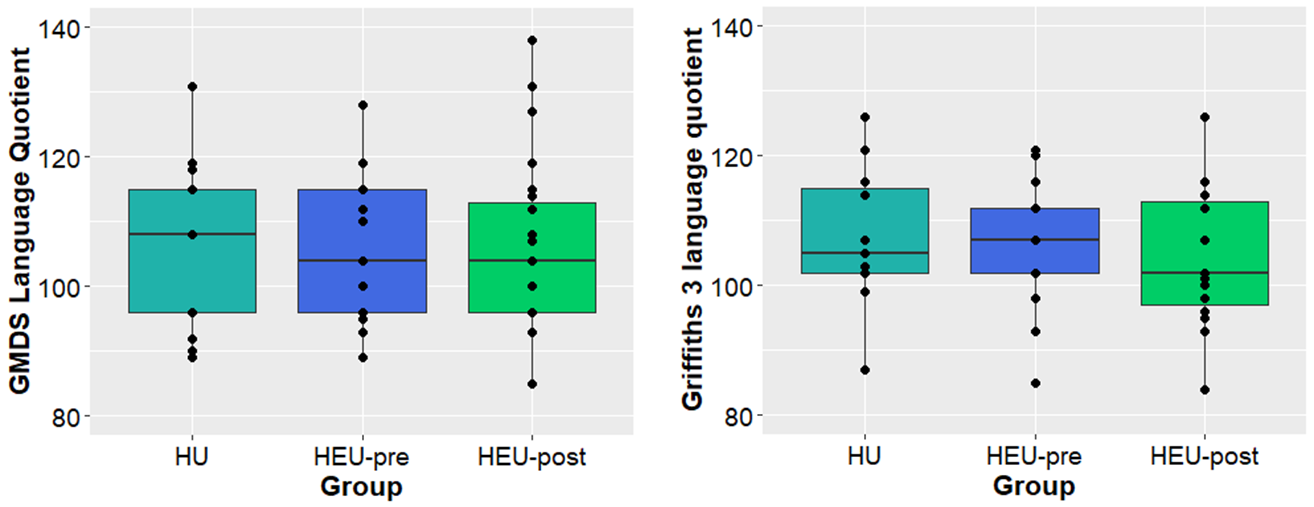

(b)

(a)

**Supplementary figure 11:** A box and whisker plot summarizing the upper, middle and lower quartiles of (a) GMDS language quotient and (b) Griffiths III language and communication quotient scores according to HU, HEU-pre and HEU-post infants; whiskers include data within 1.5 times the interquartile range above and below the upper and lower quartiles.

**Supplementary table 5:** Summary of linear regression analysis of associations between FA and GMDS language scores at 9-14 months across all infants, showing standardised beta (Std β), standardised standard error (Std SE), p values, p values adjusted for multiple comparisons (q values), 95% confidence interval (CI) for standardised beta coefficients and the r and r^2^ values for these relationships.

| **Structures** | | **Std β FA** | **Std SE FA** | **p FA** | **q FA** | **Lower CI** | **Upper CI** | **r** | **r^2^** |
| --- | --- | --- | --- | --- | --- | --- | --- | --- | --- |
| R Pal | R Medial geniculate nucleus | 0.314 | 0.130 | **0.020** | 0.995 | 0.052 | 0.576 | 0.314 | 0.099 |
| Vermis | L Cochlear nucleus | -0.237 | 0.133 | 0.081 | 0.995 | -0.505 | 0.031 | -0.237 | 0.056 |
| Pons | R Cochlear nucleus | 0.232 | 0.134 | 0.088 | 0.995 | -0.036 | 0.500 | 0.232 | 0.054 |
| L Caud | L Cochlear nucleus | 0.201 | 0.135 | 0.140 | 0.995 | -0.068 | 0.471 | 0.201 | 0.041 |
| R VDC | R Inferior colliculus | 0.184 | 0.135 | 0.179 | 0.995 | -0.087 | 0.455 | 0.184 | 0.034 |
| R Cb | R Cochlear nucleus | -0.168 | 0.135 | 0.221 | 0.995 | -0.439 | 0.104 | -0.168 | 0.028 |
| R Thal | R Heschl's gyrus | 0.164 | 0.135 | 0.230 | 0.995 | -0.107 | 0.436 | 0.164 | 0.027 |
| L Put | L Heschl's gyrus | 0.163 | 0.136 | 0.233 | 0.995 | -0.108 | 0.435 | 0.163 | 0.027 |
| L CC | L Heschl's gyrus | 0.159 | 0.136 | 0.247 | 0.995 | -0.113 | 0.431 | 0.159 | 0.025 |
| R Caud | R Heschl's gyrus | 0.156 | 0.136 | 0.257 | 0.995 | -0.117 | 0.428 | 0.156 | 0.024 |
| R VDC | L Cochlear nucleus | 0.150 | 0.136 | 0.274 | 0.995 | -0.122 | 0.422 | 0.150 | 0.023 |
| R CC | R Inferior colliculus | 0.148 | 0.136 | 0.280 | 0.995 | -0.124 | 0.421 | 0.148 | 0.022 |
| L Pal | L Cochlear nucleus | 0.147 | 0.136 | 0.284 | 0.995 | -0.125 | 0.420 | 0.147 | 0.022 |
| L CC | L Medial geniculate nucleus | 0.142 | 0.136 | 0.299 | 0.995 | -0.130 | 0.415 | 0.142 | 0.020 |
| R Heschl's gyrus | R Medial geniculate nucleus | 0.141 | 0.136 | 0.306 | 0.995 | -0.132 | 0.413 | 0.141 | 0.020 |
| Midbrain | R Cochlear nucleus | 0.136 | 0.136 | 0.321 | 0.995 | -0.137 | 0.409 | 0.136 | 0.019 |
| R Thal | R Inferior colliculus | 0.134 | 0.136 | 0.329 | 0.995 | -0.139 | 0.407 | 0.134 | 0.018 |
| L CC | L Cochlear nucleus | -0.128 | 0.136 | 0.352 | 0.995 | -0.401 | 0.145 | -0.128 | 0.016 |
| R VDC | R Cochlear nucleus | 0.124 | 0.136 | 0.368 | 0.995 | -0.150 | 0.397 | 0.124 | 0.015 |
| L Thal | R Medial geniculate nucleus | 0.121 | 0.136 | 0.377 | 0.995 | -0.152 | 0.395 | 0.121 | 0.015 |
| L Heschl's gyrus | L Medial geniculate nucleus | -0.121 | 0.136 | 0.380 | 0.995 | -0.394 | 0.153 | -0.121 | 0.015 |
| L VDC | R Inferior colliculus | 0.120 | 0.136 | 0.383 | 0.995 | -0.154 | 0.393 | 0.120 | 0.014 |
| Pons | R Heschl's gyrus | 0.115 | 0.136 | 0.404 | 0.995 | -0.159 | 0.388 | 0.115 | 0.013 |
| L Put | R Medial geniculate nucleus | 0.113 | 0.136 | 0.413 | 0.995 | -0.161 | 0.386 | 0.113 | 0.013 |
| R VDC | R Heschl's gyrus | 0.111 | 0.137 | 0.418 | 0.995 | -0.162 | 0.385 | 0.111 | 0.012 |
| Midbrain | R Medial geniculate nucleus | 0.110 | 0.137 | 0.424 | 0.995 | -0.164 | 0.384 | 0.110 | 0.012 |
| R Thal | R Cochlear nucleus | 0.109 | 0.137 | 0.428 | 0.995 | -0.165 | 0.383 | 0.109 | 0.012 |
| R Put | R Inferior colliculus | 0.108 | 0.137 | 0.434 | 0.995 | -0.166 | 0.382 | 0.108 | 0.012 |
| R VDC | L Inferior colliculus | 0.108 | 0.137 | 0.434 | 0.995 | -0.166 | 0.381 | 0.108 | 0.012 |
| Pons | R Medial geniculate nucleus | 0.108 | 0.137 | 0.434 | 0.995 | -0.166 | 0.381 | 0.108 | 0.012 |
| R Put | R Heschl's gyrus | -0.105 | 0.137 | 0.444 | 0.995 | -0.379 | 0.169 | -0.105 | 0.011 |
| 3rd V | L Inferior colliculus | 0.105 | 0.137 | 0.445 | 0.995 | -0.169 | 0.379 | 0.105 | 0.011 |
| R Lat V | R Medial geniculate nucleus | -0.105 | 0.137 | 0.446 | 0.995 | -0.379 | 0.169 | -0.105 | 0.011 |
| L CC | R Medial geniculate nucleus | -0.103 | 0.137 | 0.455 | 0.995 | -0.377 | 0.171 | -0.103 | 0.011 |
| Medulla | R Medial geniculate nucleus | 0.099 | 0.137 | 0.471 | 0.995 | -0.175 | 0.373 | 0.099 | 0.010 |
| 4th V | R Cochlear nucleus | -0.098 | 0.137 | 0.476 | 0.995 | -0.372 | 0.176 | -0.098 | 0.010 |
| Pons | L Cochlear nucleus | 0.098 | 0.137 | 0.476 | 0.995 | -0.176 | 0.372 | 0.098 | 0.010 |
| Medulla | L Medial geniculate nucleus | 0.097 | 0.137 | 0.482 | 0.995 | -0.177 | 0.371 | 0.097 | 0.009 |
| R Caud | R Medial geniculate nucleus | 0.095 | 0.137 | 0.492 | 0.995 | -0.180 | 0.369 | 0.095 | 0.009 |
| Midbrain | L Cochlear nucleus | 0.092 | 0.137 | 0.504 | 0.995 | -0.182 | 0.366 | 0.092 | 0.008 |
| L Hipp | L Heschl's gyrus | 0.088 | 0.137 | 0.523 | 0.995 | -0.187 | 0.362 | 0.088 | 0.008 |
| Vermis | L Medial geniculate nucleus | -0.086 | 0.137 | 0.534 | 0.995 | -0.360 | 0.189 | -0.086 | 0.007 |
| 4th V | R Inferior colliculus | -0.082 | 0.137 | 0.550 | 0.995 | -0.357 | 0.192 | -0.082 | 0.007 |
| L Inferior colliculus | L Cochlear nucleus | -0.081 | 0.137 | 0.557 | 0.995 | -0.356 | 0.194 | -0.081 | 0.007 |
| L VDC | L Inferior colliculus | 0.081 | 0.137 | 0.558 | 0.995 | -0.194 | 0.355 | 0.081 | 0.007 |
| R Cb | L Medial geniculate nucleus | 0.080 | 0.137 | 0.560 | 0.995 | -0.194 | 0.355 | 0.080 | 0.006 |
| L Inferior colliculus | R Inferior colliculus | -0.077 | 0.137 | 0.577 | 0.995 | -0.352 | 0.198 | -0.077 | 0.006 |
| L Medial geniculate nucleus | R Medial geniculate nucleus | -0.075 | 0.137 | 0.586 | 0.995 | -0.350 | 0.200 | -0.075 | 0.006 |
| R CC | L Inferior colliculus | 0.073 | 0.137 | 0.595 | 0.995 | -0.202 | 0.348 | 0.073 | 0.005 |
| L Thal | L Medial geniculate nucleus | 0.069 | 0.137 | 0.615 | 0.995 | -0.205 | 0.344 | 0.069 | 0.005 |
| 4th V | L Inferior colliculus | -0.069 | 0.137 | 0.618 | 0.995 | -0.344 | 0.206 | -0.069 | 0.005 |
| Medulla | L Cochlear nucleus | 0.066 | 0.137 | 0.630 | 0.995 | -0.209 | 0.341 | 0.066 | 0.004 |
| L Cb | L Cochlear nucleus | 0.063 | 0.137 | 0.649 | 0.995 | -0.212 | 0.338 | 0.063 | 0.004 |
| R VDC | R Medial geniculate nucleus | 0.062 | 0.137 | 0.651 | 0.995 | -0.213 | 0.337 | 0.062 | 0.004 |
| L Caud | L Heschl's gyrus | -0.061 | 0.137 | 0.661 | 0.995 | -0.336 | 0.215 | -0.061 | 0.004 |
| L Lat V | L Heschl's gyrus | 0.060 | 0.137 | 0.666 | 0.995 | -0.216 | 0.335 | 0.060 | 0.004 |
| R CC | R Medial geniculate nucleus | 0.059 | 0.137 | 0.666 | 0.995 | -0.216 | 0.335 | 0.059 | 0.004 |
| 4th V | L Cochlear nucleus | -0.058 | 0.137 | 0.674 | 0.995 | -0.333 | 0.217 | -0.058 | 0.003 |
| Midbrain | L Inferior colliculus | 0.058 | 0.137 | 0.675 | 0.995 | -0.217 | 0.333 | 0.058 | 0.003 |
| L Cb | R Medial geniculate nucleus | 0.057 | 0.137 | 0.681 | 0.995 | -0.218 | 0.332 | 0.057 | 0.003 |
| R Hipp | R Medial geniculate nucleus | 0.056 | 0.137 | 0.683 | 0.995 | -0.219 | 0.331 | 0.056 | 0.003 |
| L Thal | R Inferior colliculus | 0.055 | 0.137 | 0.692 | 0.995 | -0.221 | 0.330 | 0.055 | 0.003 |
| L Thal | L Heschl's gyrus | 0.054 | 0.137 | 0.693 | 0.995 | -0.221 | 0.330 | 0.054 | 0.003 |
| Medulla | R Cochlear nucleus | 0.053 | 0.137 | 0.699 | 0.995 | -0.222 | 0.328 | 0.053 | 0.003 |
| R Cb | L Inferior colliculus | 0.052 | 0.137 | 0.709 | 0.995 | -0.224 | 0.327 | 0.052 | 0.003 |
| Pons | R Inferior colliculus | 0.051 | 0.137 | 0.712 | 0.995 | -0.224 | 0.326 | 0.051 | 0.003 |
| Midbrain | R Inferior colliculus | 0.048 | 0.137 | 0.725 | 0.995 | -0.227 | 0.324 | 0.048 | 0.002 |
| Vermis | R Inferior colliculus | -0.047 | 0.137 | 0.733 | 0.995 | -0.322 | 0.228 | -0.047 | 0.002 |
| R Cb | R Medial geniculate nucleus | -0.044 | 0.137 | 0.748 | 0.995 | -0.320 | 0.231 | -0.044 | 0.002 |
| R Thal | L Medial geniculate nucleus | 0.042 | 0.137 | 0.762 | 0.995 | -0.233 | 0.317 | 0.042 | 0.002 |
| R Inferior colliculus | R Medial geniculate nucleus | 0.041 | 0.137 | 0.767 | 0.995 | -0.234 | 0.316 | 0.041 | 0.002 |
| R Lat V | R Heschl's gyrus | 0.041 | 0.137 | 0.768 | 0.995 | -0.234 | 0.316 | 0.041 | 0.002 |
| L Caud | L Medial geniculate nucleus | 0.040 | 0.137 | 0.772 | 0.995 | -0.235 | 0.315 | 0.040 | 0.002 |
| R CC | R Heschl's gyrus | 0.039 | 0.137 | 0.780 | 0.995 | -0.237 | 0.314 | 0.039 | 0.001 |
| Medulla | R Inferior colliculus | -0.038 | 0.137 | 0.783 | 0.995 | -0.313 | 0.237 | -0.038 | 0.001 |
| L VDC | L Medial geniculate nucleus | 0.038 | 0.137 | 0.786 | 0.995 | -0.238 | 0.313 | 0.038 | 0.001 |
| Pons | L Inferior colliculus | 0.034 | 0.137 | 0.805 | 0.995 | -0.241 | 0.309 | 0.034 | 0.001 |
| L VDC | L Cochlear nucleus | 0.029 | 0.137 | 0.831 | 0.995 | -0.246 | 0.305 | 0.029 | 0.001 |
| R CC | L Medial geniculate nucleus | -0.028 | 0.137 | 0.838 | 0.995 | -0.304 | 0.247 | -0.028 | 0.001 |
| L Cb | R Inferior colliculus | -0.027 | 0.137 | 0.844 | 0.995 | -0.303 | 0.248 | -0.027 | 0.001 |
| R Amygdala | R Medial geniculate nucleus | 0.026 | 0.137 | 0.851 | 0.995 | -0.250 | 0.301 | 0.026 | 0.001 |
| R Put | R Medial geniculate nucleus | -0.025 | 0.137 | 0.859 | 0.995 | -0.300 | 0.251 | -0.025 | 0.001 |
| L Cb | L Inferior colliculus | 0.020 | 0.137 | 0.883 | 0.995 | -0.255 | 0.296 | 0.020 | 0.000 |
| L Lat V | L Inferior colliculus | -0.020 | 0.137 | 0.885 | 0.995 | -0.295 | 0.256 | -0.020 | 0.000 |
| R Thal | L Inferior colliculus | 0.019 | 0.137 | 0.891 | 0.995 | -0.256 | 0.294 | 0.019 | 0.000 |
| L Thal | L Inferior colliculus | -0.019 | 0.137 | 0.891 | 0.995 | -0.294 | 0.257 | -0.019 | 0.000 |
| L Lat V | R Medial geniculate nucleus | 0.018 | 0.137 | 0.896 | 0.995 | -0.257 | 0.293 | 0.018 | 0.000 |
| R Inferior colliculus | R Cochlear nucleus | 0.017 | 0.137 | 0.905 | 0.995 | -0.259 | 0.292 | 0.017 | 0.000 |
| R Cb | L Cochlear nucleus | -0.016 | 0.137 | 0.906 | 0.995 | -0.292 | 0.259 | -0.016 | 0.000 |
| Midbrain | L Medial geniculate nucleus | 0.016 | 0.137 | 0.910 | 0.995 | -0.260 | 0.291 | 0.016 | 0.000 |
| L Hipp | L Medial geniculate nucleus | 0.014 | 0.137 | 0.917 | 0.995 | -0.261 | 0.290 | 0.014 | 0.000 |
| L Thal | L Cochlear nucleus | -0.014 | 0.137 | 0.920 | 0.995 | -0.289 | 0.262 | -0.014 | 0.000 |
| Midbrain | R Heschl's gyrus | 0.014 | 0.137 | 0.921 | 0.995 | -0.262 | 0.289 | 0.014 | 0.000 |
| L Inferior colliculus | L Medial geniculate nucleus | -0.013 | 0.137 | 0.924 | 0.995 | -0.289 | 0.262 | -0.013 | 0.000 |
| Vermis | R Medial geniculate nucleus | 0.012 | 0.137 | 0.930 | 0.995 | -0.263 | 0.288 | 0.012 | 0.000 |
| Vermis | L Inferior colliculus | 0.012 | 0.137 | 0.933 | 0.995 | -0.264 | 0.287 | 0.012 | 0.000 |
| L Cb | L Medial geniculate nucleus | -0.009 | 0.137 | 0.947 | 0.995 | -0.285 | 0.266 | -0.009 | 0.000 |
| Pons | L Medial geniculate nucleus | 0.009 | 0.137 | 0.947 | 0.995 | -0.266 | 0.285 | 0.009 | 0.000 |
| L CC | L Inferior colliculus | 0.007 | 0.137 | 0.961 | 0.995 | -0.269 | 0.282 | 0.007 | 0.000 |
| L Put | L Medial geniculate nucleus | 0.007 | 0.137 | 0.962 | 0.995 | -0.269 | 0.282 | 0.007 | 0.000 |
| R Cb | R Inferior colliculus | 0.005 | 0.137 | 0.969 | 0.995 | -0.270 | 0.281 | 0.005 | 0.000 |
| L Lat V | L Medial geniculate nucleus | 0.004 | 0.137 | 0.976 | 0.995 | -0.271 | 0.280 | 0.004 | 0.000 |
| L Cochlear nucleus | R Cochlear nucleus | -0.003 | 0.137 | 0.982 | 0.995 | -0.279 | 0.272 | -0.003 | 0.000 |
| R Thal | R Medial geniculate nucleus | 0.002 | 0.137 | 0.990 | 0.995 | -0.274 | 0.277 | 0.002 | 0.000 |
| R Caud | R Inferior colliculus | 0.002 | 0.137 | 0.991 | 0.995 | -0.274 | 0.277 | 0.002 | 0.000 |
| L Caud | L Inferior colliculus | -0.001 | 0.137 | 0.995 | 0.995 | -0.276 | 0.275 | -0.001 | 0.000 |

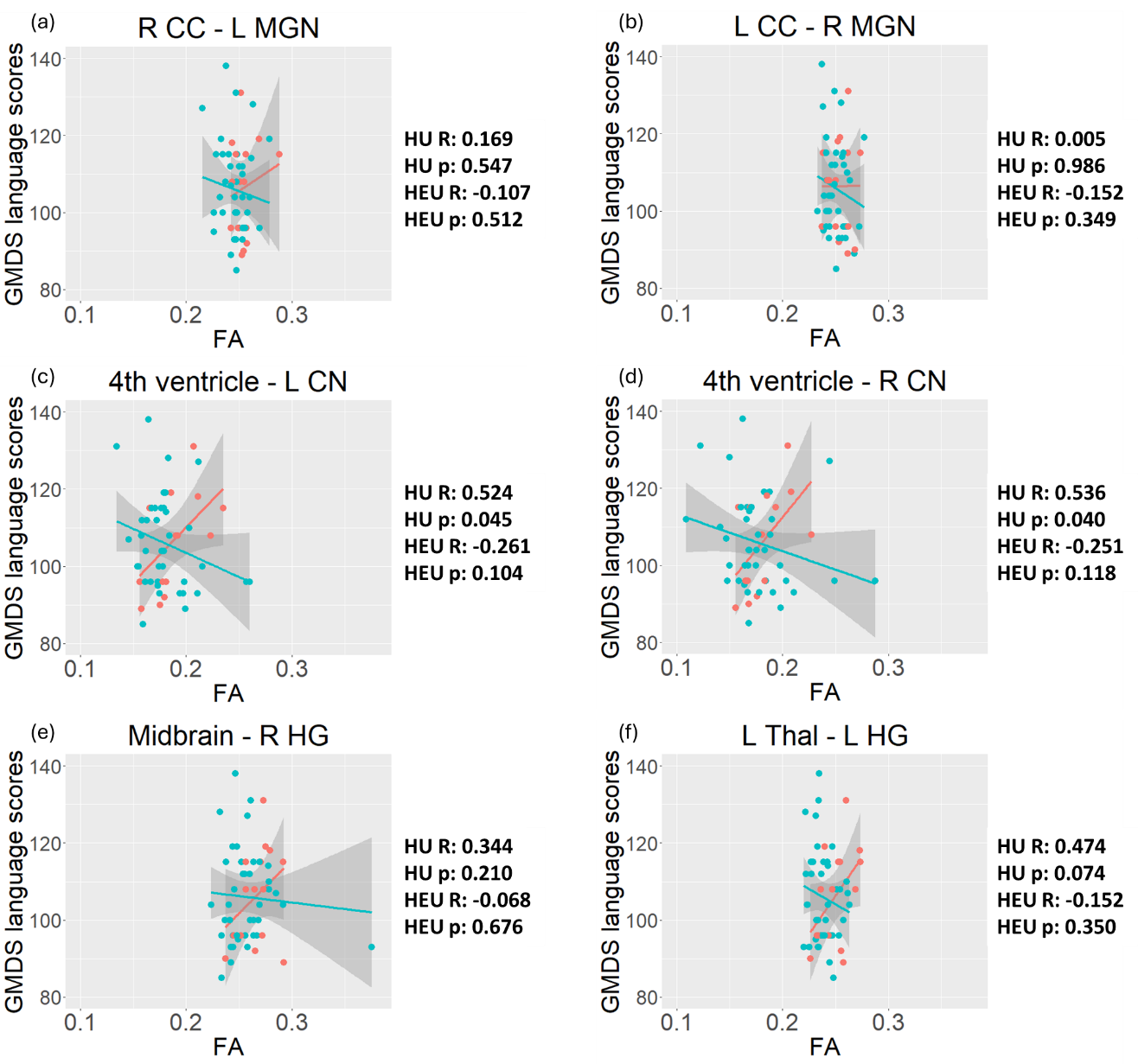
**Supplementary figure 12:** The relationship between fractional anisotropy (FA) and language outcomes according to HIV exposure groups, for tracts in which we formerly observed differences in FA between iHEU and iHU. Red=iHU; Blue =iHEU.

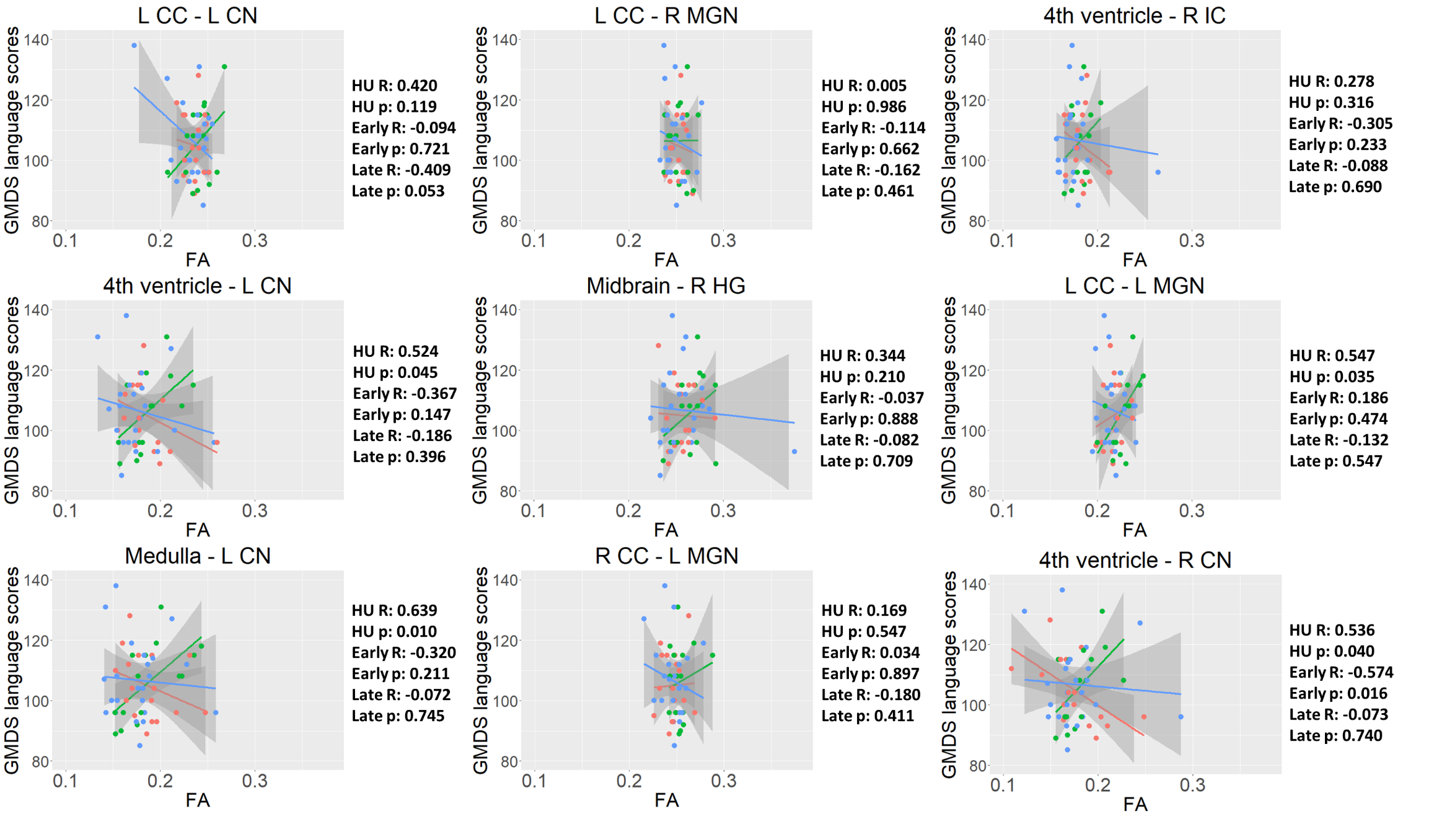
**Supplementary figure 13:** The relationship between fractional anisotropy (FA) and language outcomes according to ART exposure groups, for tracts in which we formerly observed differences in FA between iHEU-post and iHU. Green = iHU Red=iHEU-pre (early); Blue = iHEU-post (late).

**Supplementary table 6:** Summary of linear regression analysis of associations between MD and GMDS language scores at 9-14 months across all infants, showing standardised beta (Std β), standardised standard error (Std SE), p values, p values adjusted for multiple comparisons (q values), 95% confidence interval (CI) for standardised beta coefficients and the r and r^2^ values for these relationships.

| **Structures** | | **Std β MD** | **Std SE MD** | **p MD** | **q MD** | **Lower CI** | **Upper CI** | **r** | **r^2^** |
| --- | --- | --- | --- | --- | --- | --- | --- | --- | --- |
| R Caud | R Heschl's gyrus | -0.350 | 0.129 | **0.009** | 0.859 | -0.608 | -0.092 | -0.350 | 0.123 |
| L Cb | R Inferior colliculus | -0.320 | 0.130 | **0.017** | 0.859 | -0.581 | -0.059 | -0.320 | 0.102 |
| R VDC | R Cochlear nucleus | -0.284 | 0.132 | **0.036** | 0.859 | -0.548 | -0.019 | -0.284 | 0.080 |
| L VDC | L Inferior colliculus | 0.274 | 0.133 | **0.045** | 0.859 | 0.006 | 0.541 | 0.274 | 0.075 |
| Pons | L Medial geniculate nucleus | -0.257 | 0.134 | 0.060 | 0.859 | -0.526 | 0.011 | -0.257 | 0.066 |
| Pons | R Inferior colliculus | -0.253 | 0.133 | 0.062 | 0.859 | -0.520 | 0.013 | -0.253 | 0.064 |
| L Pal | L Cochlear nucleus | 0.251 | 0.133 | 0.065 | 0.859 | -0.016 | 0.518 | 0.251 | 0.063 |
| Medulla | L Cochlear nucleus | -0.247 | 0.133 | 0.069 | 0.859 | -0.514 | 0.020 | -0.247 | 0.061 |
| Pons | L Inferior colliculus | -0.244 | 0.133 | 0.073 | 0.859 | -0.511 | 0.023 | -0.244 | 0.059 |
| R Lat V | R Heschl's gyrus | -0.228 | 0.134 | 0.094 | 0.926 | -0.496 | 0.040 | -0.228 | 0.052 |
| Vermis | L Cochlear nucleus | 0.236 | 0.139 | 0.096 | 0.926 | -0.043 | 0.515 | 0.236 | 0.056 |
| L Cb | L Medial geniculate nucleus | -0.221 | 0.134 | 0.105 | 0.927 | -0.490 | 0.048 | -0.221 | 0.049 |
| R Cb | R Medial geniculate nucleus | -0.215 | 0.134 | 0.114 | 0.932 | -0.484 | 0.054 | -0.215 | 0.046 |
| R Hipp | R Medial geniculate nucleus | -0.193 | 0.135 | 0.157 | 0.975 | -0.464 | 0.077 | -0.193 | 0.037 |
| L Caud | L Cochlear nucleus | -0.192 | 0.135 | 0.159 | 0.975 | -0.463 | 0.078 | -0.192 | 0.037 |
| L Cb | L Inferior colliculus | -0.186 | 0.135 | 0.173 | 0.975 | -0.457 | 0.084 | -0.186 | 0.035 |
| L CC | L Cochlear nucleus | 0.185 | 0.135 | 0.177 | 0.975 | -0.086 | 0.455 | 0.185 | 0.034 |
| L Hipp | L Medial geniculate nucleus | -0.183 | 0.135 | 0.181 | 0.975 | -0.454 | 0.088 | -0.183 | 0.033 |
| R Cb | R Inferior colliculus | -0.166 | 0.135 | 0.225 | 0.975 | -0.438 | 0.105 | -0.166 | 0.028 |
| Vermis | R Inferior colliculus | -0.166 | 0.135 | 0.227 | 0.975 | -0.437 | 0.106 | -0.166 | 0.027 |
| Medulla | R Cochlear nucleus | -0.164 | 0.136 | 0.232 | 0.975 | -0.436 | 0.108 | -0.164 | 0.027 |
| R Thal | R Heschl's gyrus | -0.161 | 0.136 | 0.240 | 0.975 | -0.433 | 0.111 | -0.161 | 0.026 |
| Pons | R Medial geniculate nucleus | -0.160 | 0.136 | 0.244 | 0.975 | -0.432 | 0.112 | -0.160 | 0.025 |
| L Put | R Medial geniculate nucleus | 0.159 | 0.136 | 0.247 | 0.975 | -0.113 | 0.431 | 0.159 | 0.025 |
| Midbrain | L Medial geniculate nucleus | -0.158 | 0.136 | 0.250 | 0.975 | -0.430 | 0.114 | -0.158 | 0.025 |
| Vermis | L Medial geniculate nucleus | -0.154 | 0.136 | 0.261 | 0.975 | -0.427 | 0.118 | -0.154 | 0.024 |
| L Inferior colliculus | R Inferior colliculus | -0.153 | 0.136 | 0.264 | 0.975 | -0.425 | 0.119 | -0.153 | 0.023 |
| R Thal | L Medial geniculate nucleus | -0.150 | 0.136 | 0.276 | 0.975 | -0.422 | 0.123 | -0.150 | 0.022 |
| R Cb | L Medial geniculate nucleus | -0.137 | 0.136 | 0.318 | 0.975 | -0.410 | 0.136 | -0.137 | 0.019 |
| L VDC | L Cochlear nucleus | -0.136 | 0.136 | 0.324 | 0.975 | -0.409 | 0.137 | -0.136 | 0.018 |
| L Caud | L Medial geniculate nucleus | -0.133 | 0.136 | 0.335 | 0.975 | -0.406 | 0.141 | -0.133 | 0.018 |
| L Caud | L Inferior colliculus | -0.129 | 0.136 | 0.347 | 0.975 | -0.402 | 0.144 | -0.129 | 0.017 |
| Vermis | L Inferior colliculus | -0.128 | 0.136 | 0.352 | 0.975 | -0.401 | 0.145 | -0.128 | 0.016 |
| L Thal | L Medial geniculate nucleus | -0.126 | 0.136 | 0.359 | 0.975 | -0.400 | 0.147 | -0.126 | 0.016 |
| R Thal | L Inferior colliculus | 0.121 | 0.136 | 0.380 | 0.975 | -0.153 | 0.394 | 0.121 | 0.015 |
| L CC | L Medial geniculate nucleus | -0.119 | 0.136 | 0.385 | 0.975 | -0.393 | 0.154 | -0.119 | 0.014 |
| Pons | R Heschl's gyrus | -0.119 | 0.136 | 0.387 | 0.975 | -0.393 | 0.154 | -0.119 | 0.014 |
| L Thal | L Inferior colliculus | -0.118 | 0.136 | 0.391 | 0.975 | -0.391 | 0.156 | -0.118 | 0.014 |
| R Pal | R Medial geniculate nucleus | 0.113 | 0.136 | 0.412 | 0.975 | -0.161 | 0.387 | 0.113 | 0.013 |
| L Lat V | L Medial geniculate nucleus | -0.111 | 0.137 | 0.418 | 0.975 | -0.385 | 0.162 | -0.111 | 0.012 |
| Midbrain | L Inferior colliculus | -0.108 | 0.137 | 0.434 | 0.975 | -0.381 | 0.166 | -0.108 | 0.012 |
| L Cb | R Medial geniculate nucleus | -0.101 | 0.137 | 0.461 | 0.975 | -0.376 | 0.173 | -0.101 | 0.010 |
| R Inferior colliculus | R Cochlear nucleus | -0.099 | 0.137 | 0.471 | 0.975 | -0.373 | 0.175 | -0.099 | 0.010 |
| R Put | R Inferior colliculus | 0.099 | 0.137 | 0.472 | 0.975 | -0.175 | 0.373 | 0.099 | 0.010 |
| L Inferior colliculus | L Medial geniculate nucleus | -0.099 | 0.137 | 0.473 | 0.975 | -0.373 | 0.175 | -0.099 | 0.010 |
| Midbrain | L Cochlear nucleus | -0.097 | 0.137 | 0.482 | 0.975 | -0.371 | 0.177 | -0.097 | 0.009 |
| R Amygdala | R Medial geniculate nucleus | -0.095 | 0.137 | 0.491 | 0.975 | -0.369 | 0.179 | -0.095 | 0.009 |
| R Thal | R Cochlear nucleus | -0.090 | 0.137 | 0.512 | 0.975 | -0.365 | 0.184 | -0.090 | 0.008 |
| L CC | L Inferior colliculus | -0.090 | 0.137 | 0.512 | 0.975 | -0.365 | 0.184 | -0.090 | 0.008 |
| R Caud | R Medial geniculate nucleus | -0.089 | 0.137 | 0.518 | 0.975 | -0.363 | 0.185 | -0.089 | 0.008 |
| R CC | R Heschl's gyrus | -0.089 | 0.137 | 0.520 | 0.975 | -0.363 | 0.186 | -0.089 | 0.008 |
| R CC | L Inferior colliculus | -0.086 | 0.137 | 0.534 | 0.975 | -0.360 | 0.189 | -0.086 | 0.007 |
| R Cb | L Inferior colliculus | -0.085 | 0.137 | 0.538 | 0.975 | -0.359 | 0.190 | -0.085 | 0.007 |
| L Caud | L Heschl's gyrus | 0.083 | 0.137 | 0.549 | 0.975 | -0.192 | 0.357 | 0.083 | 0.007 |
| L Lat V | R Medial geniculate nucleus | -0.082 | 0.137 | 0.550 | 0.975 | -0.357 | 0.192 | -0.082 | 0.007 |
| Pons | L Cochlear nucleus | -0.082 | 0.137 | 0.554 | 0.975 | -0.356 | 0.193 | -0.082 | 0.007 |
| L Thal | L Heschl's gyrus | 0.079 | 0.137 | 0.565 | 0.975 | -0.195 | 0.354 | 0.079 | 0.006 |
| R CC | R Inferior colliculus | -0.078 | 0.137 | 0.569 | 0.975 | -0.353 | 0.196 | -0.078 | 0.006 |
| R VDC | L Inferior colliculus | 0.075 | 0.137 | 0.585 | 0.975 | -0.199 | 0.350 | 0.075 | 0.006 |
| R Cb | R Cochlear nucleus | -0.073 | 0.137 | 0.594 | 0.975 | -0.348 | 0.201 | -0.073 | 0.005 |
| R VDC | L Cochlear nucleus | -0.073 | 0.137 | 0.597 | 0.975 | -0.348 | 0.202 | -0.073 | 0.005 |
| Vermis | R Medial geniculate nucleus | -0.072 | 0.137 | 0.603 | 0.975 | -0.347 | 0.203 | -0.072 | 0.005 |
| R VDC | R Inferior colliculus | -0.069 | 0.137 | 0.615 | 0.975 | -0.344 | 0.206 | -0.069 | 0.005 |
| L Put | L Medial geniculate nucleus | 0.068 | 0.137 | 0.621 | 0.975 | -0.207 | 0.343 | 0.068 | 0.005 |
| R VDC | R Heschl's gyrus | 0.068 | 0.137 | 0.621 | 0.975 | -0.207 | 0.343 | 0.068 | 0.005 |
| L VDC | R Inferior colliculus | 0.068 | 0.137 | 0.624 | 0.975 | -0.207 | 0.343 | 0.068 | 0.005 |
| L Thal | R Medial geniculate nucleus | -0.067 | 0.137 | 0.629 | 0.975 | -0.341 | 0.208 | -0.067 | 0.004 |
| Midbrain | R Cochlear nucleus | -0.062 | 0.137 | 0.653 | 0.975 | -0.337 | 0.213 | -0.062 | 0.004 |
| L Heschl's gyrus | L Medial geniculate nucleus | 0.060 | 0.137 | 0.666 | 0.975 | -0.215 | 0.335 | 0.060 | 0.004 |
| L Medial geniculate nucleus | R Medial geniculate nucleus | 0.058 | 0.137 | 0.672 | 0.975 | -0.217 | 0.333 | 0.058 | 0.003 |
| Pons | R Cochlear nucleus | -0.056 | 0.137 | 0.684 | 0.975 | -0.331 | 0.219 | -0.056 | 0.003 |
| R Cb | L Cochlear nucleus | 0.054 | 0.137 | 0.694 | 0.975 | -0.221 | 0.329 | 0.054 | 0.003 |
| L Cb | L Cochlear nucleus | -0.054 | 0.137 | 0.697 | 0.975 | -0.329 | 0.221 | -0.054 | 0.003 |
| L VDC | L Medial geniculate nucleus | -0.050 | 0.137 | 0.715 | 0.975 | -0.326 | 0.225 | -0.050 | 0.003 |
| L Thal | R Inferior colliculus | -0.049 | 0.137 | 0.725 | 0.975 | -0.324 | 0.227 | -0.049 | 0.002 |
| L CC | R Medial geniculate nucleus | 0.048 | 0.137 | 0.730 | 0.975 | -0.228 | 0.323 | 0.048 | 0.002 |
| L Lat V | L Heschl's gyrus | 0.045 | 0.137 | 0.745 | 0.975 | -0.230 | 0.320 | 0.045 | 0.002 |
| R Inferior colliculus | R Medial geniculate nucleus | -0.042 | 0.137 | 0.763 | 0.975 | -0.317 | 0.234 | -0.042 | 0.002 |
| 4th V | R Inferior colliculus | -0.041 | 0.137 | 0.766 | 0.975 | -0.316 | 0.234 | -0.041 | 0.002 |
| Medulla | L Medial geniculate nucleus | -0.041 | 0.137 | 0.768 | 0.975 | -0.316 | 0.235 | -0.041 | 0.002 |
| R Caud | R Inferior colliculus | -0.040 | 0.137 | 0.770 | 0.975 | -0.316 | 0.235 | -0.040 | 0.002 |
| R CC | L Medial geniculate nucleus | 0.040 | 0.137 | 0.770 | 0.975 | -0.235 | 0.316 | 0.040 | 0.002 |
| Medulla | R Medial geniculate nucleus | -0.040 | 0.137 | 0.772 | 0.975 | -0.315 | 0.235 | -0.040 | 0.002 |
| R Put | R Medial geniculate nucleus | 0.040 | 0.137 | 0.774 | 0.975 | -0.236 | 0.315 | 0.040 | 0.002 |
| R Thal | R Inferior colliculus | -0.038 | 0.137 | 0.782 | 0.975 | -0.314 | 0.237 | -0.038 | 0.001 |
| Midbrain | R Heschl's gyrus | 0.034 | 0.137 | 0.807 | 0.981 | -0.242 | 0.309 | 0.034 | 0.001 |
| 4th V | L Inferior colliculus | 0.033 | 0.137 | 0.811 | 0.981 | -0.242 | 0.308 | 0.033 | 0.001 |
| L Put | L Heschl's gyrus | 0.032 | 0.137 | 0.814 | 0.981 | -0.243 | 0.308 | 0.032 | 0.001 |
| Midbrain | R Inferior colliculus | -0.027 | 0.137 | 0.845 | 0.989 | -0.302 | 0.248 | -0.027 | 0.001 |
| R VDC | R Medial geniculate nucleus | -0.025 | 0.137 | 0.855 | 0.989 | -0.301 | 0.250 | -0.025 | 0.001 |
| Midbrain | R Medial geniculate nucleus | 0.025 | 0.137 | 0.857 | 0.989 | -0.251 | 0.300 | 0.025 | 0.001 |
| 4th V | L Cochlear nucleus | 0.024 | 0.137 | 0.861 | 0.989 | -0.251 | 0.300 | 0.024 | 0.001 |
| 3rd V | L Inferior colliculus | 0.020 | 0.137 | 0.884 | 0.989 | -0.255 | 0.296 | 0.020 | 0.000 |
| L Thal | L Cochlear nucleus | 0.020 | 0.137 | 0.886 | 0.989 | -0.256 | 0.295 | 0.020 | 0.000 |
| L Lat V | L Inferior colliculus | -0.014 | 0.137 | 0.920 | 0.989 | -0.289 | 0.262 | -0.014 | 0.000 |
| L Inferior colliculus | L Cochlear nucleus | 0.014 | 0.137 | 0.921 | 0.989 | -0.262 | 0.289 | 0.014 | 0.000 |
| L Cochlear nucleus | R Cochlear nucleus | 0.010 | 0.137 | 0.940 | 0.989 | -0.265 | 0.286 | 0.010 | 0.000 |
| L Hipp | L Heschl's gyrus | 0.010 | 0.137 | 0.943 | 0.989 | -0.266 | 0.285 | 0.010 | 0.000 |
| Medulla | R Inferior colliculus | -0.009 | 0.137 | 0.949 | 0.989 | -0.284 | 0.267 | -0.009 | 0.000 |
| L CC | L Heschl's gyrus | 0.009 | 0.137 | 0.949 | 0.989 | -0.267 | 0.284 | 0.009 | 0.000 |
| R Thal | R Medial geniculate nucleus | -0.009 | 0.137 | 0.949 | 0.989 | -0.284 | 0.267 | -0.009 | 0.000 |
| R Lat V | R Medial geniculate nucleus | -0.005 | 0.137 | 0.968 | 0.989 | -0.281 | 0.270 | -0.005 | 0.000 |
| R Put | R Heschl's gyrus | 0.005 | 0.137 | 0.970 | 0.989 | -0.270 | 0.281 | 0.005 | 0.000 |
| R CC | R Medial geniculate nucleus | 0.003 | 0.137 | 0.983 | 0.989 | -0.273 | 0.278 | 0.003 | 0.000 |
| 4th V | R Cochlear nucleus | 0.002 | 0.137 | 0.986 | 0.989 | -0.273 | 0.278 | 0.002 | 0.000 |
| R Heschl's gyrus | R Medial geniculate nucleus | -0.002 | 0.137 | 0.989 | 0.989 | -0.277 | 0.274 | -0.002 | 0.000 |

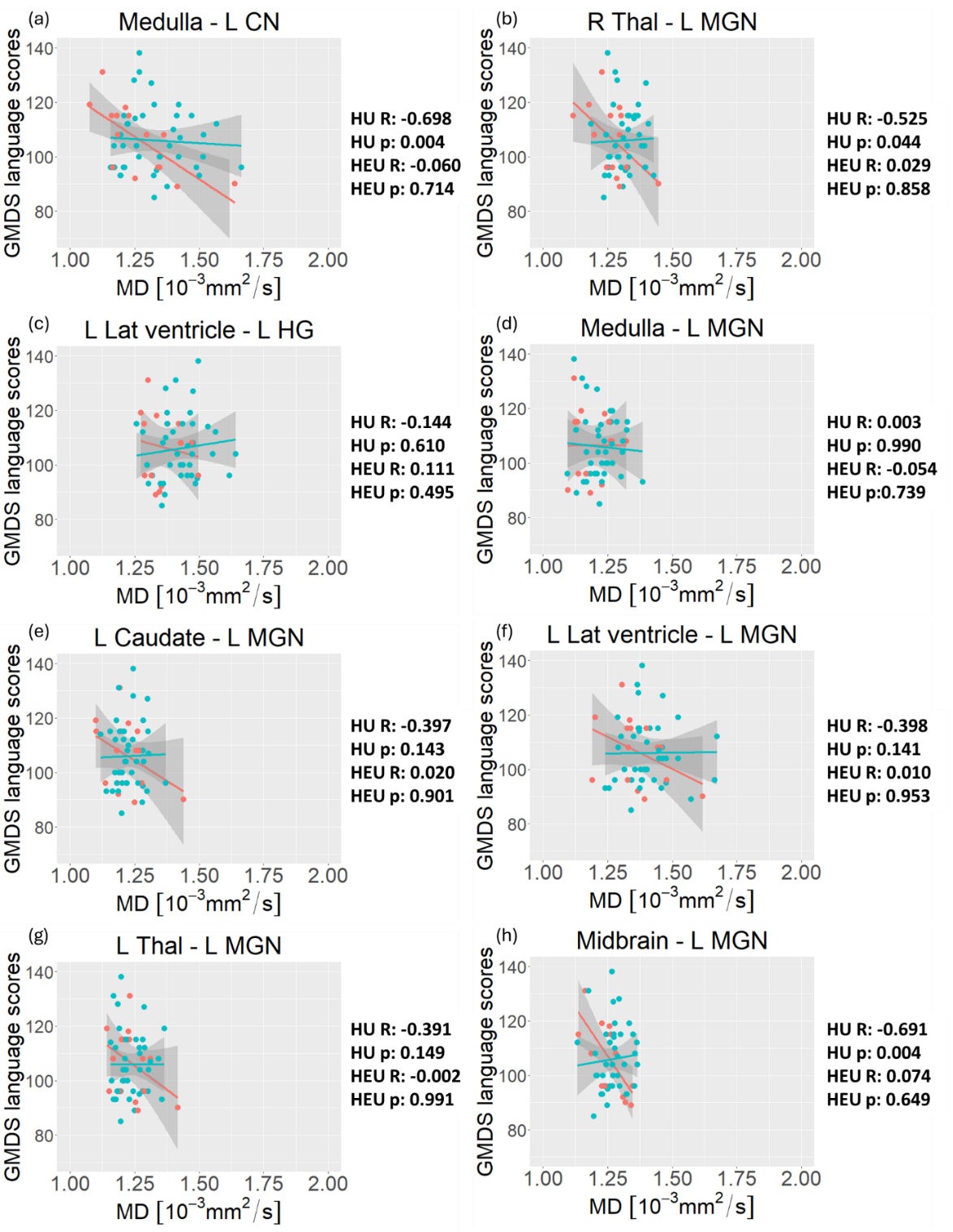

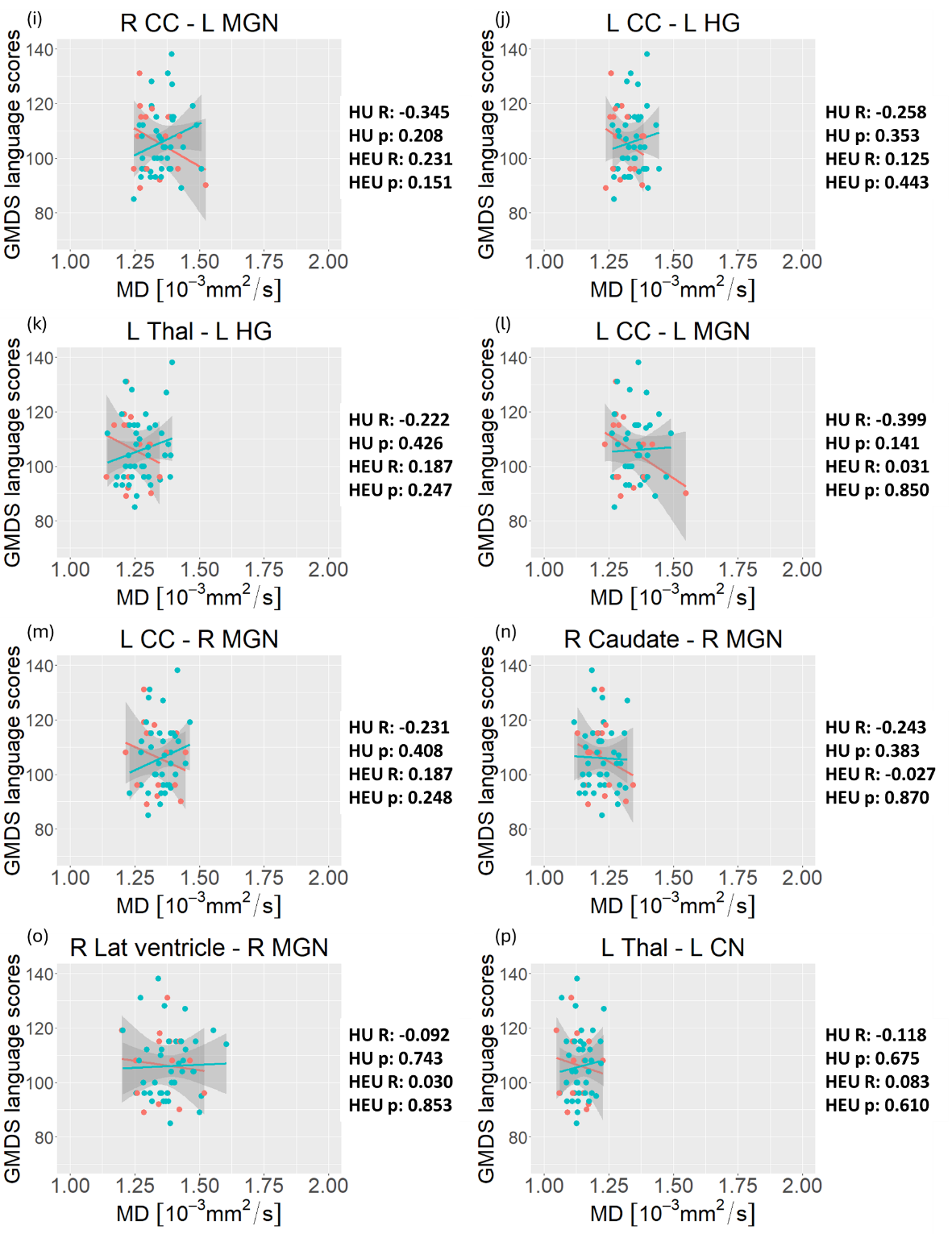

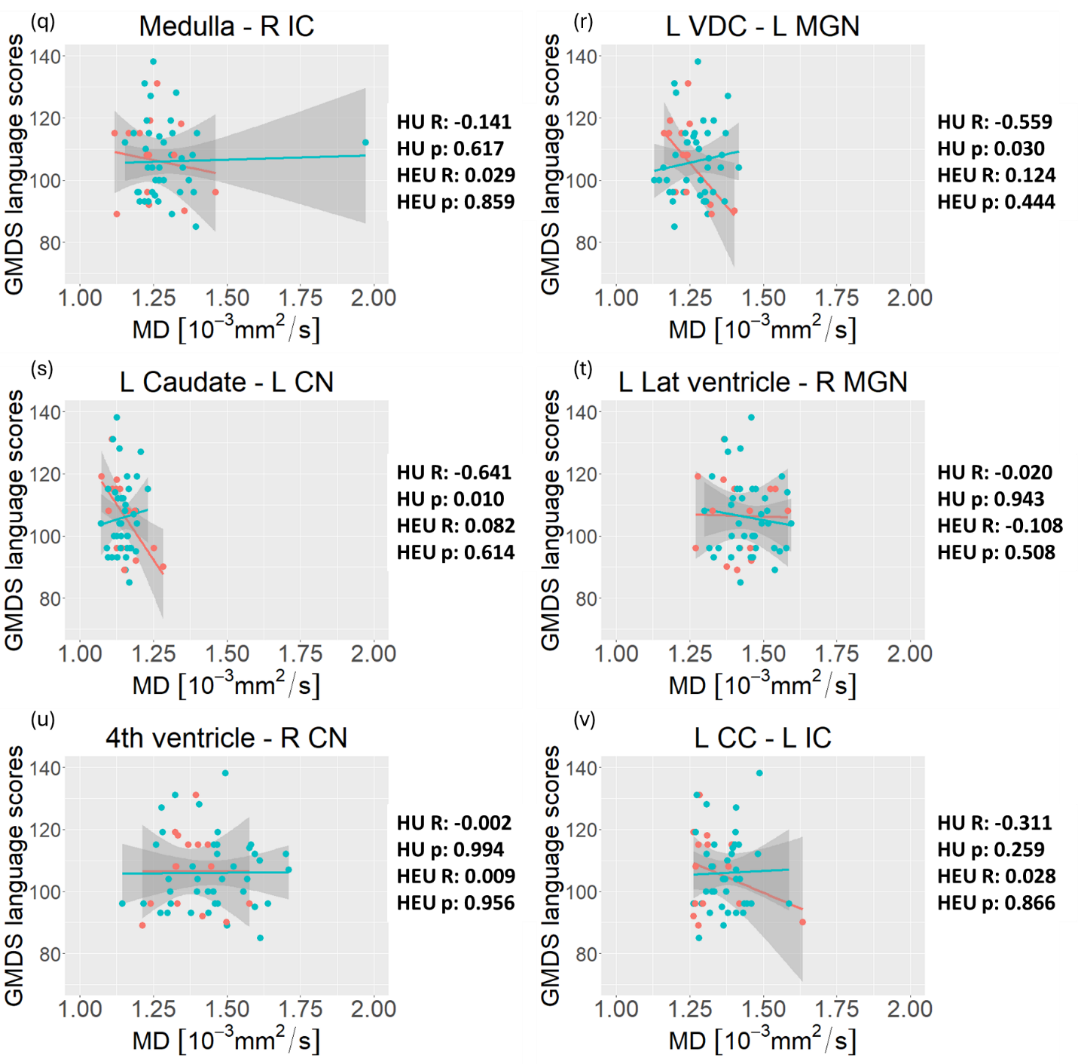

**Supplementary figure 14:** The relationship between mean diffusivity (MD) and language outcomes according to HIV exposure groups, for tracts in which we formerly observed differences in MD between iHEU and iHU. Red=iHU; Blue =iHEU.

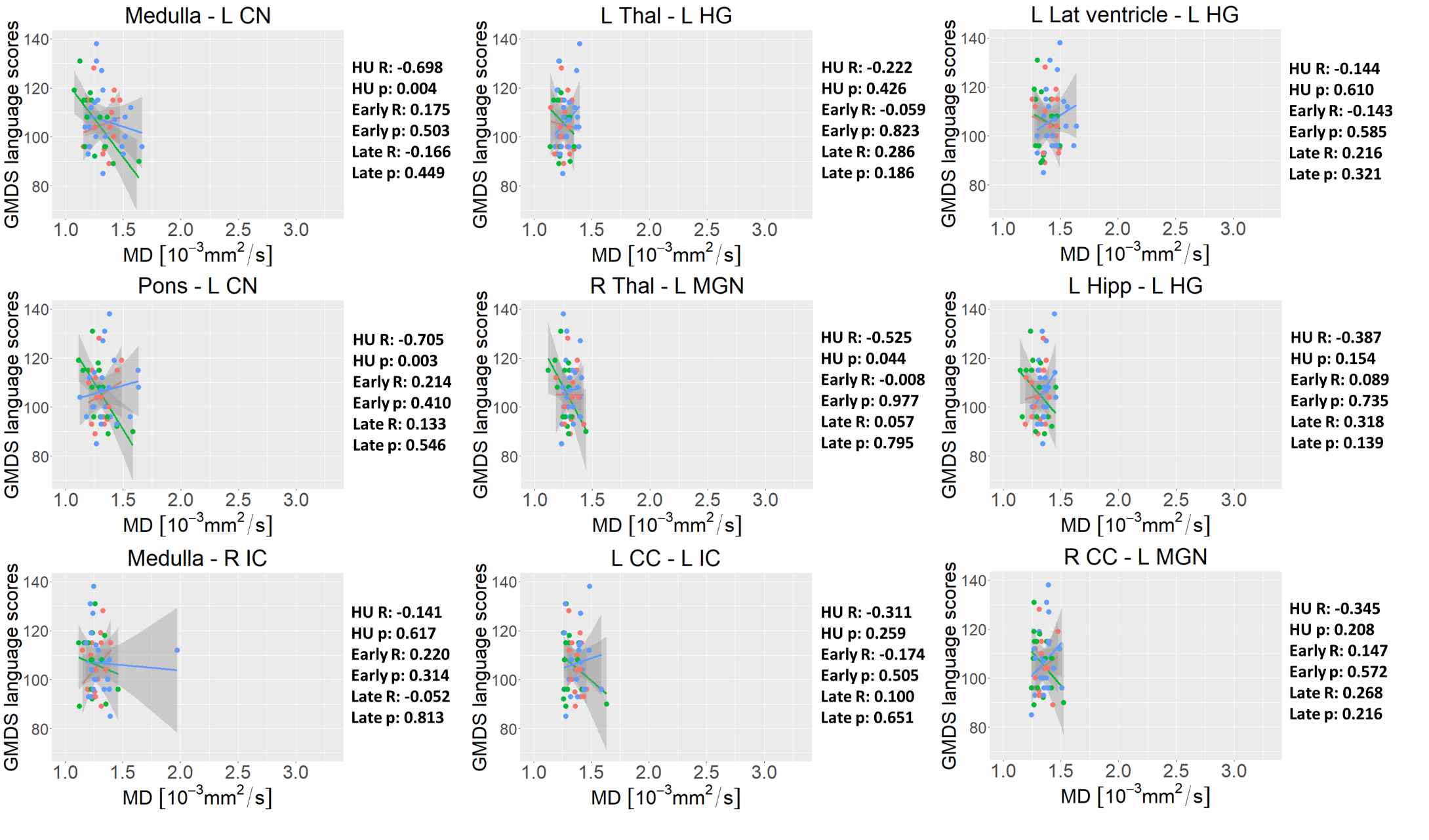

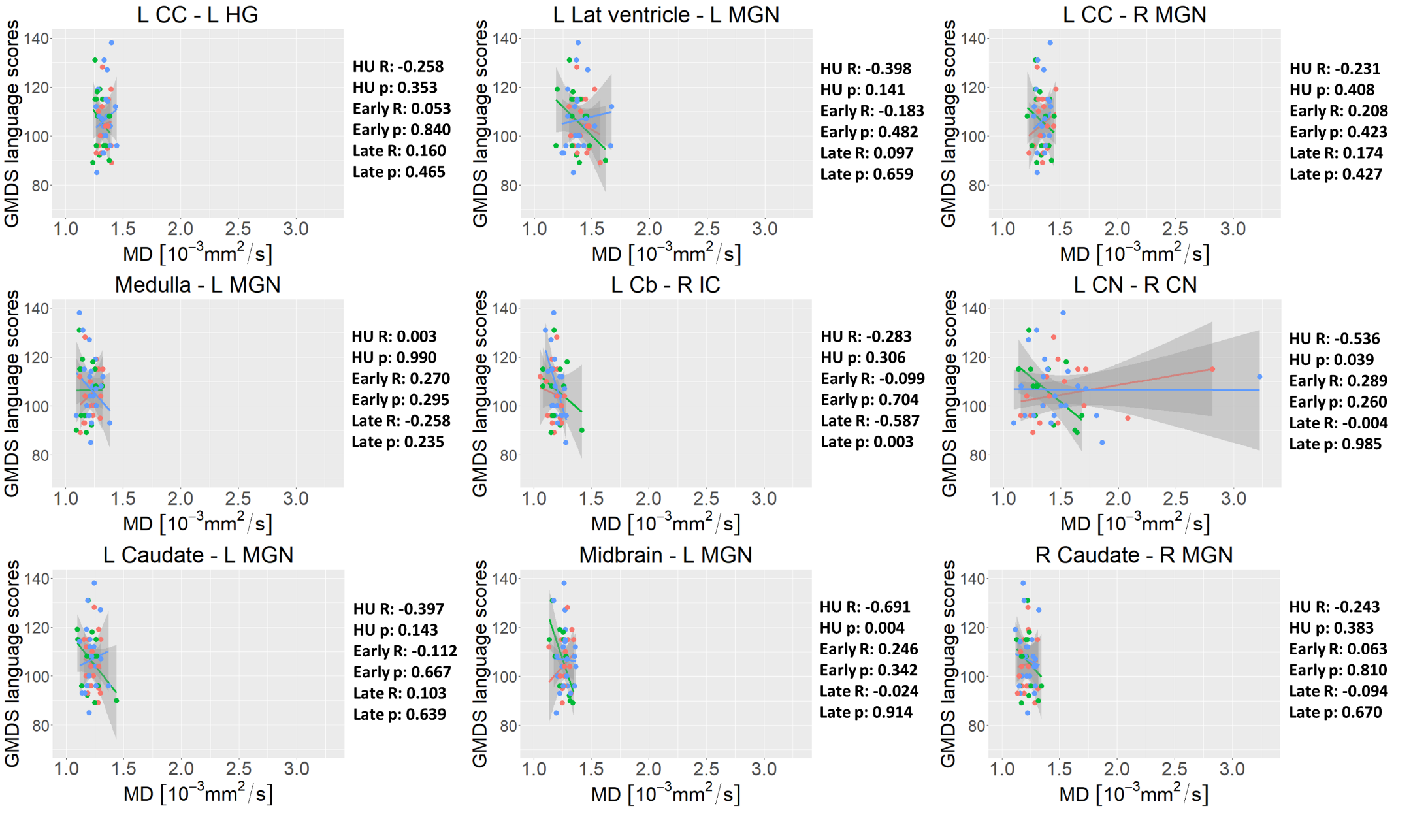

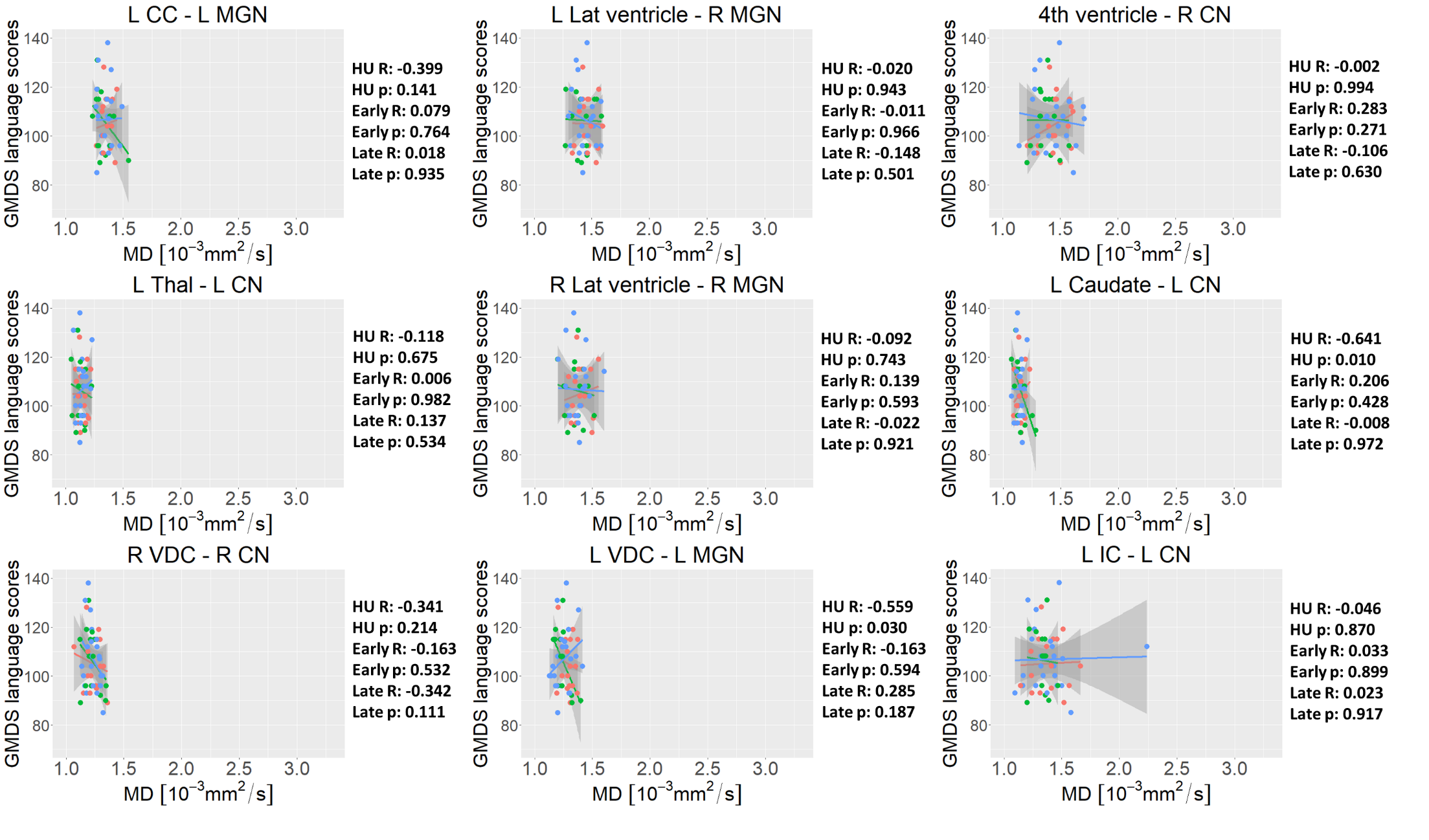

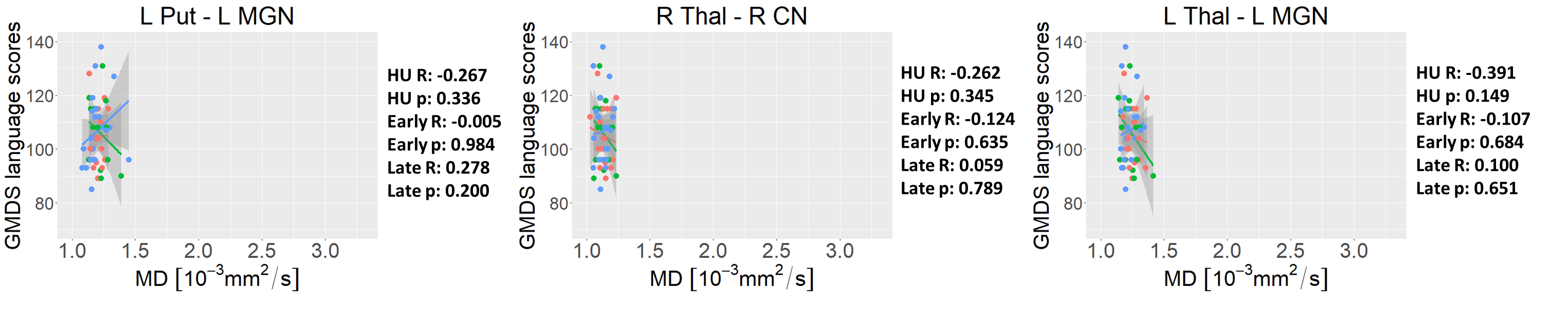

**Supplementary figure 15:** The relationship between mean diffusivity (MD) and language outcomes according to ART exposure groups, for tracts in which we formerly observed differences in MD between iHEU-pre and/or iHEU-post compared to iHU. Green = iHU; Red=iHEU-pre (early); Blue = iHEU-post (late).

**Cohen’s d calculations to compare effect size between this study and other published studies**

Pooled standard deviations and Cohen’s d could be calculated based on the following equations, as in previous literature ^60^.

**Supplementary equation 1:**

$$S\mathrm{pooled}=\sqrt{\frac{\left( n_{1}-1 \right)s_{1}^{2}+\left( n_{2}-1 \right)s_{2}^{2}+\ldots+\left( n_{k}-1 \right)s_{k}^{2}}{n_{1}+n_{2}+\ldots+n_{k}-k}}$$

**Supplementary equation 2:**

$$\mathrm{Cohen}^{'}s d=\frac{m_{1}-m_{2}}{S_{pooled}}$$

Where S_pooled_ = the pooled standard deviation, n_1_= number of subjects in group 1, n_2_= number of subjects in group 2, s_1_= standard deviation in group 1, s_2_= standard deviation in group 2, k= number of groups, m_1_= mean in group 1 and m_2_= mean in group 2.
